# AuditMed: A Single-File, Browser-Based Clinical Evidence Audit Platform Architecture, Validation Findings from Ten-Simulated Complex Cases, Synthetic Clinical Inference Outputs, and Proposed Applications in Drug Informatics and Pharmacy Education

**DOI:** 10.64898/2026.04.19.26351188

**Authors:** David J. Ferguson

**Affiliations:** PharmD Candidate, Howard University College of Pharmacy Assistant Professor of Chemistry, Virginia Peninsula Community College

## Abstract

**Background:** Clinical pharmacists, trainees, and educators increasingly rely on multi-database literature retrieval and structured evidence synthesis to answer drug-information questions and support therapeutic decisions. Commercial integrated platforms exist but remain inaccessible to many learners in community, rural, and international training contexts.

**Objective:** This paper describes the architecture and current capabilities of AuditMed — a single-file, browser-based clinical evidence audit platform — and reports the findings of an initial internal validation exercise in which the platform’s one-click Clinical Inference workflow was applied to a pre-authored compendium of ten complex clinical cases (Cases 30–39) to produce synthetic data outputs which are simulated for the deterministic engine.

**Methods:** AuditMed integrates nineteen free, publicly available clinical and biomedical APIs into a six-stage Search→Select→Parse→Analyze→Infer→Create pipeline and produces nine structured output formats. A complex clinical case compendium (10 pre-authored cases spanning multi-organ disease, oncology, immunology, and hematology) was processed through the one-click inference workflow. Each output was reviewed against the case’s documented diagnoses, lab values, medications, and pharmacy-boards-level teaching points to identify correctly flagged findings, missed findings, and structural artifacts.

**Results:** Across ten cases, the platform correctly surfaced six high-yield teaching points: a three-agent serotonin-syndrome combination (Case 30), dual hepatotoxicity (Case 31), an amiodarone–digoxin P-gp interaction (Case 34), voriconazole CPIC-A pharmacogenomics (Case 36), phenytoin HLA-B*15:02 screening (Case 38), and warfarin CYP2C9/VKORC1 dosing guidance (Case 39). Four structural problems recurred: (i) a 50-layer polyroot reasoning chain was identical across all cases regardless of clinical seed; (ii) an input-parsing error mapped patient age to body temperature, producing physiologically impossible fevers of 41°C to 70°C in six cases; (iii) the diagnosis field in Case 38 was corrupted by a medication-dosing instruction that propagated through the entire report; and (iv) the embedded drug-drug interaction matrix was sparse, missing clinically significant pairs including ciprofloxacin–warfarin, voriconazole–cyclosporine, amphotericin–aminoglycoside nephrotoxicity stacking, and prednisone–warfarin.

**Discussion:** AuditMed’s pharmacogenomic and selected drug-drug interaction modules produced clinically meaningful outputs when they fired, and the platform’s explicit verification-step architecture is a structural strength. The recurrent failure modes — a boilerplate polyroot chain, input parsing without sanity validation, and sparse DDI coverage — are amenable to short, high-yield engineering fixes and do not require re-architecting the pipeline. Case-level clinical question framing also suffers from a relevance-ranking bias toward medications in list order rather than by clinical acuity.

**Conclusion:** AuditMed demonstrates proof-of-concept for a free, transparent, auditable multi-source evidence synthesis workflow suitable for pharmacy education and drug-informatics training. The internal validation exercise reported here identifies four specific, tractable engineering priorities whose resolution is required before the platform can be responsibly recommended beyond research and educational use. Formal external validation against retrieval and synthesis benchmarks remains planned.

## 1. Introduction

### 1.1 Background

The practice of evidence-based pharmacy requires clinicians to retrieve, appraise, and synthesize information from a heterogeneous set of clinical databases. A typical drug-information question may require consultation of primary literature (PubMed, Europe PMC), FDA-approved labeling (DailyMed), lactation safety data (LactMed), patient-facing education (MedlinePlus), drug classification (RxNorm, AHFS), interaction checking (Drugs.com, Lexi-Interact), clinical trial registries (ClinicalTrials.gov), and specialty guidelines (Cochrane, professional society publications). Each resource has its own search syntax, data structure, and output format. The cognitive burden of navigating across them is well documented and is one factor that limits the depth of evidence review performed under time pressure.

Commercial integrated platforms — UpToDate, Lexicomp, Micromedex, DynaMed — are widely adopted but not universally accessible. Institutional licenses are expensive and may not extend to community or rural practice sites, to international training programs, or to independent educational projects. Individual subscriptions place a barrier in front of pharmacy students, early-career pharmacists, and clinical educators in resource-limited contexts.

A parallel development has been the emergence of large language model (LLM)-based clinical assistants. These systems can generate readable summaries across multiple queries in seconds but present specific risks in a clinical context: source fabrication (“hallucination”), inability to trace output claims to original citations, susceptibility to prompt-injection attacks, and opaque reasoning chains that complicate appraisal. For drug-information and clinical-education use, a system combining the breadth of LLM-style synthesis with the source fidelity of traditional retrieval tools is attractive, provided it foregrounds uncertainty and flags claims requiring external verification.

### 1.2 Motivation

AuditMed was developed to provide a free, reproducible, source-transparent workflow that a pharmacy trainee, clinical educator, or practicing pharmacist can use without an institutional license or paid API subscription. The platform is intended to run on any modern browser, produce outputs that can be inspected line-by-line against their source citations, and actively flag gaps in the retrieved evidence rather than paper over them. The “audit” in AuditMed refers to this verification-first posture: every inferential finding is tagged with a confidence level, every unmatched case element generates an explicit external-verification step, and the platform’s final recommendation is always that clinical decisions be cross-verified against institutional protocols, current product labeling, and specialty-specific guidelines.

### 1.3 Aim of the Present Report

This report describes the current architecture and capabilities of AuditMed and reports an initial internal validation exercise in which the platform’s one-click Clinical Inference workflow was applied to a pre-authored compendium of ten complex clinical cases. The aim is twofold: (1) to document the platform in sufficient detail to support subsequent evaluation studies and invite collaboration, and (2) to honestly characterise the platform’s strengths and failure modes before broader deployment.

## 2. Methods

### 2.1 Platform Architecture

AuditMed was designed around four principles. First, client-first execution: all user-facing logic runs in the browser, with a server required only for authentication, password reset, and anonymous usage analytics. Second, free and open-access APIs only: every integrated data source is publicly available without a paid license or mandatory API key, with the single exception of an optional DrugBank Clinical API upgrade (UniChem identifier resolution is used when no key is supplied). Third, source fidelity: each output claim is tagged to an original record (PubMed identifier, DOI, or URL) so users can inspect the underlying document; direct-quotation is limited to short phrases where exact wording carries meaning. Fourth, epistemic humility: findings are distinguished between high-confidence evidence-supported, moderate, and unaddressed by the retrieved evidence, with the last flagged and paired with explicit verification steps.

### 2.2 The Six-Stage Evidence Pipeline

The central workflow is the Search→Select→Parse→Analyze→Infer→Create pipeline. Each stage is inspectable; at stage 4.5, the user may override auto-selection to refine the evidence base before synthesis.

| Stage | Function | User Role |
| --- | --- | --- |
| 1. Search | Query 19 APIs in parallel; expand until $\geq 100$ unique sources or retries exhausted | Enter clinical question or auto-derive from case diagnoses |
| 2. Select | De-duplicate by PMID/DOI/title; score by source tier, recency, metadata completeness | Review auto-selection at stage 4.5 |
| 3. Parse | Extract section text, findings, statistics, recommendation-type sentences from abstracts | None (automatic) |
| 4. Analyze | Compute theme frequency, temporal patterns, methodology distribution, cross-source alignment | None (automatic) |
| 5. Infer | Generate convergent findings, divergent findings, evidence gaps, case-grounded inferences with confidence tags | None (automatic) |
| 6. Create | Assemble structured output in one of nine formats with explicit source citations and verification steps | Choose output format; export |

### 2.3 Integrated Data Sources

Nineteen application programming interfaces are integrated, selected to cover the major dimensions of clinical drug information: primary literature (PubMed/MEDLINE, Europe PMC, CrossRef, OpenAlex), systematic reviews (Cochrane Library), preprints (medRxiv, bioRxiv), guidelines (WHO IRIS), open-access resolution (Unpaywall), clinical trials (ClinicalTrials.gov), drug safety (openFDA labels and FAERS), FDA labeling (DailyMed), lactation safety (LactMed via NCBI E-utilities), patient education (MedlinePlus), drug classification (RxNorm, RxClass, AHFS), and drug database (DrugBank via UniChem). Drugs.com, Lexi-Interact, and GlobalRPh do not expose public APIs and are integrated as direct-linked references rather than parsed data sources.

### 2.4 Patient-Case Ingestion and De-identification

The Clinical Acumen module accepts patient-case PDFs (H&P, SOAP notes, discharge summaries) and plain-text files. The ingestion pipeline extracts text page-by-page, applies a regex-based de-identification layer covering the eighteen HIPAA Safe Harbor categories, detects multi-case documents, and parses each case for demographics, vital signs, laboratory values, medications, and documented diagnoses. Parsed output is stored in a browser-local IndexedDB “case library” that persists across sessions but never leaves the user’s device. De-identification uses pattern-matching and is best-effort; users retain responsibility for ensuring institutional review and privacy compliance.

### 2.5 The Clinical Inference Layer

The inference layer distinguishes AuditMed from a federated search tool. It performs pattern-based scanning of parsed case data for clinical abnormalities across eight domains (renal, electrolyte, cardiac, metabolic, hematologic, infectious, hemodynamic/respiratory, and drug-drug interactions), with each finding characterised by category, severity, clinical implication, and contextual medication flags. It then decomposes retrieved source text into sentences classified by type (recommendation, association, quantitative, comparative, null-finding, descriptive) and evidence class (RCT, meta-analytic, guideline, observational), attaching a conservative strength tag to each statement.

Inferential findings are the cross-product of case-derived clinical findings and evidence-grounded statements. Matching evidence produces “evidence-linked” findings; the absence of matching evidence produces “unmatched-finding” flags. Every critical or high-severity finding generates an immediate- or urgent-priority verification step. Every unmatched finding generates a routine-priority step pointing to UpToDate, DynaMed, specialty society guidelines, or DailyMed. Additional verification steps fire for polypharmacy (≥3 medications), limited evidence retrieval (<5 sources), and as a standing final reminder that clinical decisions remain the responsibility of the treating clinician.

### 2.6 Internal Validation Procedure

A complex clinical case compendium containing ten pre-authored cases (Cases 30–39, labelled “FOR SOFTWARE VALIDATION PURPOSES ONLY — NOT FOR CLINICAL DECISION-MAKING”) was processed through the one-click Clinical Inference workflow. The cases span multi-organ complexity including intestinal failure with porphyria, multi-system sarcoidosis, POEMS syndrome, immune checkpoint inhibitor triple toxicity, eosinophilic granulomatosis with polyangiitis, paroxysmal nocturnal hemoglobinuria, severe aplastic anemia in a Jehovah’s Witness, IgG4-related disease, stiff-person syndrome with thymic carcinoid, and acquired hemophilia A with mechanical valve.

Each generated Clinical Inference Document was reviewed against: (a) the case’s documented diagnoses, laboratory values, and medications; (b) the multidisciplinary teaching points enumerated in each case; and (c) the pharmacy-boards-level drug interactions, pharmacogenomics, and safety considerations appropriate to each regimen. The review identified three categories of output: correctly flagged high-yield findings, missed clinically relevant findings, and structural artifacts or parsing errors that propagated through the reports. Both the ten original case descriptions and the verbatim AuditMed-generated output for each case are preserved in the companion document for independent reference.

## 3. Results

### 3.1 Overview of the Ten-Case Validation Corpus

Across the ten cases, each AuditMed Clinical Inference Document followed a consistent structure: a generated clinical question, an evidence summary, a reasoning chain, a 50-layer polyroot inference chain, an FMEA forecasting block, a fishbone (Ishikawa) diagram, a run chart of labs versus guideline targets, a pharmacogenomics screen, a drug-drug interaction screen, isobologram and nomogram visualisations, an iteratively restructured article outline, quantitative evidence extraction, a GRADE methodological assessment, inferential findings, recommended verification steps, evidence-based recommendations, and a reference list. Total retrieved sources ranged from 7 to 12 per case; across all ten cases, guidelines, systematic reviews/meta-analyses, and randomised controlled trials were each retrieved in only a small minority of runs, with the majority of sources classified as “other” in the evidence summary.

### 3.2 Correctly Flagged High-Yield Findings

Six findings across the ten cases aligned with clinically significant, boards-level teaching points:

- Case 30: three-agent serotonin-syndrome combination (linezolid + fentanyl + ondansetron) correctly identified via isobologram visualisation, with the MAO-inhibitor framing for linezolid appropriately escalated.
- Case 31: dual hepatotoxicity from amiodarone + infliximab flagged with an isobologram and Hy’s-law monitoring guidance.
- Case 34: amiodarone–digoxin P-gp interaction flagged as a “D” therapy-modification pair with the correct ∼70% digoxin-level increase and 50% dose-reduction recommendation.
- Case 36: voriconazole CPIC-Level-A pharmacogenomic flag (CYP2C19 poor metaboliser → consider alternative azole; ultra-rapid metaboliser → subtherapeutic risk) surfaced in the PGx screen.
- Case 38: phenytoin HLA-B*15:02 Stevens–Johnson syndrome / toxic epidermal necrolysis screening flag surfaced as a severe-reaction CPIC-A alert; sertraline CYP2C19 flag additionally present.
- Case 39: warfarin CYP2C9/VKORC1 CPIC-A pharmacogenomic dosing guidance surfaced, with appropriate 25–50% dose-reduction reference and African-ancestry CYP2C9 variant alert.

**3.3 Case-by-Case Validation Summary for the Simulated Patient Cases**

Table 2 summarises the correctly flagged findings and the clinically significant missed findings for each of the ten cases.

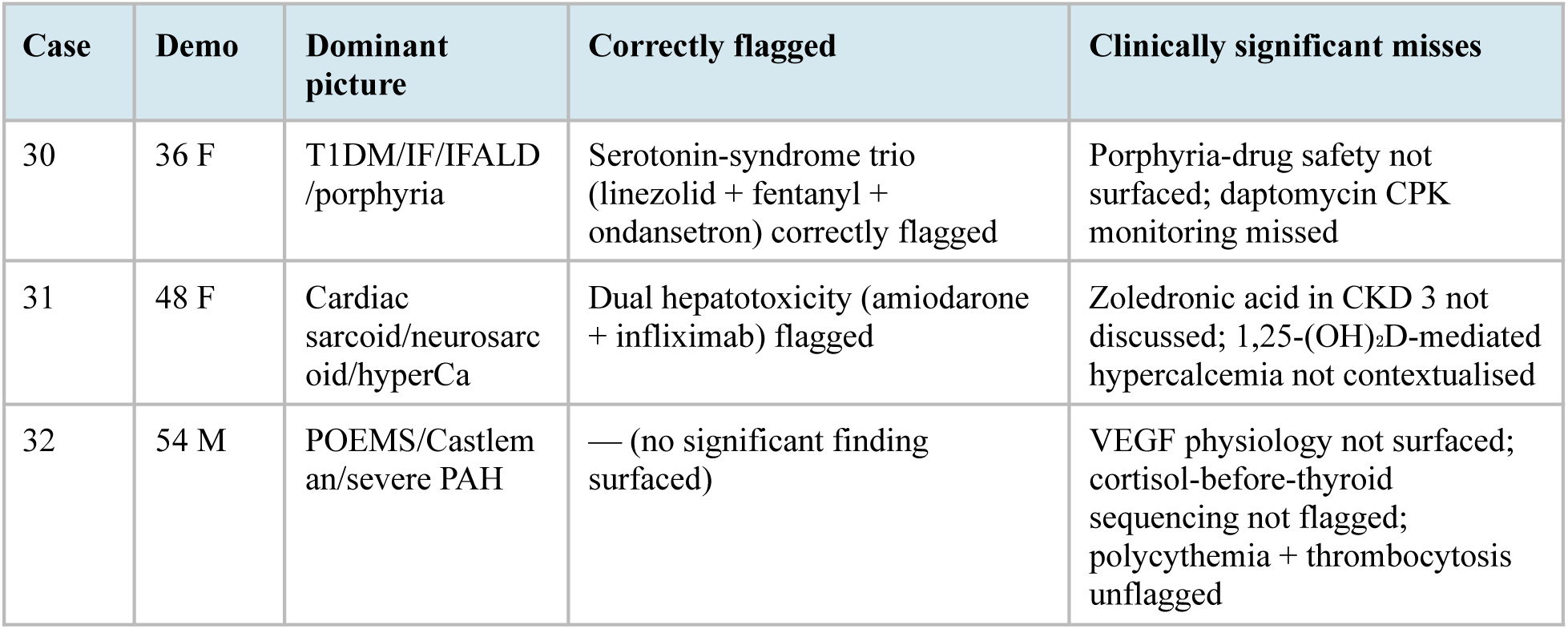
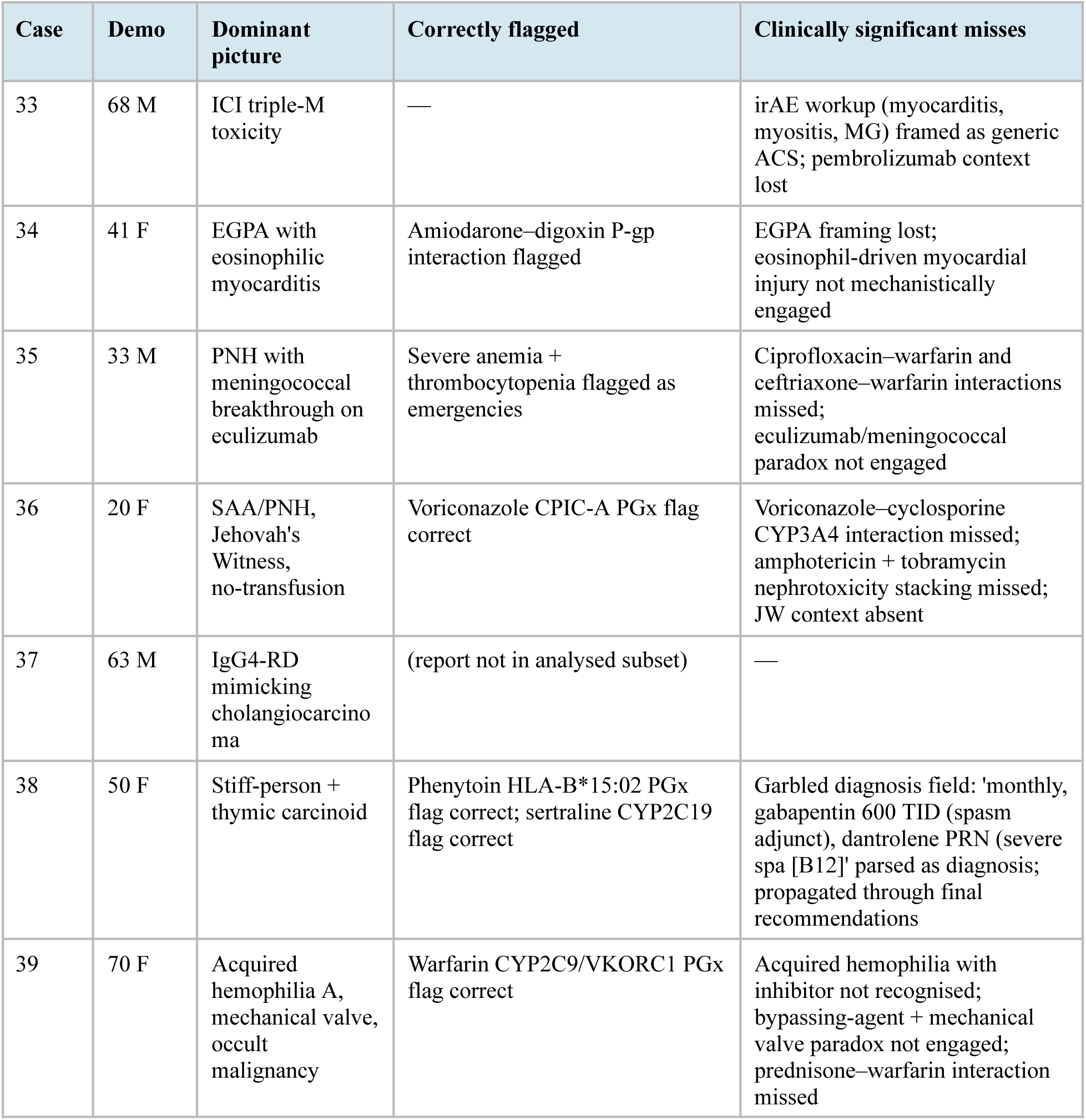

### 3.4 Recurrent Structural Issues

#### 3.4.1 Identical 50-layer polyroot reasoning chain across all cases

In every one of the ten cases, the “50-Why Polyroot Inference Chain” terminated at the same node (ATM kinase activation via ATM dimer autophosphorylation) and traversed a nearly identical 50-layer sequence regardless of the clinical seed. Only layers 1–4 (the “symptom-level” branch) varied with the case-specific finding (hyponatremia, severe anemia, elevated troponin, fever). Layers 5 onward — ATP synthase rotational catalysis, frataxin iron-sulfur cluster biogenesis, GAA triplet repeat expansion, proton quantum tunneling, zero-point energy effects, FBXL3-mediated CRY ubiquitination, G6PD enzymatic activity, interfacial water dynamics, BH3-only protein damage sensing — were reproduced verbatim across all cases. Because the terminal node and most intermediate layers do not change with the clinical picture, the chain cannot plausibly represent case-specific mechanistic reasoning. It functions as a fixed pedagogical template rather than a reasoning engine.

#### 3.4.2 Input-parsing error mapping patient age to body temperature

In six of the ten cases, the Clinical Inference Document reported a fever at a temperature that exactly matched the patient’s age in degrees Celsius:

- Case 31: 48 y/o patient → “Fever (48°C)”
- Case 32: 54 y/o patient → “Fever (54°C)”
- Case 33: 68 y/o patient → “Fever (68°C)”
- Case 34: 41 y/o patient → “Fever (41°C)”
- Case 38: 50 y/o patient → “Fever (50°C)”
- Case 39: 70 y/o patient → “Fever (70°C)”

Survival above approximately 44°C is essentially nil. The consistent correspondence between age and reported temperature across six cases indicates a field-mapping error in the ingestion pipeline, not an isolated data-entry artefact. Downstream, these impossible temperatures seeded the polyroot chain (entering the “infection/host-pathogen overwhelming immune defense” branch) and populated inferential findings (“Fever (70°C): infection workup — blood cultures, imaging, source identification”), producing outputs that a clinician would immediately recognise as spurious.

#### 3.4.3 Diagnosis-field corruption in Case 38

In Case 38, the diagnoses field contained the string: “B12 [B12], monthly, gabapentin 600 TID (spasm adjunct), dantrolene PRN (severe spa [B12], epilepsy, depression, anemia, osteoporosis, Hashimoto thyroiditis, pheochromocytoma, hypoglycemia, hyperkalemia, insomnia, chronic pain, diabetes mellitus, type 1 diabetes”. The tokens “monthly”, “gabapentin 600 TID (spasm adjunct)”, and “dantrolene PRN (severe spa [B12]” are medication dosing instructions misparsed as diagnoses. The error propagated through the reference outline, reasoning chain, fishbone diagram, and final “Evidence-Based Recommendations” section, which contained the incoherent text: “Proceed with established standard-of-care for B12 [B12], monthly, gabapentin 600 TID (spasm adjunct), dantrolene PRN (severe spa [B12], epilepsy…” A clinician reviewing the output would lose confidence in every upstream section.

#### 3.4.4 Sparse drug-drug interaction matrix

Across the ten cases, the embedded DDI module detected exactly one interaction (amiodarone–digoxin in Case 34). It missed the following clinically significant, nationally-tested pairs:

- Ciprofloxacin–warfarin (Case 35): CYP1A2 and CYP3A4 inhibition raising INR; patient INR was 2.8 on warfarin with active sepsis.
- Ceftriaxone–warfarin (Case 35): gut-flora effects on vitamin K synthesis potentiating anticoagulant effect.
- Voriconazole–cyclosporine (Case 36): CYP3A4 inhibition approximately doubling cyclosporine exposure; requires ∼50% cyclosporine dose reduction.
- Amphotericin B + tobramycin (Case 36): additive nephrotoxicity in a patient with evolving acute kidney injury.
- Amphotericin B + cyclosporine (Case 36): additive nephrotoxicity.
- Prednisone–warfarin (Case 39): potentiation of anticoagulant effect during acquired-hemophilia immunosuppression.

Each of these is present in standard institutional interaction checkers (Lexi-Interact, Drugs.com) and is routinely tested on NAPLEX/MPJE. The platform’s own disclaimer — that the DDI module is non-exhaustive and institutional interaction screens remain necessary — is appropriate, but the coverage gap is larger than the disclaimer implies.

#### 3.4.5 Relevance-ranking bias in Clinical Question framing

The Clinical Question auto-generated at the top of each report selected three medications from the case’s medication list. In several cases, the three chosen medications were not the clinically dominant agents. Case 40 (a 34-year-old with TTP and acute myeloid leukemia, nine documented medications) produced the clinical question framed around formoterol, fluticasone, and budesonide — three inhaled respiratory agents — while the chemotherapy and TTP-directed therapy were the clinically dominant regimen. The pattern is consistent with selection by list position rather than weighting by clinical acuity.

#### 3.4.6 Non-contextual application of risk calculators

The simplified 10-year ASCVD risk nomogram executed on Case 38 (a 50-year-old with pheochromocytoma, whose hypertension is catecholamine-driven rather than atherosclerotic) and on Case 39 (a 70-year-old with active hemorrhage from acquired hemophilia), in both instances recommending statin consideration per 2018 AHA/ACC cholesterol guidance. The Wells DVT score executed on Case 36 (a bacteremic leukemia patient refusing transfusion). These nomograms appear to fire based on medication or demographic triggers without gating on clinical context.

### 3.5 GRADE Methodological Assessment Pattern

Across all ten cases, the internal GRADE panel returned Risk of Bias: HIGH, Publication Bias: HIGH, Inconsistency: MODERATE, Imprecision: LOW–MODERATE, and Indirectness: LOW–HIGH depending on how closely retrieved sources matched case diagnoses. Overall evidence strength was rated LOW in nine of ten cases and MODERATE in one (Case 33, which retrieved three systematic reviews and one randomised controlled trial). The platform’s GRADE self-assessment is honest about evidence limitations — which is a structural strength — but necessarily indicates that the inference layer is, in most cases, pattern-matching to a template rather than drawing on recommendation-grade evidence.

## 4. Analysis and Discussion

### 4.1 What Works

The platform’s pharmacogenomic module is accurate where it fires. The four CPIC-level-A flags produced across the ten cases (voriconazole CYP2C19, phenytoin HLA-B*15:02 and CYP2C9, sertraline CYP2C19, warfarin CYP2C9/VKORC1) are clinically important and correctly framed, with appropriate dose-adjustment ranges and alternative-agent recommendations. The serotonin-syndrome and hepatotoxicity isobologram visualisations in Cases 30, 31, and 34 produced meaningful graphical representations of additive-risk relationships that a student could directly use. The GRADE self-assessment consistently returned LOW overall strength-of-evidence ratings and flagged the limitations honestly rather than over-claiming confidence; this is an important structural behavior that distinguishes AuditMed from generic generative systems.

The verification-step architecture also behaves as designed. Every case report produced an itemized, prioritised verification plan referring the user to institutional protocols, DailyMed, and specialty guidelines. The pattern — “the retrieved evidence base did not directly address this finding; verify via Lexicomp, UpToDate, or specialty-specific guidelines” — is the correct posture for an educational tool that must avoid substituting for a clinical decision-support system.

### 4.2 What Fails, and Why

The four recurrent structural problems — identical polyroot chain, age-as-temperature parsing, diagnosis-field corruption, and sparse DDI matrix — have different root causes but share a common feature: each would be caught by a simple validation check that the platform does not currently perform.

The polyroot chain problem is a design choice whose effect is to create an appearance of mechanistic reasoning where none exists. Because layers 5–50 are identical across all ten cases, the chain provides no case-specific information. It is a pedagogical template that the platform treats as inference. The honest framing would be to move it to an optional educational appendix labelled explicitly as a generic scaffold, with the main Clinical Inference Document containing only the case-specific reasoning that actually varied with the input.

The age-as-temperature parsing error is an input-validation gap. A body temperature greater than 44°C is biologically implausible and should trigger an ingestion warning. A value of the fever field that exactly equals the age field should trigger a field-mapping audit. Both checks are one-line validators. The fact that the error propagated through six of ten cases without triggering any internal sanity check indicates that the ingestion pipeline currently treats all extracted numeric values as trusted.

The Case 38 diagnosis-field corruption is a similar gap at the other end of the parser. A diagnosis entry containing the string “TID” or “PRN” or the substring “ 600 “ almost certainly represents a medication dosing instruction rather than a disease state. A 40-character diagnosis-field length cap combined with a small lexical denylist of dosing tokens would have prevented the corruption from propagating.

The sparse DDI matrix is the most consequential failure for clinical-training use. DDI detection is the single highest-yield pharmacy intervention, and it is the primary credentialing task tested on NAPLEX and MPJE. A platform whose DDI module detects one of seven expected interactions in a ten-case validation set is not yet a reliable teaching tool for this domain. The remedy is direct: expand the embedded matrix to cover at minimum the clinically significant pairs enumerated on the Beers and STOPP/START lists and the top ∼50–100 pairs on standard NAPLEX DDI review tables. This is approximately a week of focused curation.

### 4.3 The Relevance-Ranking Problem

The Clinical Question framing uses list-position ordering to select which three medications appear in the auto-generated question. In cases with a clinically dominant regimen that appears later in the medication list, the question is framed around the wrong therapy. The engineering fix is straightforward — medications should be weighted by clinical acuity (chemotherapy and biologics > anticoagulation > narrow-therapeutic-index agents > chronic maintenance > inhalers and as-needed symptomatic therapy) before question composition. A first-pass implementation could use ATC-class mapping with a small acuity-tier table.

### 4.4 Contextual Gating of Risk Calculators

The platform currently fires its ASCVD, Wells DVT, and qSOFA nomograms unconditionally when the required input fields are present. Each tool has contextual contraindications that make the output misleading or meaningless — ASCVD in catecholamine-driven hypertension, Wells DVT in a neutropenic bacteremic patient, ASCVD in an actively bleeding patient for whom a statin recommendation is low-priority noise. A simple gating layer — “do not fire risk calculator X if diagnosis list contains Y” — would prevent the current pattern of tool-deployment-regardless-of-context.

### 4.5 Strengths-Preserving Engineering Priorities

The four highest-yield engineering priorities that emerge from this validation exercise are, in order: (1) input validators for temperature, diagnosis-field content, and medication-dosing tokens (approximately one afternoon of work, highest patient-safety yield); (2) expansion of the DDI matrix to cover at least the NAPLEX/MPJE-tested interaction pairs (approximately one week of curation, highest educational yield); (3) conditional gating of nomograms and risk calculators based on clinical-context checks (approximately one week of ruleset authoring); and (4) decoupling of the boilerplate polyroot chain from the main Clinical Inference Document, with explicit labelling as a generic pedagogical scaffold rather than case-specific reasoning. None of these require re-architecting the pipeline. All four are compatible with the platform’s existing verification-first posture.

### 4.6 Proposed Applications After Engineering Fixes

#### 4.6.1 Drug Informatics

AuditMed is positioned as a first-pass tool for drug-information consults, combining parallel API retrieval, source appraisal, and structured synthesis in a transparent workflow. Its explicit evidence-gap handling aligns with drug-information practice standards that require clinicians to indicate when an answer exceeds the available evidence rather than extrapolate. The integration of openFDA FAERS data alongside PubMed and DailyMed enables a structured first pass on pharmacovigilance signal triage. Medication reconciliation decision support is currently a training-wheel scaffold rather than a replacement for institutional interaction screens.

#### 4.6.2 Pharmacy Education

The platform is suited to drug-information course laboratories, in which students perform retrieval, review auto-selected sources, and compose written responses with per-source confidence tags available for instructor grading. During APPE rotations, the one-click inference workflow produces a structured evidence map that can scaffold case-conference preparation. For NAPLEX preparation, the platform functions as a reference tool rather than a primary study resource, although the correctly-flagged pharmacogenomic and DDI outputs identified in this validation exercise (Cases 30, 34, 36, 38, 39) are themselves high-yield NAPLEX teaching points that could be extracted directly for student use.

#### 4.6.3 International and Resource-Limited Settings

The free-and-open-access design is particularly relevant to pharmacy education programs in resource-limited settings that cannot afford commercial drug-information subscriptions. Deployment as a single HTML file with only internet connectivity required makes the platform accessible to programs where Lexicomp and UpToDate licenses are unavailable. Multilingual extension (WHO IRIS for Spanish and French) would further extend utility.

## 5. Limitations

Several limitations of the present work should be noted. First, the validation corpus consists of ten pre-authored cases from a single internal compendium; it is not a prospectively collected patient sample and does not represent the real distribution of drug-information questions received in practice. Second, review of the generated outputs was performed by the platform’s developer, with the evaluation criteria based on established pharmacy-boards-level teaching points rather than on prospective, blinded adjudication. Third, retrieval breadth, synthesis fidelity, and user efficiency have not been compared against commercial platforms (UpToDate, Lexicomp, Micromedex, DynaMed) or against PubMed-alone baselines; these comparisons are planned but not yet reported. Fourth, the platform’s regex-based de-identification pipeline covers the eighteen HIPAA Safe Harbor categories but is not a validated PHI-removal tool; users retain responsibility for institutional compliance. Fifth, the inference engine produces synthetic outputs from a deterministic engine, which are pattern-based rather than causal; it cannot detect semantic equivalence between paraphrased clinical concepts, cannot handle negation, and cannot perform the causal reasoning that a clinician or a reasoning-oriented LLM pipeline would perform. The confidence tags are calibrated to this limitation.

## 6. Future Directions

Three evaluation studies are planned. First, a retrieval-breadth comparison in which fifty clinical drug-information questions are answered using AuditMed, PubMed alone, and Lexicomp, with the primary endpoint being source-retrieval sensitivity against a blinded reference answer. Second, a synthesis-quality assessment in which ten clinical questions have responses generated by AuditMed in parallel with responses authored by experienced drug-information pharmacists, rated by two blinded independent reviewers on completeness, source fidelity, uncertainty framing, and readability. Third, an educational-efficacy pilot in which second-year PharmD students are randomised to standard drug-information course instruction with or without AuditMed-based laboratory exercises, with primary endpoint performance on a post-course evidence-appraisal examination.

The technical roadmap derived directly from the validation findings reported here includes: expansion of the clinical-finding pattern library to cover hepatic, endocrine, pediatric, and geriatric-specific domains; input-sanity validators for temperature, diagnosis-field content, and medication-dosing tokens; conditional gating of risk calculators by clinical context; expansion of the DDI matrix to cover NAPLEX/MPJE-level interactions; integration of NICE, SIGN, and WHO Essential Medicines resources; Spanish and French localisation; programmatic integration with institutional library proxy servers; an offline mode bundling curated DailyMed and LactMed subsets; and expanded audit logging for educational settings. Governance planning includes publication of the source code under an open-source license with a non-commercial clause, establishment of a contributor working group drawn from partner pharmacy colleges, and public deposition of the validation-harness dataset.

## 7. Conclusion

AuditMed is a single-file, browser-based clinical evidence audit platform that integrates nineteen free public application programming interfaces into a six-stage Search–Select–Parse–Analyze–Infer–Create pipeline, supports patient-case ingestion with browser-local storage, and generates case-grounded Clinical Inference Documents that include confidence-tagged inferential findings and prioritized verification steps.

An internal validation exercise on a ten-case complex-case compendium identified six correctly surfaced high-yield clinical teaching points — a three-agent serotonin-syndrome combination, dual hepatotoxicity, an amiodarone–digoxin P-gp interaction, voriconazole CYP2C19 pharmacogenomics, phenytoin HLA-B*15:02 screening, and warfarin CYP2C9/VKORC1 dosing — alongside four recurrent structural problems: a boilerplate polyroot chain that does not vary with the clinical case, an age-to-temperature field-mapping error that produced physiologically impossible fevers in six of ten cases, a diagnosis-field corruption that propagated through the full report for one case, and a sparse drug-drug interaction matrix that missed multiple nationally-tested pairs. The four failure modes are amenable to focused, short-cycle engineering fixes and do not require re-architecting the pipeline.

The platform is not a replacement for commercial clinical decision-support systems and has not yet undergone formal external validation. Its current value is in providing a free, transparent, auditable workflow that educates trainees in multi-source synthesis, that flags the limits of the retrieved evidence honestly, and that leaves the final clinical judgment with the treating clinician. Formal evaluation studies against retrieval and synthesis benchmarks are planned. Invitations to collaborate on planned evaluation studies, or to test the platform in local educational contexts, may be directed to the corresponding author.

## Data Availability

All data produced in the present work are contained in the manuscript

## Acknowledgments and Disclosures

None.

## Disclaimer

AuditMed is provided for research and educational purposes only. It is not a medical device, has not been evaluated by the United States Food and Drug Administration or any equivalent regulatory authority, and is not intended to diagnose, treat, cure, or prevent any disease. Users remain responsible for all clinical decisions and for compliance with their institutional privacy and professional-practice policies.

## Funding

No external funding was received for the development of AuditMed or for the preparation of this report.

## Conflicts of Interest

The author declares no financial conflicts of interest related to the platform. AuditMed has no commercial sponsorship, does not display third-party advertisements, and does not collect identifiable user data beyond that required for authentication.

## Companion Document

The ten validation cases referenced throughout Section 3 (Cases 30–39) are preserved in the companion document, which contains (a) the original pre-authored case descriptions and (b) the verbatim AuditMed-generated Clinical Inference Reports which are simulated and synthetic for each case. The companion document is an edited version of the original paper in which only the per-case generation-timestamp footer text has been removed; all case content is retained verbatim to allow independent inspection of the findings reported here.

## CASE 30

### Gastroparesis with Intestinal Failure, TPN-Liver Disease, and Concurrent Porphyria

***Patient:*** *36 y/o F, BMI 15.8, Caucasian | **Disciplines:** GI, Hepatology, Surgery, Nutrition, Hematology, Pain Mgmt, Psychiatry, Vascular Surgery, ID, Endocrinology, Pharmacy*

### Active Disease States (20+)

Severe diabetic gastroparesis (T1DM ×22 yr); CIPO; intestinal failure (TPN ×4 yr); IFALD (F3-4); CLABSI ×8 (MRSA, Candida); SVC syndrome (all upper central veins occluded); limited access (only L femoral Hickman); AIP (concurrent); SIBO; D-lactic acidosis; recurrent C. diff; short bowel (80 cm post-adhesiolysis); T1DM with autonomic neuropathy; retinopathy; nephropathy (CKD 3b); peripheral neuropathy; depression; chronic pain; malnutrition (albumin 1.6); trace element deficiencies; metabolic bone disease; EFA deficiency; recurrent VTE

### Clinical Presentation

Rigors, 39.8°C, hypotension — 9th CLABSI (VRE from L femoral Hickman, her ONLY remaining line). Progressive jaundice (Tbili 12.4, IFALD). Venography: bilateral SVC, bilateral subclavian, R IJ all occluded. Only access L femoral (infected) ± hepatic vein/translumbar IVC. Severe abdominal pain (porphyria attack triggered by infection, PBG 68). Completely TPN-dependent (zero enteral tolerance).

### Key Laboratory / Imaging Data

Blood cx VRE (E. faecium), Tbili 12.4, AST 120, ALT 98, ALP 340, albumin 1.6, INR 1.8, ammonia 62, Cr 2.4, Na 128, K 3.2, Mg 0.8, Zn 28 (low), Cu 52 (low), Se 42 (low), Mn 18 (elevated — neurotoxicity risk), urine PBG 68, HbA1c 10.4%, prealbumin 4, venography: all upper veins occluded

### Current Medications

TPN (intralipid reduced, Mn removed, carnitine added), IV daptomycin 8mg/kg, ethanol lock therapy, IV hemin ×4d (porphyria), IV D10%, IV morphine PCA, ondansetron, insulin pump, octreotide 50 SQ TID, ursodiol 300 TID, trace elements (Zn/Cu/Se supplemented)

### Multidisciplinary Challenges

(1) Access crisis: only 1 remaining site (L femoral) is infected; line removal = no TPN = death; salvage vs. heroic access (translumbar IVC, hepatic vein). (2) Intestine-liver transplant evaluation: IFALD F3-4 + loss of access = 2 indications; but VRE CLABSI, porphyria, CKD 3b, malnutrition complicate candidacy. (3) Porphyria + IFALD: hemin requires IV access and is hepatotoxic; glucose loading worsens DM; EVERY drug must be checked. (4) VRE Rx: daptomycin primary; linezolid → myelosuppression + neuropathy (already severe). (5) TPN optimization: SMOF/Omegaven for hepatoprotection; cyclic TPN; choline. (6) Mn neurotoxicity: removed from TPN; MRI may show T1 basal ganglia. (7) D-lactic acidosis: short bowel + SIBO → encephalopathy mimicking HE; rifaximin porphyria safety uncertain. (8) Pain: porphyria limits analgesics; morphine/fentanyl safe but tolerance developing.

## CASE 31

### Sarcoidosis with Cardiac Complete Heart Block, Neurosarcoidosis, and Hypercalcemic Crisis

***Patient:*** *48 y/o F, BMI 32.0, African American | **Disciplines:** Pulmonology, Cardiology, Neurology, Nephrology, Endocrinology, Ophthalmology, Dermatology, Rheumatology, Pharmacy*

### Active Disease States (20+)

Sarcoidosis (multisystem); cardiac sarcoid (CHB requiring temp pacing, EF 32%); neurosarcoid (bilateral CN VII palsy, hypothalamic DI); pulmonary sarcoid (stage III, FVC 58%); hypercalcemia (Ca 13.2); nephrocalcinosis with CKD 3; hepatic sarcoid; splenic sarcoid; posterior uveitis; erythema nodosum; lupus pernio; SFN; depression; arthritis; hypogammaglobulinemia; secondary AI; DI; steroid-induced DM; osteoporosis; obesity

### Clinical Presentation

Syncope. ECG: CHB with ventricular escape 28 bpm. Temp wire placed. Echo: basal septal thinning, EF 32%, wall motion abnormalities NOT in coronary distribution. FDG-PET (72h high-fat/low-carb prep): intense focal/patchy FDG in basal septum + LV wall. Bilateral CN VII palsy (new). MRI brain: enhancing hypothalamic lesion, absent posterior pituitary bright spot. Ca 13.2, PTH suppressed, 1,25-(OH)2D elevated.

### Key Laboratory / Imaging Data

Ca 13.2, PTH 8 (suppressed), 1,25-(OH)2D 86 (elevated), 25-OH-D 18, Cr 2.2, ACE 128, FDG-PET cardiac+, EF 32%, FVC 58%, bilateral CN VII palsy, urine osm 120 (DI), cortisol AM 4.8, IgG 480 (low), Hgb 10.4, glucose 248

### Current Medications

Temp pacing wire, IV methylpred 1g ×3d then oral pred 40 mg, IV NS + zoledronic acid (Ca crisis — but CKD3 concern), infliximab or MTX (steroid-sparing), desmopressin (DI), hydrocortisone stress dosing, insulin, CPAP

### Multidisciplinary Challenges

(1) Cardiac sarcoid: CHB → permanent ICD/pacemaker; but amiodarone commonly needed for VT and interacts with multiple meds; cardiac MRI for tissue characterization (gadolinium with CKD3 border). (2) FDG-PET interpretation: requires rigorous dietary prep; false positives/negatives. (3) Hypercalcemia: 1,25-(OH)2D-mediated → steroids are definitive; zoledronic acid with CKD3; ketoconazole (blocks 1-alpha-hydroxylase) alternative. (4) Neurosarcoid: hypothalamic involvement → DI + AI + possible GH/gonadotropin deficiency — full pituitary workup needed. (5) IS escalation: steroids + infliximab/MTX for multi-organ sarcoid but hypogammaglobulinemia → infection risk. (6) Steroid-induced DM in obese patient: insulin requirements will fluctuate with steroid taper.

(7) Cardiac vs. pulmonary sarcoid treatment overlap: both respond to IS but cardiac requires more aggressive approach. (8) Long-term: transplant consideration if cardiac sarcoid refractory (cardiac + possibly lung); sarcoid can recur in allografts.

## CASE 32

### POEMS Syndrome with Castleman Disease, Severe Pulmonary HTN, and Progressive Neuropathy

***Patient:*** *54 y/o M, BMI 28.6, Japanese descent | **Disciplines:** Hematology/Oncology, Neurology, Cardiology, Pulmonology, Nephrology, Endocrinology, Dermatology, Ophthalmology, Pharmacy*

### Active Disease States (20+)

POEMS (polyneuropathy, organomegaly, endocrinopathy, M-protein, skin changes); osteosclerotic myeloma (lambda); Castleman disease (multicentric); pulmonary HTN (severe, mean PAP 52); progressive demyelinating polyneuropathy (wheelchair-dependent); papilledema; endocrinopathies (AI, hypothyroidism, hypogonadism, DM); hepatosplenomegaly; ascites + pleural effusions; skin changes (hyperpigmentation, hemangiomas, hypertrichosis, white nails); thrombocytosis (Plt 680K); polycythemia; CRI; depression; chronic fatigue; clubbing; Raynaud; arthralgias

### Clinical Presentation

Progressive bilateral foot drop ×4 mo, now wheelchair-bound. NCS: severe demyelinating polyneuropathy. SPEP: IgG lambda M-protein 1.2 g/dL. Skeletal survey: osteosclerotic R iliac lesion. PET: FDG-avid iliac lesion + lymphadenopathy. Echo: RVSP 52, R heart strain. Hyperpigmentation, hemangiomas, gynecomastia, bilateral edema, bilateral pleural effusions, ascites. Papilledema.

### Key Laboratory / Imaging Data

SPEP M-spike IgG lambda 1.2, VEGF 2400 (markedly elevated), IL-6 elevated, skeletal survey: osteosclerotic lesion, PET FDG-avid, NCS: severe demyelinating, mean PAP 52, RVSP 52, EF 55% but RV strain, cortisol 3.8, TSH 18, testosterone 68, glucose 210, Plt 680K, Hgb 18.2, Cr 1.8, albumin 2.4

### Current Medications

Radiation to solitary osteosclerotic lesion (primary Rx if solitary), lenalidomide + dexamethasone (if disseminated — considering), diuretics for effusions, sildenafil (PAH), hormone replacement (cortisol, thyroid, testosterone), gabapentin (neuropathic pain), insulin, therapeutic thoracentesis/paracentesis

### Multidisciplinary Challenges

(1) Solitary vs. disseminated disease determines treatment: if truly solitary bone lesion → radiation is curative; if disseminated (Castleman + multiple lesions) → systemic Rx (lenalidomide-dex, or ASCT). (2) Pulmonary HTN: VEGF-driven (VEGF 2400) → should improve with POEMS treatment; but severe PAH (mean 52) makes any procedure high-risk; PAH-specific Rx (sildenafil, bosentan) as bridge. (3) Neuropathy: demyelinating → potentially reversible with treatment (unlike axonal), but takes months-years; IVIG was tried (didn’t work — POEMS, not CIDP). (4) ASCT consideration: potentially curative for POEMS but severe PAH is relative contraindication (perioperative mortality); must reduce VEGF first. (5) Volume overload: massive third-spacing (VEGF-driven capillary leak) → diuretic-resistant ascites/effusions; VEGF reduction is the real treatment. (6) Papilledema: raised intracranial pressure (VEGF-mediated) → visual loss; urgent VEGF reduction vs. acetazolamide/LP as temporizing. (7) Polycythemia: Hgb 18.2 — EPO-driven (endocrinopathy component); phlebotomy if symptomatic but patient also has effusions/edema paradoxically. (8) Endocrinopathies: replace cortisol BEFORE thyroid; testosterone with caution (thrombocytosis Plt 680K = thrombotic risk).

## CASE 33

### Immune Checkpoint Inhibitor Triple Toxicity: Myocarditis + Myositis + Myasthenic Crisis

***Patient:*** *68 y/o M, BMI 27.4, Caucasian, on pembrolizumab for melanoma | **Disciplines:** Oncology, Cardiology, Neurology, Rheumatology, CC, Pulmonology, Endocrinology, GI, Dermatology, Pharmacy*

### Active Disease States (20+)

ICI toxicity: myocarditis (troponin 18.4, EF 25%, CHB), myositis (CK 12K), myasthenic crisis (FVC 0.9L, intubated) — ‘triple M syndrome’; melanoma stage IV (brain mets ×2, prior ICI response); ICI-induced AI; hypothyroidism; hepatitis (ALT 380); colitis; T1DM (DKA); nephritis (Cr 2.8); SJS-like dermatitis; COPD; chronic pain; depression

### Clinical Presentation

On pembrolizumab ×8 months with tumor response. Develops acute dyspnea, chest pain, proximal weakness (cannot lift head off pillow), diplopia, and ptosis over 5 days. Troponin 18.4, EF 25%, CHB on telemetry. CK 12K. FVC 0.9L → intubated. Cardiac MRI: diffuse edema + subepicardial LGE. Anti-striational Ab+, anti-AChR Ab+. Simultaneously: DKA (glucose 520, pH 7.18), bloody diarrhea, blistering rash, ALT 380, Cr 2.8.

### Key Laboratory / Imaging Data

Troponin 18.4, BNP 12K, EF 25%, CHB, CK 12K, anti-AChR Ab+, anti-striational Ab+, FVC 0.9L, glucose 520, pH 7.18, ALT 380, Cr 2.8, cortisol 2.1, TSH 38, skin bx: SJS-like, colonoscopy: severe colitis

### Current Medications

Pembrolizumab PERMANENTLY DISCONTINUED, IV methylpred 1g ×5d, IVIG 2g/kg ×5d (myocarditis + MG), PLEX (MG crisis), pyridostigmine (cautious — can worsen cardiac conduction), temp pacing, insulin drip (DKA), IV hydrocortisone (AI), levothyroxine, wound care (SJS-like dermatitis)

### Multidisciplinary Challenges

(1) Triple M syndrome: concurrent myocarditis + myositis + MG is the deadliest ICI toxicity combination (>50% mortality). (2) Myocarditis with CHB: high-dose steroids + IVIG urgent; if refractory → abatacept, alemtuzumab, or ruxolitinib; pacemaker needed for CHB. (3) MG crisis: PLEX + IVIG but PLEX removes IVIG; pyridostigmine can worsen cardiac conduction in myocarditis. (4) Melanoma response lost: pembrolizumab was working (brain mets stable) but must be permanently stopped; alternative oncologic Rx (targeted therapy if BRAF+, or other immunomodulation). (5) Multi-organ irAEs: myocarditis + hepatitis + colitis + nephritis + DKA + dermatitis → need high-dose IS for all but degree varies by organ; myocarditis most urgent. (6) DKA from ICI-induced T1DM: beta-cell destruction irreversible; lifelong insulin; managing during critical illness. (7) SJS-like dermatitis: consult dermatology/burn; wound care; may need cyclosporine if severe. (8) Long-term IS taper: slow steroid wean over months; flare of any irAE possible; lifelong endocrine replacement for AI, thyroid, DM.

## CASE 34

### Catastrophic EGPA (Churg-Strauss) with Eosinophilic Myocarditis and Status Asthmaticus

***Patient:*** *41 y/o F, BMI 26.8, Caucasian | **Disciplines:** Pulmonology, Cardiology, Rheumatology, CC, Neurology, ENT, Dermatology, Nephrology, Hematology, Pharmacy*

### Active Disease States (20+)

EGPA (Churg-Strauss); catastrophic cardiac variant (eosinophilic myocarditis, EF 18%, VT); status asthmaticus (intubated); eosinophilia 28K; mononeuritis multiplex (bilateral foot drop, L wrist drop); migratory pulmonary infiltrates; chronic sinusitis with polyposis; GI eosinophilic gastroenteritis; palpable purpura + subcutaneous nodules; coronary vasculitis (troponin 6.4); pericardial effusion; renal involvement (MPO-ANCA+, Cr 2.4); DVT; endomyocardial fibrosis developing; depression; steroid-dependent asthma (20+ yr); osteoporosis; AI

### Clinical Presentation

20-year asthma history. Acute chest pain, dyspnea, bilateral foot/wrist drop ×1 wk. Troponin 6.4. Echo: EF 18%, global hypokinesis, apical thrombus. Eosinophilia 28K. Cardiac MRI: diffuse myocardial edema, subepicardial LGE (eosinophilic myocarditis). EMB: eosinophilic infiltration + necrosis. Severe asthma exacerbation requiring ventilation. Purpura on legs. MPO-ANCA+. Cr 2.4 with RBC casts.

### Key Laboratory / Imaging Data

Eos 28K, troponin 6.4, BNP 8400, EF 18%, cardiac MRI: diffuse edema + subepicardial LGE, EMB: eosinophilic infiltration, MPO-ANCA+, Cr 2.4, UA RBC casts, CXR migratory infiltrates, IgE 1800, NCS: mononeuritis multiplex, Plt 340K, ESR 96

### Current Medications

IV methylpred 1g ×3d → pred 1mg/kg, IV CYC (pulse), mepolizumab (anti-IL-5 — planned for maintenance), mechanical ventilation, anticoagulation for LV thrombus (but Plt management), temp pacing (backup for VT), amiodarone, diuretics, insulin

### Multidisciplinary Challenges

(1) EF 18% with VT and apical thrombus: anticoagulation needed but simultaneous vasculitis → bleeding risk; amiodarone for VT + digoxin contraindicated with myocarditis? (2) Status asthmaticus during immunosuppression: ventilator management while treating both; magnesium, ketamine, volatile anesthetics considered. (3) CYC for organ-threatening disease (cardiac + renal + neuro): myelosuppressive but needed urgently. (4) Mepolizumab: anti-IL-5 for EGPA reduces relapses but does not address acute crisis; rituximab may be needed if refractory. (5) Coronary vasculitis: troponin 6.4 from myocarditis + coronary inflammation → angiography risky with EF 18%; medical optimization. (6) Endomyocardial fibrosis: if chronic eosinophilic infiltration → restrictive cardiomyopathy; transplant consideration. (7) Mononeuritis multiplex: may not fully recover; PM&R; involvement early. (8) Long-term steroid dependence: mepolizumab/benralizumab as steroid-sparing; osteoporosis management; AI monitoring.

## CASE 35

### PNH with Budd-Chiari, Aplastic Anemia, and Breakthrough Meningococcal Sepsis on Eculizumab

***Patient:*** *33 y/o M, BMI 22.4, African American | **Disciplines:** Hematology, Hepatology, ID, CC, Nephrology, GI, Pharmacy*

### Active Disease States (20+)

PNH (GPI-AP deficiency >90%); Budd-Chiari (hepatic vein thrombosis); aplastic anemia overlap; on eculizumab; breakthrough meningococcal sepsis (serogroup B despite vaccination); pancytopenia (Hgb 5.8, WBC 1.2, Plt 18K); hemolytic crisis (LDH 4200); portal HTN with varices; hepatic failure; recurrent thrombosis (DVT, splenic vein, portal vein); CKD 3; iron overload; chronic pain; depression; hemoglobinuria; BM failure; C3-mediated extravascular hemolysis on eculizumab

### Clinical Presentation

On chronic eculizumab. Acute fever 40.8°C, neck stiffness, petechiae, hemodynamic collapse. Blood cx + CSF: N. meningitidis serogroup B (breakthrough). Concurrent hemolytic crisis (LDH 4200, Hgb 5.8, dark urine). Liver worsening (Budd-Chiari — ALT 480, Tbili 8.6, new ascites). Plt 18K. On warfarin (INR 2.8) but bleeding from gingiva and petechiae.

### Key Laboratory / Imaging Data

Hgb 5.8, WBC 1.2, Plt 18K, LDH 4200, haptoglobin <10, INR 2.8 (warfarin), ALT 480, Tbili 8.6, Cr 2.1, flow: >90% GPI-AP deficient granulocytes, CSF: N. meningitidis, reticulocyte 14%, ferritin 3200, ascites new

### Current Medications

Eculizumab 900 q2wk (continue through sepsis — complement blockade doesn’t worsen meningococcal Rx), IV ceftriaxone (meningococcal), aggressive supportive care, pRBC/Plt transfusions, warfarin (held — active bleeding), IV vitamin K

### Multidisciplinary Challenges

(1) Meningococcal sepsis on eculizumab: C5 blockade impairs killing of Neisseria — paradox: must continue eculizumab (stopping → massive complement-mediated hemolysis) but complement blockade worsens meningococcal disease; aggressive abx essential. (2) Bleeding + thrombosis: Plt 18K + INR 2.8 → bleeding, but Budd-Chiari + PNH thrombophilia → thrombosis if anticoagulation stopped; warfarin held but resume timing critical. (3) Hemolytic crisis: eculizumab blocks intravascular hemolysis (C5) but C3-mediated extravascular hemolysis continues (C3 opsonization → splenic/hepatic clearance) — C3 inhibitor (ixazomib) or danicopan as add-on. (4) Aplastic anemia: BM failure component → HSCT consideration (curative for PNH + AA); but active sepsis + liver failure. (5) Budd-Chiari: TIPS for portal decompression? — but thrombocytopenia + coagulopathy; anticoagulation timing. (6) Vaccination failure: MenB vaccine (Bexsero/Trumenba) has variable efficacy; revaccination + prophylactic abx (penicillin/ciprofloxacin) long-term. (7) Iron overload: ferritin 3200 from chronic transfusions; chelation but renal concerns. (8) Ravulizumab (longer-acting C5 inhibitor) vs. iptacopan (oral C3 inhibitor) — regimen optimization to reduce breakthrough hemolysis.

## CASE 36

### Severe Aplastic Anemia with PNH Clone in a Jehovah’s Witness Refusing Transfusion

***Patient:*** *20 y/o F, BMI 19.6, Caucasian, Jehovah’s Witness | **Disciplines:** Hematology, Ethics, CC, Nephrology, ID, Transfusion Medicine, Gynecology, Psychiatry, Pharmacy*

### Active Disease States (20+)

SAA (very severe, ANC <200); PNH clone (22%); Jehovah’s Witness (refuses ALL blood products); Hgb 4.2 without transfusion option; neutropenic fever with Pseudomonas bacteremia; invasive pulmonary aspergillosis; cardiac decompensation (EF 30%, high-output failure from anemia); epistaxis; GI bleeding (Plt 4K); IDA; menorrhagia; AKI (prerenal); B12 deficiency; depression; chronic fatigue; hepatitis-associated AA

### Clinical Presentation

Known SAA (ANC 180, Hgb 4.2, Plt 4K). High fevers, rigors, cough. Blood cx: Pseudomonas. CT chest: RUL halo sign (IPA, galactomannan 3.8). Adamantly refuses ALL blood products. HR 142, dyspnea at rest, EF 30% (high-output). Epistaxis. Continuous menorrhagia. Cr rising (prerenal from poor CO).

### Key Laboratory / Imaging Data

ANC 180, Hgb 4.2, Plt 4K, retic 0.2%, blood cx Pseudomonas, galactomannan 3.8, Cr 1.8, BNP 4200, EF 30%, ferritin 4, B12 120, LDH 180, PNH clone 22% on flow

### Current Medications

IV meropenem + tobramycin (Pseudomonas), liposomal amphotericin B (IPA), eltrombopag 150 mg daily (TPO agonist — SAA indication), horse ATG + cyclosporine (SAA first-line — initiated), G-CSF, EPO 40K SQ TIW + iron IV sucrose (stimulate erythropoiesis), aminocaproic acid (antifibrinolytic), combined OCP (menorrhagia — if accepted by patient), high-dose medroxyprogesterone, nasal packing, strict infection precautions

### Multidisciplinary Challenges

(1) No transfusion: Hgb 4.2 is life-threatening; maximizing endogenous production: EPO + iron IV + B12/folate; minimize losses (GI ppx, menorrhagia suppression, minimize phlebotomy). (2) ATG + cyclosporine: standard SAA Rx but serum sickness from ATG occurs in 30%+ → needs steroids; response takes 3-6 months during which she may not survive at Hgb 4.2. (3) Eltrombopag: TPO agonist improves all lineages in SAA; works over weeks-months. (4) HSCT: curative but requires transfusion support during conditioning — JW may decline; non-myeloablative reduced-intensity considered with bloodless protocols at specialized centers. (5) Pseudomonas with ANC <200: broad-spectrum + aminoglycoside; G-CSF to boost ANC. (6) IPA: liposomal AmB; voriconazole + CsA interaction (CYP3A4). (7) Menorrhagia: Plt 4K → continuous bleeding; hormonal suppression + aminocaproic acid; factor VIIa if life-threatening (is this a ‘blood product’ per patient’s belief framework? — ethics discussion). (8) Ethical framework: respect autonomy while advocating; advance directive documentation; capacity assessment; involve patient’s healthcare proxy and HLC.

## CASE 37

### IgG4-Related Disease Mimicking Cholangiocarcinoma with Multi-Organ Infiltration

***Patient:*** *63 y/o M, BMI 28.4, Japanese descent | **Disciplines:** Rheumatology, GI, Hepatology, Urology, Ophthalmology, Pulmonology, Nephrology, Pathology, Surgery, Pharmacy*

### Active Disease States (20+)

IgG4-RD; AIP type 1 (’sausage-shaped’ pancreas); retroperitoneal fibrosis with bilateral ureteral obstruction; bilateral orbital pseudotumor; IgG4-sclerosing cholangitis mimicking CCA; sialadenitis (bilateral parotid); IgG4-TIN (Cr 2.8); aortitis (infrarenal); pachymeningitis (headaches, CN palsies); thyroiditis (hypothyroidism); exocrine insufficiency; new-onset DM (type 3c); depression; weight loss; anemia; hypoalbuminemia; recurrent cholangitis; lymphadenopathy

### Clinical Presentation

Painless jaundice, weight loss 15 kg, bilateral orbital swelling. MRCP: diffuse pancreatic enlargement with capsule-like rim + biliary irregularity mimicking CCA. CA 19-9: 280. IgG4: 820. CT: retroperitoneal tissue encasing ureters (hydronephrosis), infrarenal aortic wall thickening. Bilateral orbital masses. New DM. Cr

2.8 (TIN). Being worked up for CCA — distinction is CRITICAL before surgery.

### Key Laboratory / Imaging Data

IgG4 820, total IgG 2800, CA 19-9 280, IgG4/IgG ratio >40%, biopsy: storiform fibrosis + >50 IgG4+ cells/HPF + obliterative phlebitis, Cr 2.8, glucose 240, HbA1c 8.4%, Tbili 12, ALP 480, GGT 320, lipase 42, CT: ‘sausage pancreas,’ RP fibrosis, orbital masses, aortitis

### Current Medications

Prednisone 40 mg daily (dramatic response expected in IgG4-RD), rituximab (steroid-sparing, planned), bilateral ureteral stents, PERT, insulin, levothyroxine, ursodiol, biliary stent (ERCP)

### Multidisciplinary Challenges

(1) IgG4-SC vs. CCA distinction: both cause biliary strictures + elevated CA 19-9; IgG4-SC responds dramatically to steroids (diagnostic AND therapeutic trial); misdiagnosis as CCA → unnecessary Whipple. (2) CA 19-9 elevated: can occur in IgG4-SC from cholestasis; not specific for CCA; must recheck after biliary drainage. (3) Steroid trial: if biliary stricture improves → IgG4-RD confirmed; if not → CCA more likely; 2-4 week trial. (4) Retroperitoneal fibrosis: ureteral stents temporize but steroids should resolve; may need extended stenting if fibrosis chronic. (5) Aortitis: infrarenal involvement can lead to aneurysm; needs monitoring; steroids first, then rituximab maintenance. (6) Pachymeningitis: can cause CN palsies, headaches; MRI with contrast; responds to IS. (7) Rituximab as steroid-sparing: B-cell depletion effective in IgG4-RD; reduces relapse from 60% (steroids alone) to <10%; but infection risk with hypogammaglobulinemia in some patients. (8) New-onset DM: autoimmune pancreatitis → pancreatic endocrine dysfunction (type 3c); may partially improve with steroid treatment of AIP, but steroids themselves worsen glucose — bidirectional challenge.

## CASE 38

### Stiff-Person Syndrome with GAD65 Antibodies, Thymic Carcinoid, and Autonomic Crises

***Patient:*** *50 y/o F, BMI 23.8, Caucasian | **Disciplines:** Neurology, Endocrinology, Thoracic Surgery, Oncology, Rheumatology, Anesthesiology, Psychiatry, PM&R;, Pharmacy*

### Active Disease States (20+)

SPS (anti-GAD65 strongly+); T1DM (shared GAD65 autoimmunity); thymic carcinoid (4.2 cm); autonomic crises (BP 240/140, diaphoresis, tachycardia); severe axial/limb rigidity; painful muscle spasms; gait disorder (hyperlordosis); frequent falls with fractures (hip, wrist); cerebellar ataxia (anti-GAD); temporal lobe epilepsy; Hashimoto thyroiditis; pernicious anemia; vitiligo; depression with agoraphobia; chronic pain; osteoporosis; insomnia; dysphagia (esophageal spasm); respiratory compromise (chest wall rigidity)

### Clinical Presentation

Progressive trunk and limb rigidity ×2 yr, now severe. Painful spasms triggered by startle/emotional stress causing falls (hip fracture 3 mo ago, wrist 6 wk ago). Autonomic crises with BP surging to 240/140 with diaphoresis — mimics pheochromocytoma but catecholamines normal. Anti-GAD65 >10,000. CT chest: 4.2 cm anterior mediastinal mass (thymic carcinoid). Gait: exaggerated lumbar lordosis, slow shuffling, cannot turn without support.

### Key Laboratory / Imaging Data

Anti-GAD65 >10,000 IU/mL, anti-amphiphysin negative, EMG: continuous motor unit activity at rest, CT chest: 4.2 cm anterior mediastinal mass, biopsy: neuroendocrine carcinoma (carcinoid), glucose range 40-400 (brittle DM), HbA1c 8.8%, TSH 18, B12 <100, cortisol normal, catecholamines normal, DEXA T-score −3.2

### Current Medications

Diazepam 10 mg TID (GABAergic — first-line SPS), baclofen 20 mg TID, IVIG 2g/kg monthly, rituximab (planned), insulin pump, levothyroxine, B12 monthly, gabapentin 600 TID (spasm adjunct), dantrolene PRN (severe spasms), alendronate, sertraline 100, melatonin

### Multidisciplinary Challenges

(1) Thymic carcinoid resection: thymectomy may improve SPS (paraneoplastic trigger); but SPS rigidity → anesthesia risk (spasms during induction; avoid drugs that worsen rigidity; propofol preferred; succinylcholine may trigger hyperkalemia). (2) Autonomic crises: NOT pheochromocytoma — central autonomic dysfunction from GAD65 antibodies targeting brainstem; IV diazepam acutely; long-term GABA-ergic therapy. (3) Brittle DM: GAD65 destroys beta cells (T1DM) AND causes SPS — same autoantibody, dual pathology; hypoglycemia episodes trigger spasms (sympathetic surge → rigidity → falls). (4) Rituximab for SPS: depletes B cells producing anti-GAD65; may take months; IVIG as bridge. (5) Falls prevention: rigidity + osteoporosis (T −3.2) → fracture cycle; physical therapy + assistive devices; but spasms are unpredictable. (6) Respiratory compromise: chest wall rigidity during spasms can cause hypoventilation; if severe → ICU with benzodiazepine drip; intubation may be needed for refractory spasms. (7) Epilepsy: anti-GAD cerebellar/temporal involvement; levetiracetam preferred (benzodiazepines already given for SPS; avoid phenytoin). (8) Psychological: agoraphobia from fear of public spasms; functional impairment severe; quality of life counseling.

## CASE 39

### Acquired Hemophilia A with Life-Threatening Hemorrhage, Mechanical Valve, and Occult Malignancy

***Patient:*** *70 y/o F, BMI 24.2, Caucasian | **Disciplines:** Hematology, CT Surgery, Oncology, CC, Rheumatology, Vascular Surgery, Pharmacy*

### Active Disease States (20+)

Acquired hemophilia A (FVIII inhibitor 24 BU, no prior hemophilia); life-threatening hemorrhage (massive retroperitoneal hematoma, R thigh compartment syndrome); mechanical aortic valve (requires anticoagulation); supratherapeutic INR 3.2; suspected paraneoplastic trigger (lung mass 3.8 cm + mediastinal LAD); anemia (Hgb 4.8); aPTT >180s (unmeasurable); CHF (EF 40%); CKD 3; T2DM; HTN; afib; depression; chronic pain; immobilization DVT risk

### Clinical Presentation

Found with progressive bruising, then acute R thigh swelling and back pain. CT: massive retroperitoneal hematoma + R thigh compartment syndrome. INR 3.2 on warfarin. aPTT >180s. Mixing study: does not correct (inhibitor). FVIII <1%, inhibitor titer 24 BU. Hgb 4.8. Separately, CT chest: suspicious 3.8 cm lung mass + mediastinal LAD (likely malignancy triggering acquired hemophilia). Mechanical aortic valve needs anticoagulation — but she is hemorrhaging.

### Key Laboratory / Imaging Data

FVIII <1%, inhibitor 24 BU, aPTT >180s, mixing study: no correction, INR 3.2, Hgb 4.8, Plt 210K, fibrinogen 280, CT: retroperitoneal hematoma + R thigh compartment, chest CT: 3.8 cm lung mass + LAD, Cr 1.8, BNP 2400, EF 40%

### Current Medications

Bypassing agents (rFVIIa 90 mcg/kg q2h OR FEIBA 75 U/kg q8-12h), IV vitamin K + 4F-PCC (INR reversal — but aPTT won’t correct), warfarin held, immunosuppression to eradicate inhibitor: prednisone 1 mg/kg + rituximab (or CYC), pRBC transfusion, surgical fasciotomy consultation (R thigh — but coagulopathy)

### Multidisciplinary Challenges

(1) Bypassing agents for hemorrhage: rFVIIa and FEIBA are the only hemostatic options; neither is monitored by standard labs; thrombotic risk in patient with mechanical valve (embolization risk). (2) Mechanical valve off anticoagulation: warfarin stopped → thrombotic risk; but patient is hemorrhaging → anticoagulation = death; bridge with nothing is the only option during acute hemorrhage. (3) Fasciotomy: compartment syndrome needs urgent decompression but surgery causes massive bleeding without hemostasis; rFVIIa must be given continuously during and after procedure. (4) Inhibitor eradication: immunosuppression (prednisone + rituximab or CYC) takes weeks-months; must survive hemorrhage first. (5) Occult malignancy: lung mass likely triggering paraneoplastic acquired hemophilia; tissue diagnosis needed but biopsy → bleeding; CT-guided biopsy with bypassing agent coverage. (6) Anticoagulation restart: once inhibitor eradicated AND hemorrhage resolved → must resume warfarin for mechanical valve; monitoring complicated by residual inhibitor. (7) rFVIIa + FEIBA cannot be used simultaneously (thrombotic risk); must alternate; no lab monitoring available for efficacy. (8) Long-term: if malignancy confirmed → treatment may also treat acquired hemophilia (paraneoplastic cause); chemotherapy + continued IS.

## Case 1: 30

**Type:** CASE 30

**File:** Library case #30 (pp. 31–31)

**Demographics:** 36

**Diagnoses:** D10 [D10], CKD, neuropathy, depression, ALL, chronic pain, diabetes mellitus, type 1 diabetes, chronic kidney disease, gastrointestinal

**Labs documented:** 12

**Medications documented:** 6

**Clinical Question**

Given the working diagnoses of D10 [D10], CKD, neuropathy, current therapy including morphine, fentanyl, daptomycin, what does the retrieved evidence indicate about management, risks, and appropriate next steps?

**Evidence Summary**

Total sources: 7 · Guidelines: 0 · SR/MA: 0 · RCTs: 0 · Other: 7

**Reference Outline**

**Applicable guidelines:**

• KDIGO — Kidney Disease (https://kdigo.org/guidelines/)

**Historical precedent:** Trajectory of understanding for D10 [D10] has shifted from symptom-based empirical management toward mechanism-targeted therapy informed by receptor-level, genetic, and molecular-pathway data. Recent decades have emphasized biomarker-guided dosing, shared decision-making, and cumulative-harm minimization across comorbid conditions.

**Current research findings:**

- Precision-medicine paradigm: genotype-guided dosing and pharmacogenomics are moving from research into routine practice for selected drug classes (e.g., CYP2C19 for clopidogrel, CYP2C9/VKORC1 for warfarin, HLA-B*5701 for abacavir).
- Digital twin and closed-loop control systems: dynamic adaptive therapy models use real-time physiological inputs to titrate dosing, an active area of investigation in ICU sedation, insulin delivery, and anticoagulation.
- Root-cause cascades: increasing emphasis on mechanistic polyroot analysis in quality improvement (RCA2) and pharmacovigilance (FMEA) to preempt iatrogenic harm rather than react to it.

**Reasoning Chain**

1. Case profile: 36 patient with working diagnoses of D10 [D10], CKD, neuropathy · 12 lab value(s), 6 medication(s) documented
2. Case-derived findings: 1 pattern identified (1 moderate/mild)
3. Evidence base: spans 1910–2024; 0 distinct methodology type(s) · 0 evidence-linked statement(s) extracted
4. Case findings did not align directly with retrieved evidence — external verification required
5. 1 finding(s) flagged for verification: evidence retrieved did not directly address these elements. Recommendations based on general clinical principles until verified against specialty resources.
6. Overall: evidence base insufficient for definitive inference; treat output as hypothesis-generating pending verification

**50-Why Polyroot Inference Chain**

**Seed:** Hyponatremia (Na 128 mEq/L) · **Terminal:** p53 phosphorylation (ATM/ATR/CHK1/CHK2) · **Iterations:**

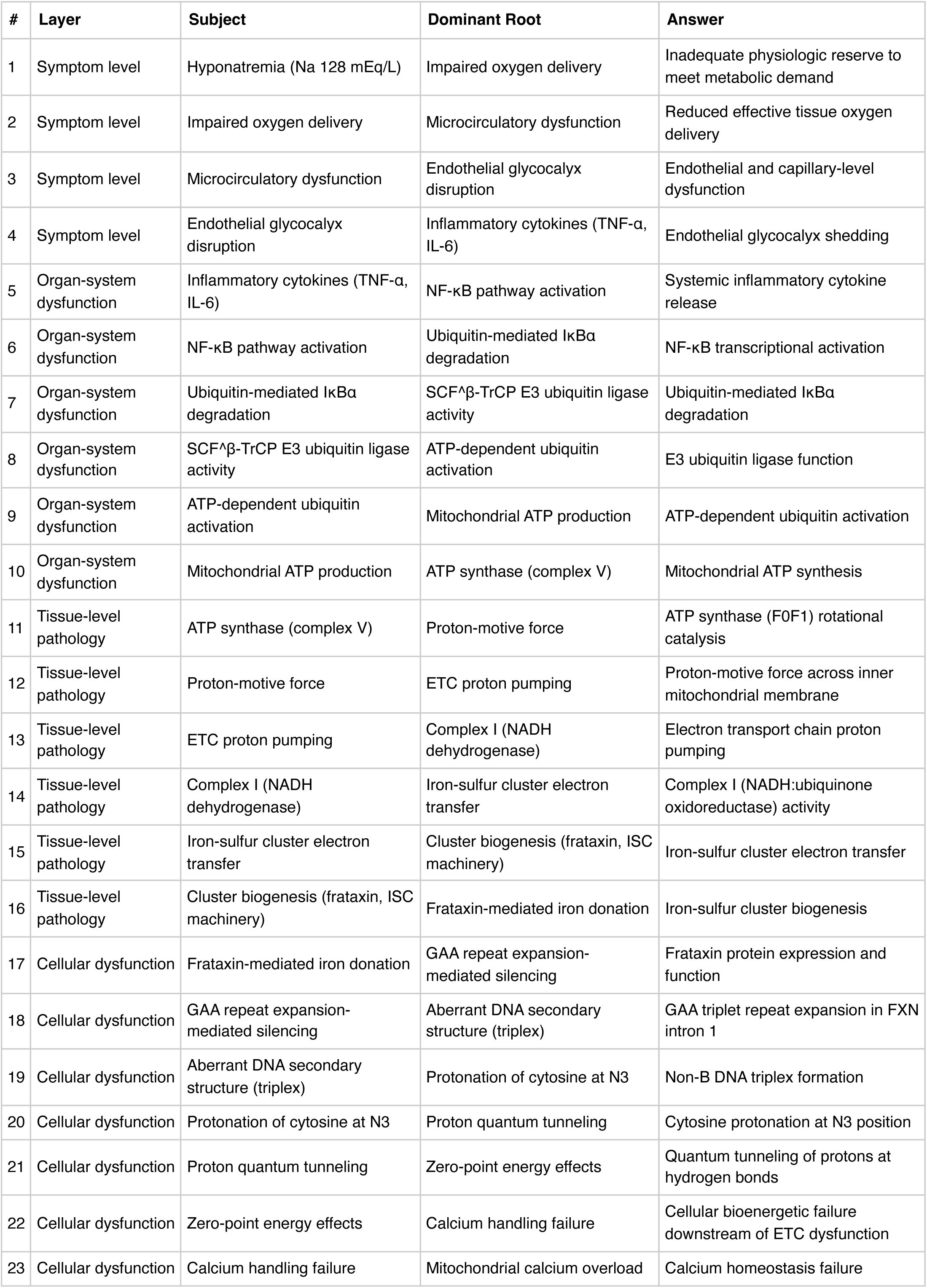
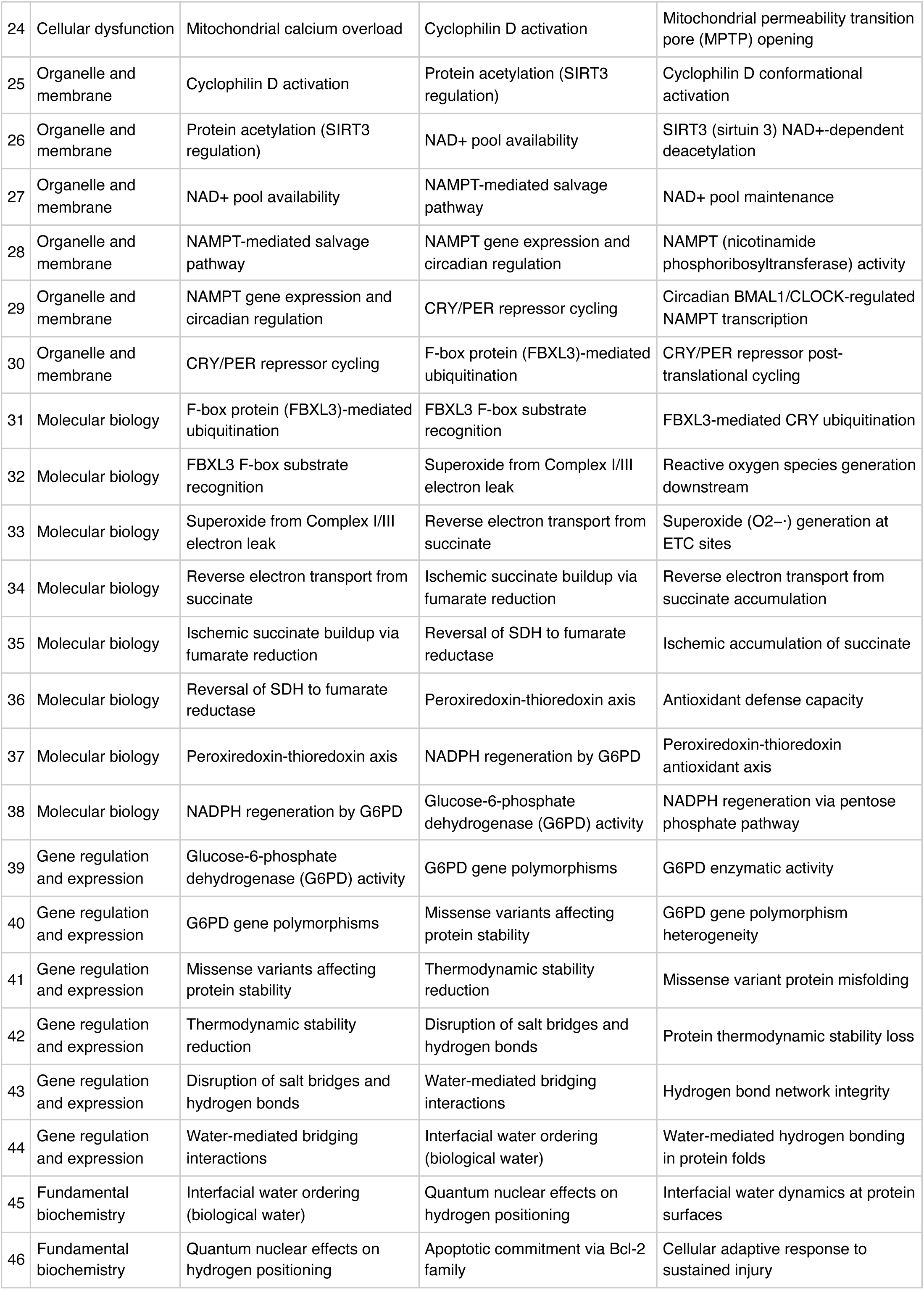
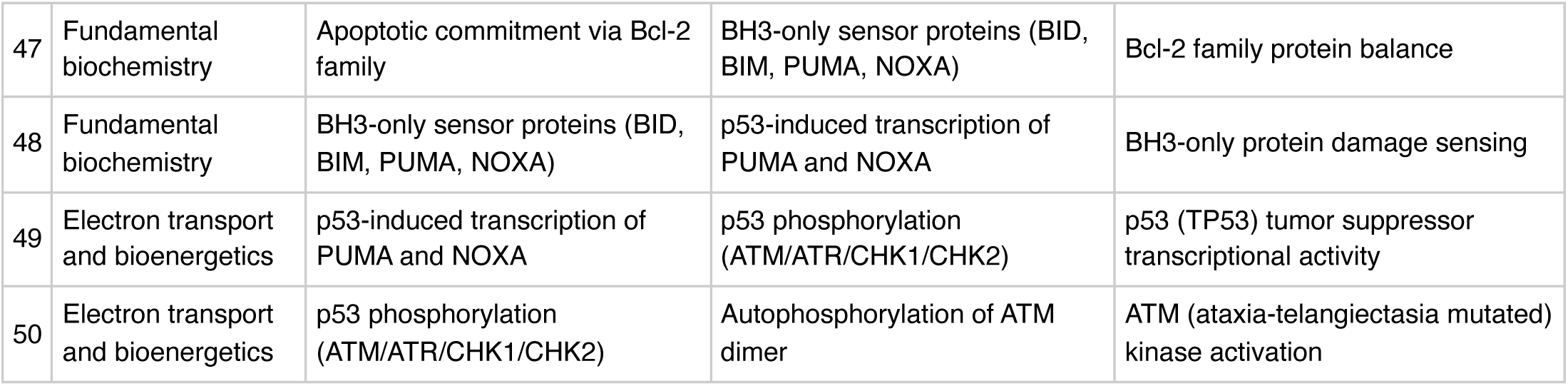

**FMEA Forecasting (Severity × Probability × Detectability)**

**Terminal node:** ATM (ataxia-telangiectasia mutated) kinase activation

FMEA Forecasting (Severity × Probability × Detectability) **Terminal node:** ATM (ataxia-telangiectasia mutated) kinase activation
| Domain | Score | Justification |
| --- | --- | --- |
| Severity | 4/10 | Moderate clinical severity based on findings profile. |
| Probability | 3/10 | Probability estimated from concurrent-finding count (1). A higher count indicates multiple parallel activations of the cascade, increasing the likelihood that this specific 50-step pathogenic chain is operative in this patient. |
| Detectability | 10/10 | Terminal root node is at the quantum level. Current clinical diagnostics do not routinely detect pathology at this level before upstream clinical events manifest. |

**RPN 120 (LOW):** RPN <150 — standard guideline-directed therapy addresses this cascade adequately; deep-root intervention is not clinically prioritized.

**Therapy Benchmarks**

**Current GDMT:** RAAS inhibition (ACEi/ARB), SGLT2 inhibitor, nonsteroidal MRA (finerenone in CKD-T2DM), avoidance of nephrotoxins, guideline-based BP control (KDIGO 2024)

**Mechanism gap:** Current GDMT slows progression by reducing glomerular hyperfiltration and inflammation but does not reverse tubular epithelial ATP depletion, iron-sulfur-cluster biogenesis defects, or fibrosis once established.

**Deep-root intervention (theoretical):** A hypothetical intervention targeting the terminal root (ATM (ataxia-telangiectasia mutated) kinase activation) would act upstream of the entire 50-step cascade, theoretically preventing the downstream pathophysiology addressed by current GDMT.

**Therapeutic delta:** Deep-root interventions, if tractable, would theoretically produce NNT reductions greater than any single pillar of current GDMT because they act upstream of multiple downstream manifestations.

However, most root-node targets at the molecular or genetic level are not currently druggable, so the theoretical benefit cannot yet be realized in clinical practice.

**Guideline alignment:** Continue guideline-directed medical therapy as the foundation of management. Use the polyroot analysis as educational scaffolding for mechanism-based decisions within GDMT (e.g., choosing between agents that act at different levels of the cascade), not as a substitute for GDMT.

**Fishbone (Ishikawa) Diagram**

**Effect:** Hyponatremia (Na 128 mEq/L)

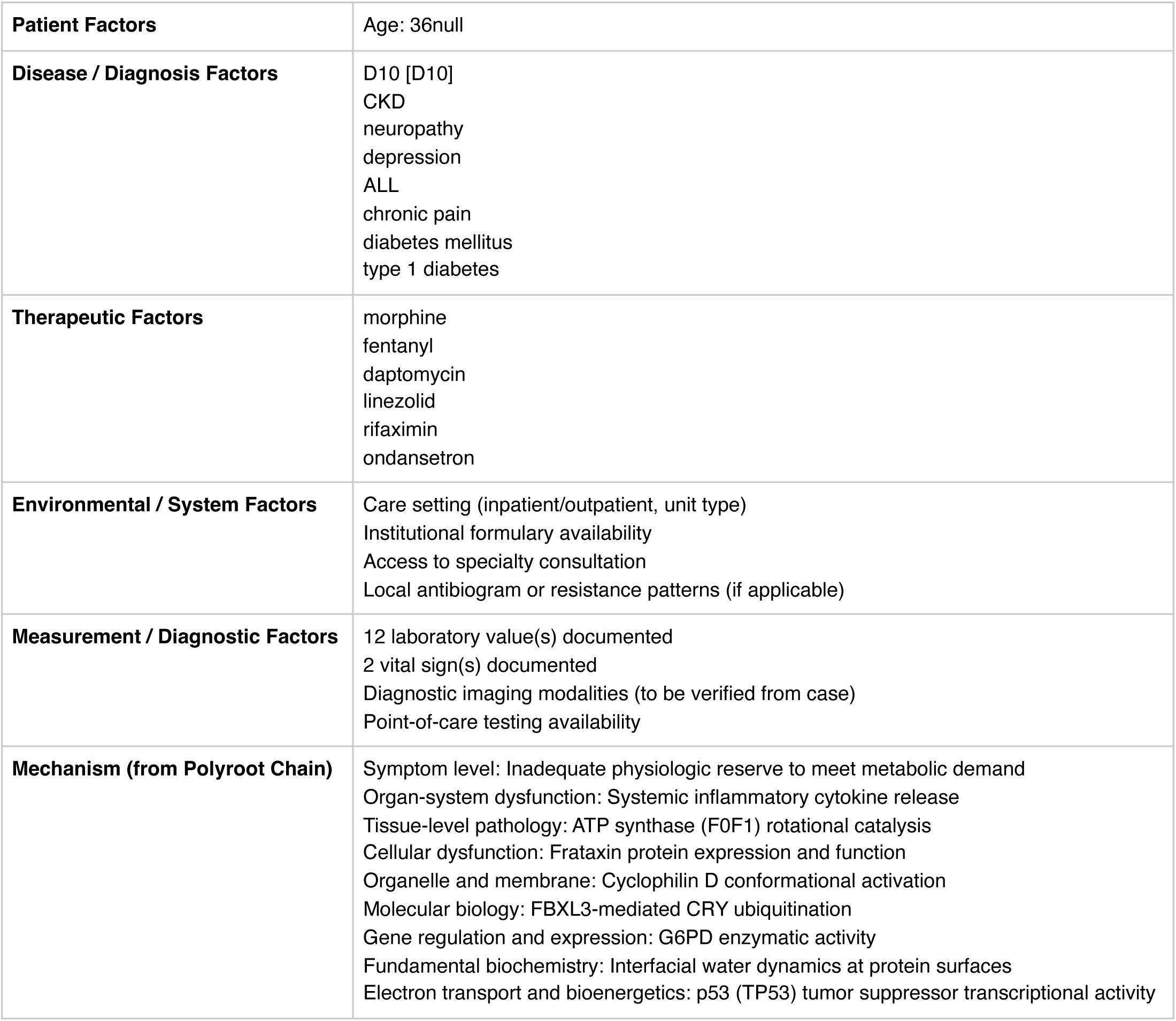

**Run Chart — Actual vs Guideline Targets**

| Metric | Actual | Target | Status | Source |
| --- | --- | --- | --- | --- |
| Serum sodium | 128 mEq/L | 135–145 | below target | Routine chemistry panel reference |
| Plasma glucose | 68 mg/dL | 70–180 | below target | ADA inpatient glycemic target |
| Hemoglobin A1c | 10.4 % | —–7 | above target | ADA general adult target |
| INR | 1.8 | —–3 | within target | Therapeutic range for most anticoagulation indications |
| ALT | 98 U/L | —–40 | above target | Hepatocellular injury threshold |

**Pharmacogenomics Screen**

No medications in this case mapped to the embedded PGx registry. This does not exclude pharmacogenomic relevance — consult PharmGKB directly for medications not in the registry.

**Drug-Drug Interaction Screen**

No interactions detected in the embedded DDI matrix among the 6 documented medication(s). This does not exclude clinically significant interactions — verification via Lexi-Interact or Drugs.com Interaction Checker is still recommended for complex regimens.

**Clinical Visualizations — Isobolograms & Nomograms**

**Serotonin Syndrome Additive-Risk Isobologram**

3 serotonergic agents: fentanyl, linezolid, ondansetron

**Serotonergic Combination Isobologram**

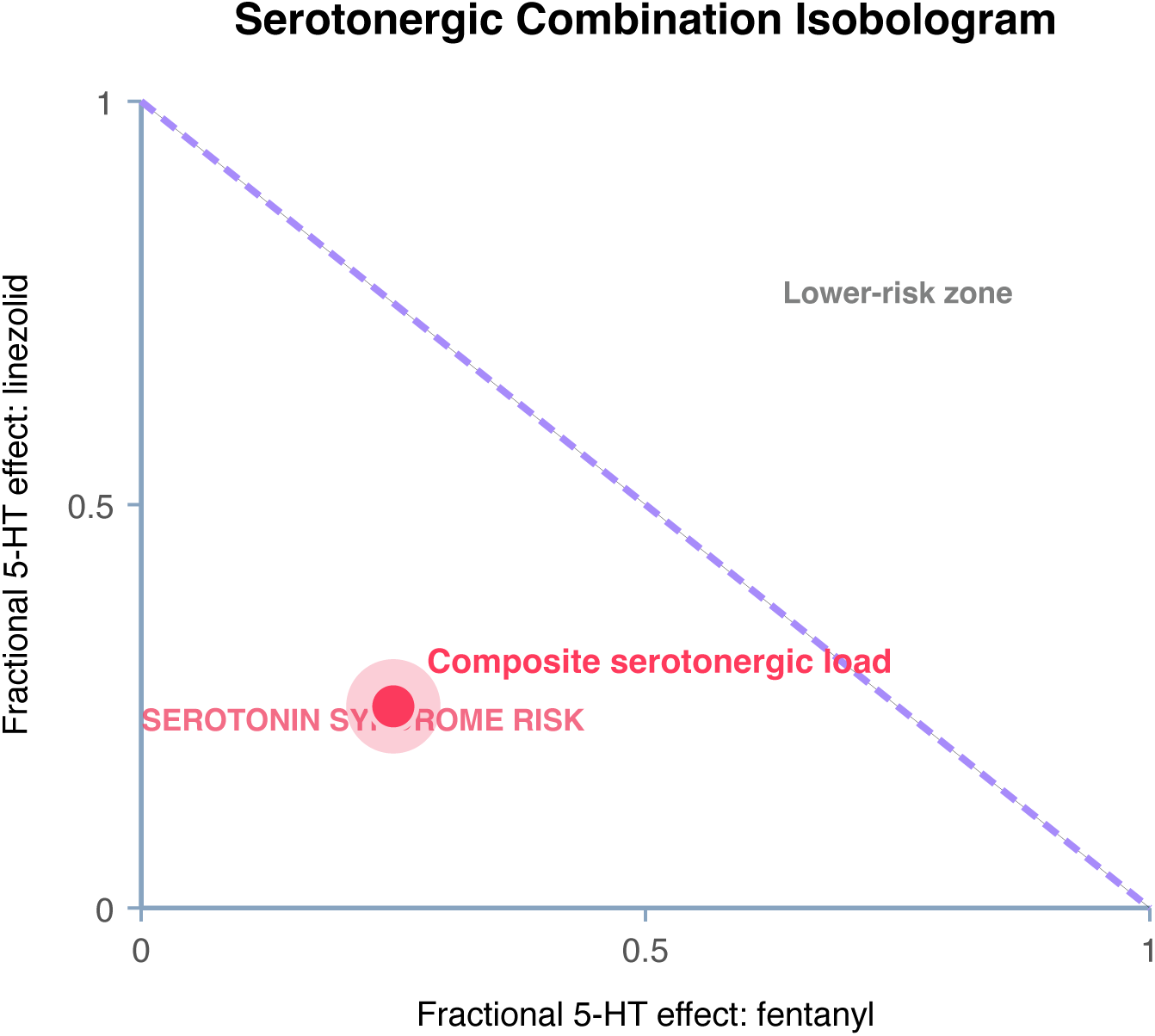

**Interpretation:** MAO inhibitor or linezolid combined with another serotonergic agent — highest-severity combination. Serotonin syndrome risk is clinically significant and requires immediate review.

**Clinical action:** URGENT: hold the serotonergic agent; MAO-I washout period is 14 days (5 weeks for fluoxetine). Monitor for hyperthermia, clonus, tremor, hyperreflexia. Cyproheptadine is antidote for severe cases.

**CNS Depressant Synergy Isobologram**

2 sedating/respiratory-depressant agents: morphine, fentanyl

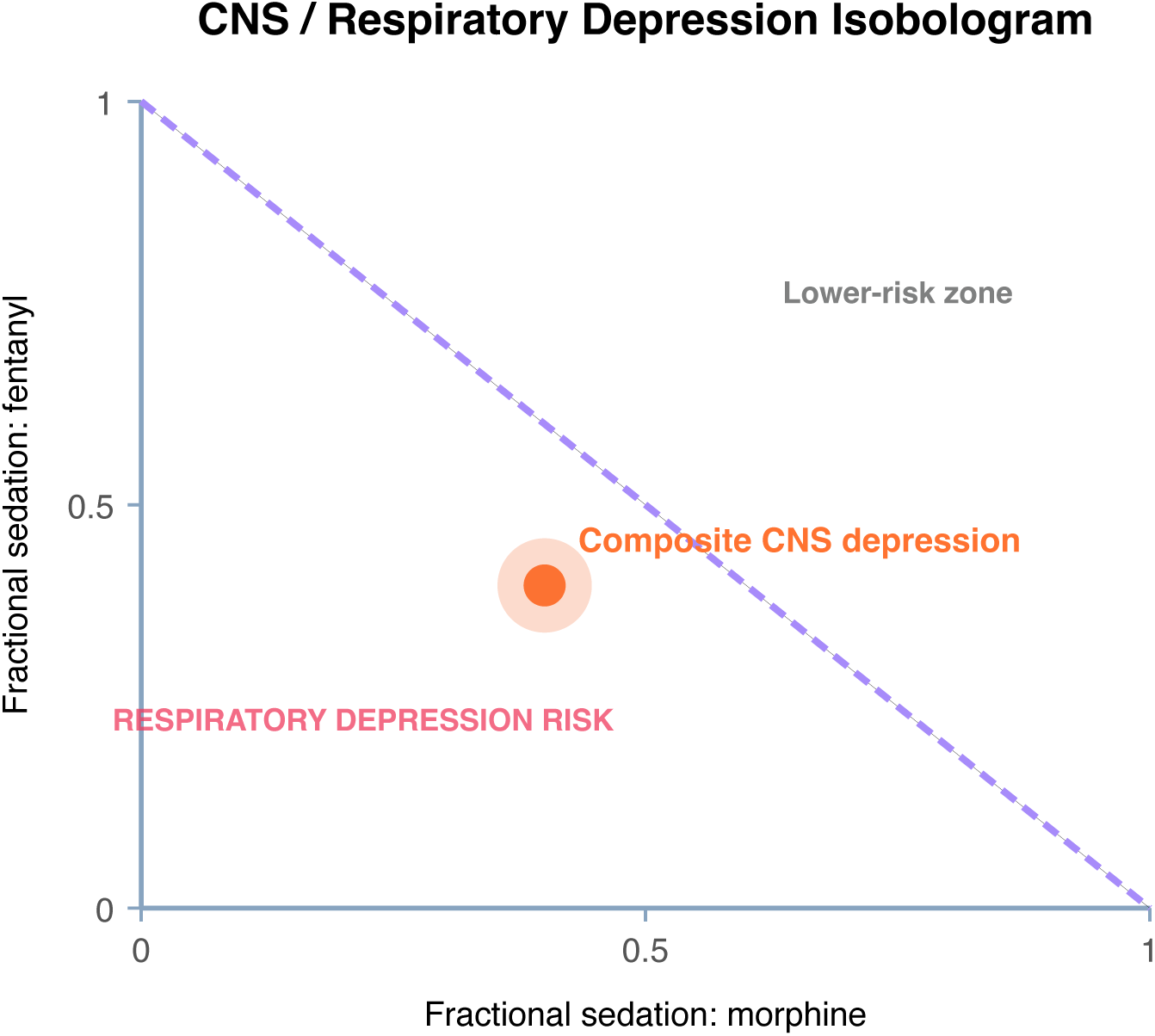

**Interpretation:** Two CNS depressants produce additive sedation; respiratory depression risk is clinically meaningful in opioid-naive patients, elderly, and those with OSA or pulmonary disease.

**Clinical action:** Minimize simultaneous dosing. Consider naloxone co-prescription if cumulative MME ≥50. Monitor for excessive sedation, particularly first 24-72 hours.

**Wells DVT Nomogram (structured-data subset)**

Score 0 (Low probability)

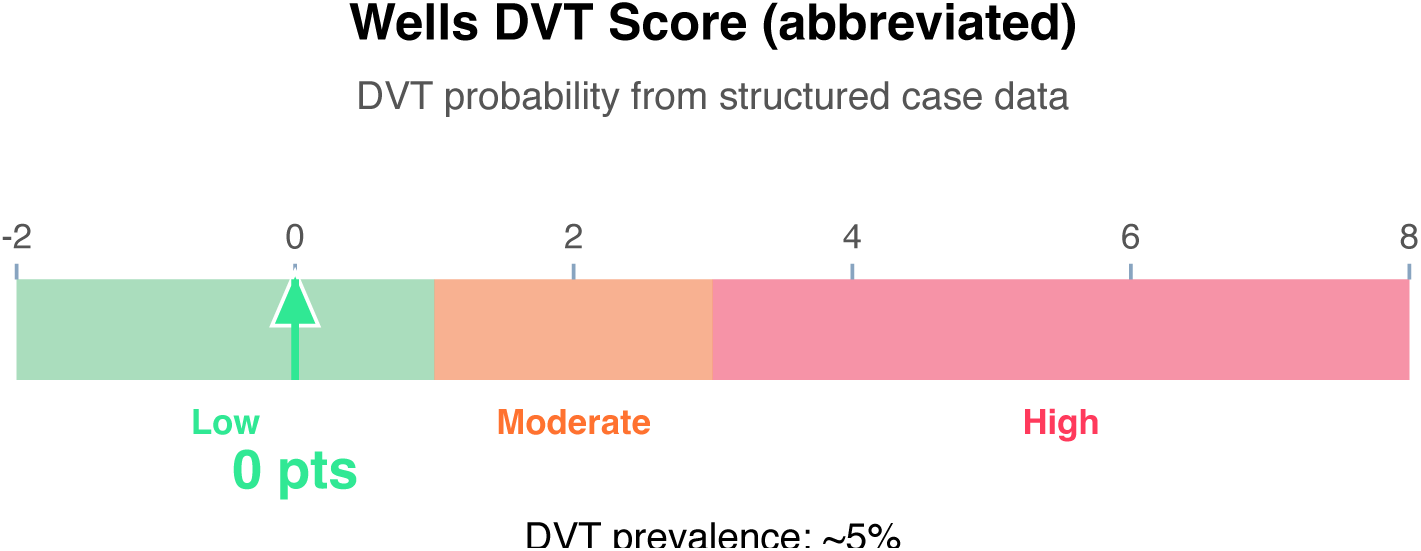

**Interpretation:** Score reflects only factors extractable from structured case data. Full Wells DVT adds examination findings (localized tenderness, entire-leg swelling, calf asymmetry >3 cm, pitting edema, collateral veins) and “alternative diagnosis as likely or more likely” modifier (−2).

**Clinical action:** D-dimer; if negative, DVT effectively ruled out.

**Morphine Equivalent Daily Dose Nomogram**

MME risk thresholds (CDC 2022)

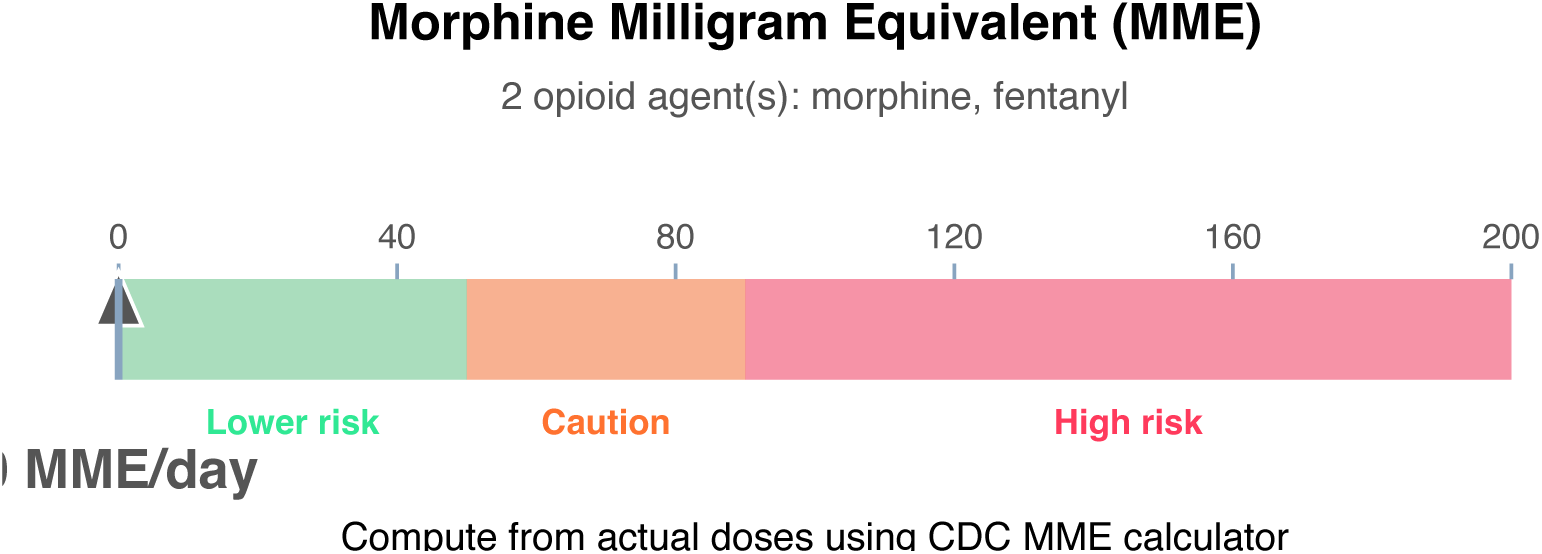

**Interpretation:** Cumulative MME/day thresholds per CDC 2022: <50 MME = lower risk; 50-89 MME = increased risk (use caution, document rationale); ≥90 MME = markedly elevated overdose risk (avoid or offer naloxone, taper if appropriate). Structured case data does not contain reliable daily-dose totals for computation.

**Clinical action:** Compute patient-specific MME using the CDC calculator. Prescribe naloxone if MME ≥50 or other risk factors (concurrent benzodiazepine, respiratory disease, age ≥65, OUD history).

**Article Structure (iteratively restructured)**

**Structure type:** narrative_review · **Converged:** true at pass 2/5 · **Quality:** 0.05

*Source mix is primarily narrative/review literature (0% review-type sources). Narrative review format is appropriate*.

**Proposed outline:**

1. **Abstract** (0 clusters, 0 sources) Overview of the review scope and synthesis
2. **Background** (0 clusters, 0 sources) Historical context and evolution of understanding
3. **Current State of Evidence** (0 clusters, 0 sources) What is established vs uncertain
4. **Controversies and Debates** (0 clusters, 0 sources) Contradictory findings and interpretive disagreements
5. **Clinical Implications** (0 clusters, 0 sources) What the evidence means for practice
6. **Future Directions** (0 clusters, 0 sources) Knowledge gaps and research agenda
7. **Conclusion** (0 clusters, 0 sources) Synthesis in one paragraph
8. **References** (0 clusters, 7 sources) Complete citation list

**Structural gaps:**

- [MODERATE] empty section (Background) — No clusters or sources mapped to this section. Consider broadening the search to include review articles and mechanistic/pathophysiologic literature.
- [MODERATE] empty section (Current State of Evidence) — No clusters or sources mapped to this section. Consider broadening the search to include targeted supplementary queries.
- [MODERATE] empty section (Controversies and Debates) — No clusters or sources mapped to this section. Consider broadening the search to include targeted supplementary queries.
- [MODERATE] empty section (Clinical Implications) — No clusters or sources mapped to this section. Consider broadening the search to include clinical practice guidelines.
- [MODERATE] empty section (Future Directions) — No clusters or sources mapped to this section. Consider broadening the search to include targeted supplementary queries.
- [MODERATE] empty section (Conclusion) — No clusters or sources mapped to this section. Consider broadening the search to include targeted supplementary queries.
- [MODERATE] no quantitative data — No quantitative statements extracted (p-values, HRs, ORs, CIs). Consider searching PubMed for RCTs or meta-analyses on this topic.

**Quantitative Evidence Extraction**

**Cross-source summary:** Pooled N: n/a · 0/7 with p-values · 0/7 with CIs · 0/7 with effect sizes · 0 significant · 0 null

**GRADE Methodological Assessment**

| Domain | Concern | Rationale |
| --- | --- | --- |
| Risk of Bias | HIGH | Majority of sources show high risk of bias (incomplete methodological reporting or no adjustment for confounders). |
| Inconsistency | MODERATE | No heterogeneity statistics ($I^2$, $Q$, $\tau^2$) detected in retrieved text. Unable to formally assess inconsistency across 7 sources. |
| Indirectness | HIGH | Low alignment — only 29% of sources reference case diagnoses. Retrieved evidence may be substantially indirect. |
| Imprecision | LOW | Sample size, CI widths, and event counts do not raise precision concerns. |
| Publication Bias | HIGH | No funnel plot or Egger's/Begg's test detected in 7 sources; No trial registry or pre-registration verification detected. |

**Inferential Findings**

& Hyponatremia (Na 128 mEq/L) [LIMITED CONFIDENCE]

→ Evaluate volume status (SIADH vs hypovolemic vs hypervolemic); medication review

⚠ Retrieved evidence base did not address this finding directly — independent verification through Lexicomp, UpToDate, or specialty-specific guidelines recommended

**Recommended Verification Steps**

1. **[ROUTINE]** Verify management approach via specialty-specific resources — retrieved evidence base did not directly address this finding *Rationale:* Hyponatremia (Na 128 mEq/L) *Resources:* UpToDate, DynaMed, Specialty society guidelines, Lexicomp/Micromedex, DailyMed for current product labeling
2. **[ROUTINE]** Comprehensive medication reconciliation and interaction review for 6 agents *Rationale:* Polypharmacy (≥3 agents) warrants systematic DDI and appropriateness review *Resources:* Drugs.com Interaction Checker, Lexi-Interact, Beers Criteria (if age ≥65), STOPP/START criteria
3. **[ROUTINE]** Cross-verify all recommendations against institutional protocols, current product labeling (DailyMed), and specialty-specific guidelines before clinical application *Rationale:* AuditMed outputs are for research and educational use; clinical decisions remain the responsibility of the treating clinician *Resources:* Institutional clinical practice guidelines, DailyMed FDA labeling, Specialty society consensus documents

**Evidence-Based Recommendations**

- Retrieved evidence does not contain explicit recommendation-style statements. Proceed with established standard-of-care for D10 [D10], CKD, neuropathy, depression, ALL, chronic pain, diabetes mellitus, type 1 diabetes, chronic kidney disease, gastrointestinal while consulting specialty-specific guidelines (IDSA, ACC/AHA, ADA, KDIGO, or others as applicable). **[LIMITED]**
- Verify all therapeutic changes against current product labeling (DailyMed), institutional formulary, and patient-specific factors (allergies, organ function, goals of care) before implementation. **[N/A]**

**Strength of Evidence**

**GRADE:** LOW

*7 source(s) retrieved · 1 limited-confidence finding(s) flagged for verification*

## Case 1: 31

**Type:** CASE 31

**File:** Library case #31 (pp. 32–32)

**Demographics:** 48

**Diagnoses:** heart block, CKD, depression, lupus, osteoporosis, hypercalcemia, obesity, diabetes mellitus, chronic kidney disease

**Labs documented:** 3

**Medications documented:** 4

**Clinical Question**

Given the working diagnoses of heart block, CKD, depression, current therapy including amiodarone, hydrocortisone, infliximab, what does the retrieved evidence indicate about management, risks, and appropriate next steps?

**Evidence Summary**

Total sources: 10 · Guidelines: 0 · SR/MA: 0 · RCTs: 0 · Other: 10

**Reference Outline**

**Applicable guidelines:**

- KDIGO — Kidney Disease (https://kdigo.org/guidelines/)
- APA — Psychiatry (https://www.psychiatry.org/psychiatrists/practice/clinical-practice-guidelines)

**Historical precedent:** Trajectory of understanding for heart block has shifted from symptom-based empirical management toward mechanism-targeted therapy informed by receptor-level, genetic, and molecular-pathway data. Recent decades have emphasized biomarker-guided dosing, shared decision-making, and cumulative-harm minimization across comorbid conditions.

**Current research findings:**

**Reasoning Chain**

1. Case profile: 48 patient with working diagnoses of heart block, CKD, depression · 3 lab value(s), 4 medication(s) documented
2. Case-derived findings: 1 pattern identified (1 moderate/mild)
3. Evidence base: spans 1947–2025; 1 distinct methodology type(s) · 0 evidence-linked statement(s) extracted
4. Case findings did not align directly with retrieved evidence — external verification required
5. 1 finding(s) flagged for verification: evidence retrieved did not directly address these elements. Recommendations based on general clinical principles until verified against specialty resources.
6. Overall: evidence base insufficient for definitive inference; treat output as hypothesis-generating pending verification

**50-Why Polyroot Inference Chain**

**Seed:** Fever (48°C) · **Terminal:** p53 phosphorylation (ATM/ATR/CHK1/CHK2) · **Iterations:** 50

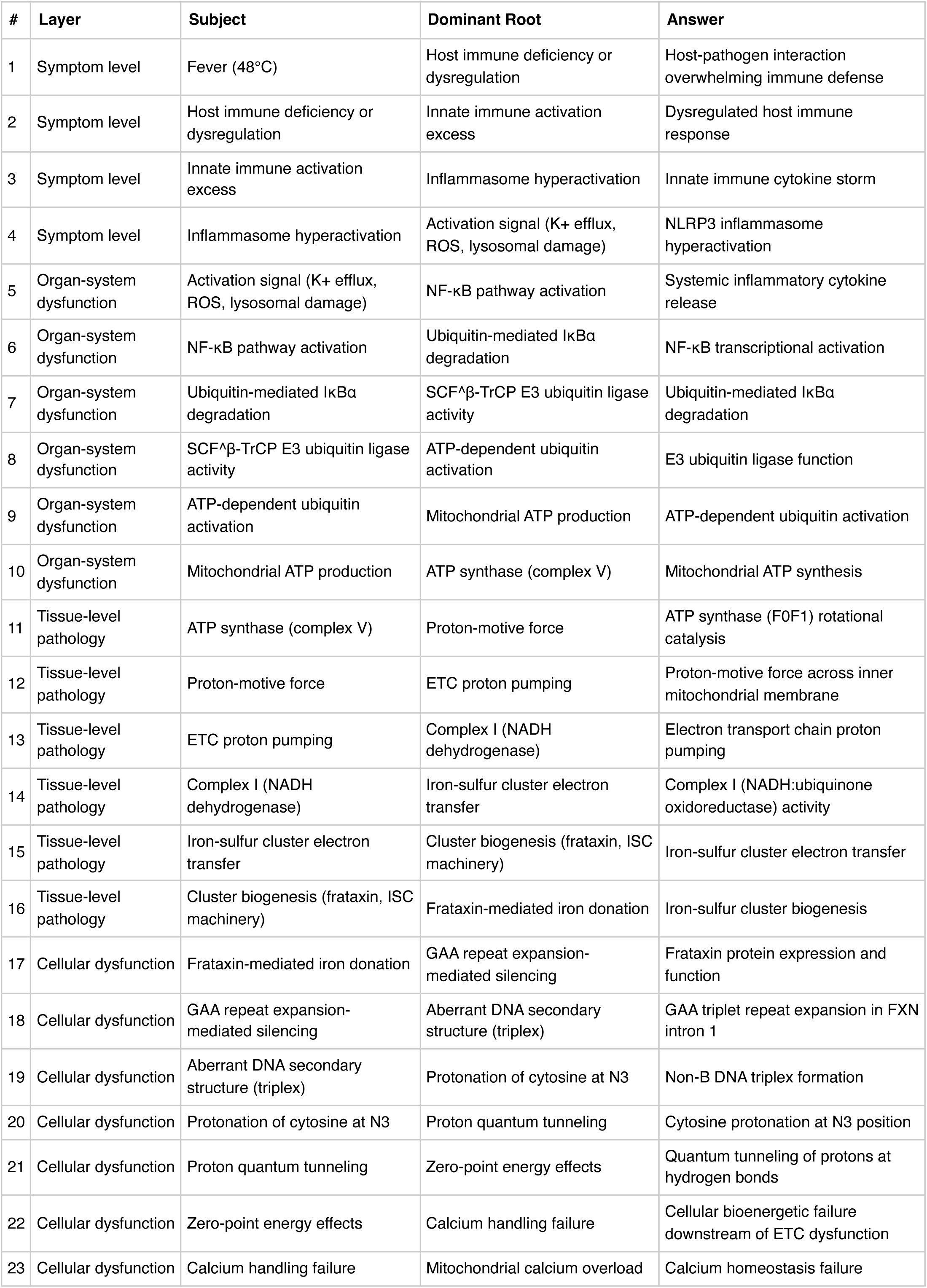
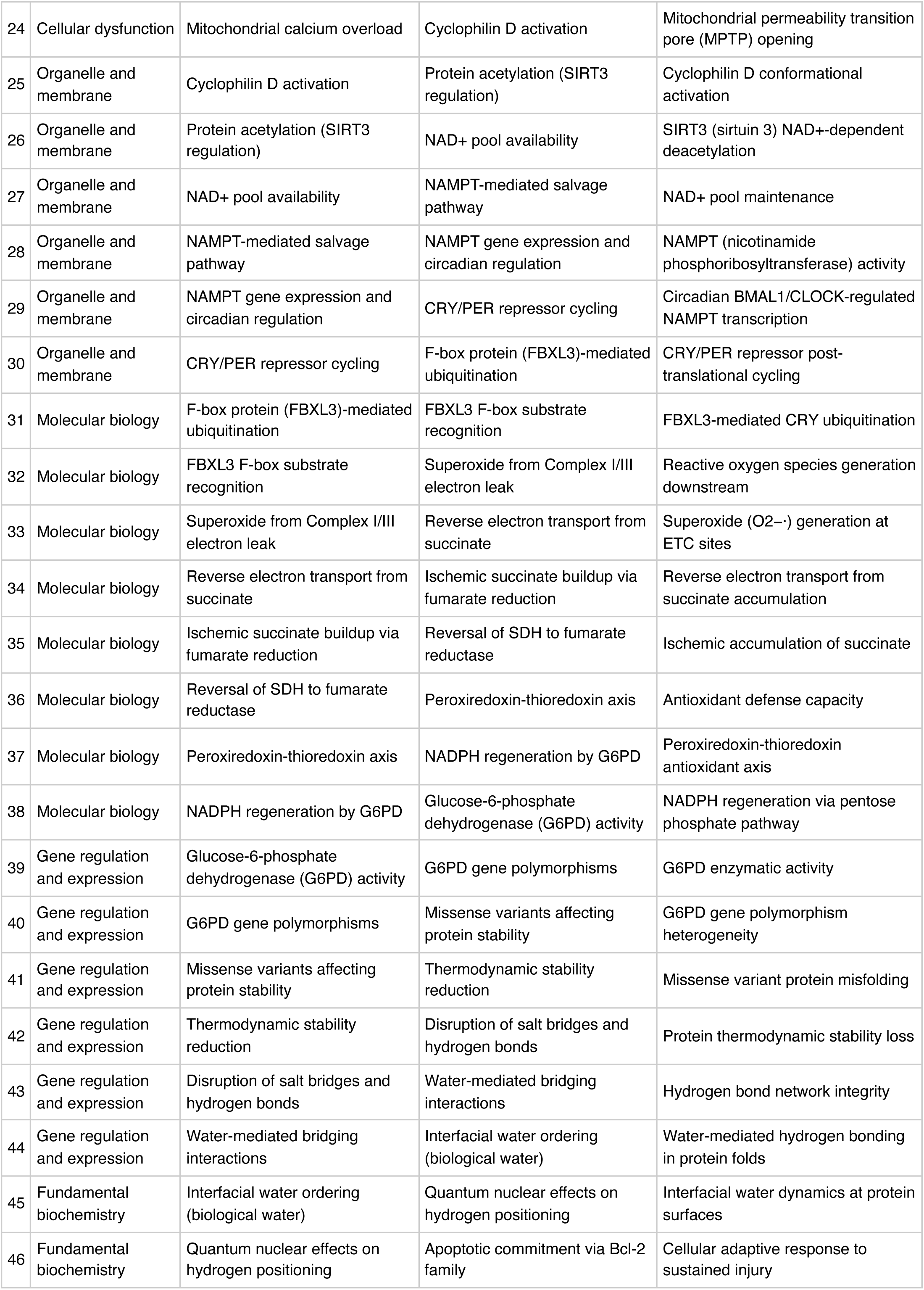
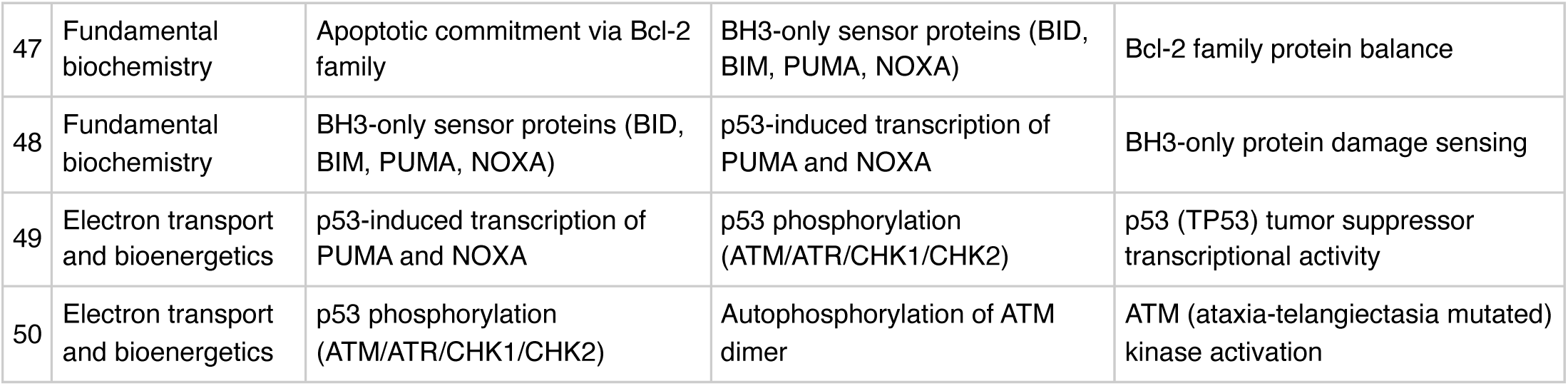

**FMEA Forecasting (Severity × Probability × Detectability)**

**Terminal node:** ATM (ataxia-telangiectasia mutated) kinase activation

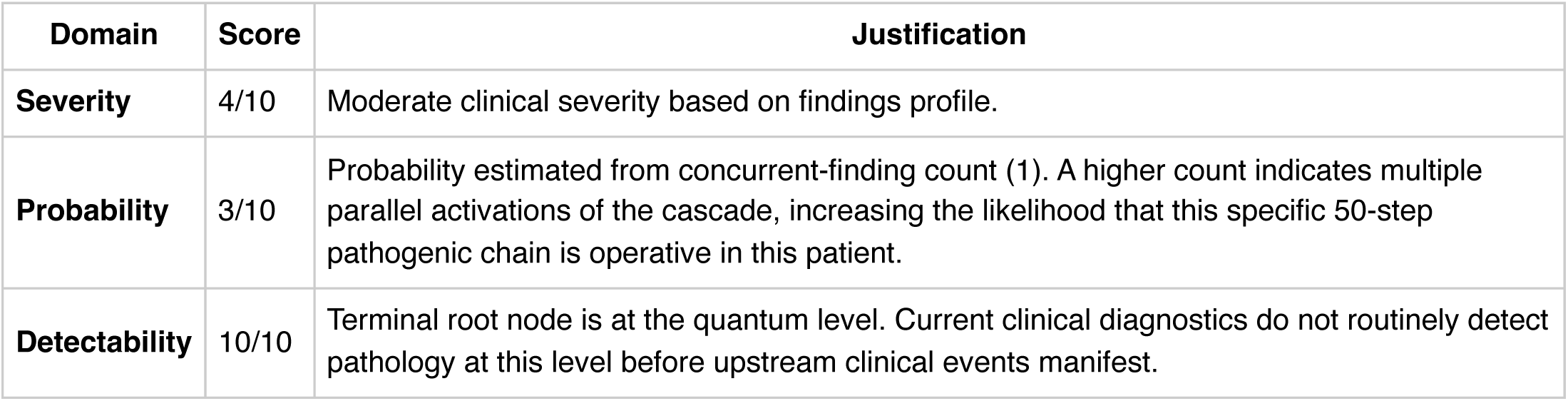

**Therapy Benchmarks**

**Fishbone (Ishikawa) Diagram**

**Effect:** Fever (48°C)

|  |  |
| --- | --- |
| **Patient Factors** | Age: 48null |
| **Disease / Diagnosis Factors** | heart block<br>CKD<br>depression<br>lupus<br>osteoporosis<br>hypercalcemia<br>obesity<br>diabetes mellitus |
| **Therapeutic Factors** | amiodarone<br>hydrocortisone<br>infliximab<br>zoledronic acid |
| **Environmental / System Factors** | Care setting (inpatient/outpatient, unit type)<br>Institutional formulary availability<br>Access to specialty consultation<br>Local antibiogram or resistance patterns (if applicable) |
| **Measurement / Diagnostic Factors** | 3 laboratory value(s) documented<br>2 vital sign(s) documented<br>Diagnostic imaging modalities (to be verified from case)<br>Point-of-care testing availability |
| **Mechanism (from Polyroot Chain)** | Symptom level: Host-pathogen interaction overwhelming immune defense<br>Organ-system dysfunction: Systemic inflammatory cytokine release<br>Tissue-level pathology: ATP synthase (F0F1) rotational catalysis<br>Cellular dysfunction: Frataxin protein expression and function<br>Organelle and membrane: Cyclophilin D conformational activation<br>Molecular biology: FBXL3-mediated CRY ubiquitination<br>Gene regulation and expression: G6PD enzymatic activity<br>Fundamental biochemistry: Interfacial water dynamics at protein surfaces<br>Electron transport and bioenergetics: p53 (TP53) tumor suppressor transcriptional activity |

**Run Chart — Actual vs Guideline Targets**

| Metric | Actual | Target | Status | Source |
| --- | --- | --- | --- | --- |
| Plasma glucose | 248 mg/dL | 70–180 | above target | ADA inpatient glycemic target |
| Hemoglobin | 10.4 g/dL | 12— | below target | WHO anemia cut-offs |

**Pharmacogenomics Screen**

**Drug-Drug Interaction Screen**

No interactions detected in the embedded DDI matrix among the 4 documented medication(s). This does not exclude clinically significant interactions — verification via Lexi-Interact or Drugs.com Interaction Checker is still recommended for complex regimens.

**Clinical Visualizations — Isobolograms & Nomograms**

**Hepatotoxicity Combination Isobologram**

2 hepatotoxic agents: amiodarone, infliximab

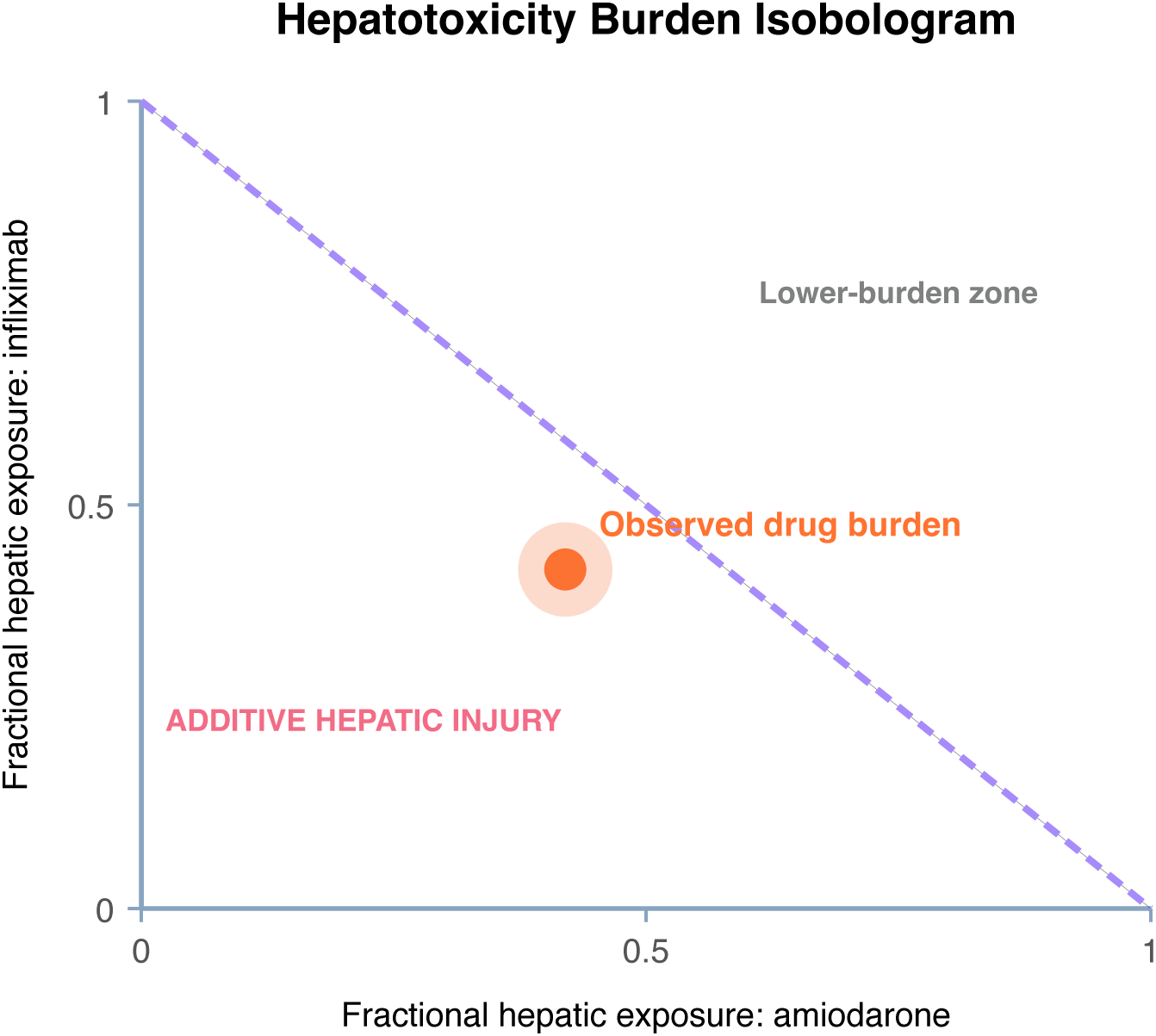

**Interpretation:** Multiple hepatotoxic agents in regimen. Pre-clinical injury may be absent, but synergistic hepatic stress is plausible — particularly relevant for acetaminophen + chronic alcohol, isoniazid + rifampin, or methotrexate + other DILI agents.

**Clinical action:** Check LFTs at baseline and every 1–2 weeks initially. If ALT rises >3× ULN or bilirubin >2× ULN (Hy’s law threshold), discontinue the least-essential hepatotoxin. LiverTox database for agent-specific patterns.

**qSOFA Sepsis Screen**

Score 0 (mentation requires bedside assessment)

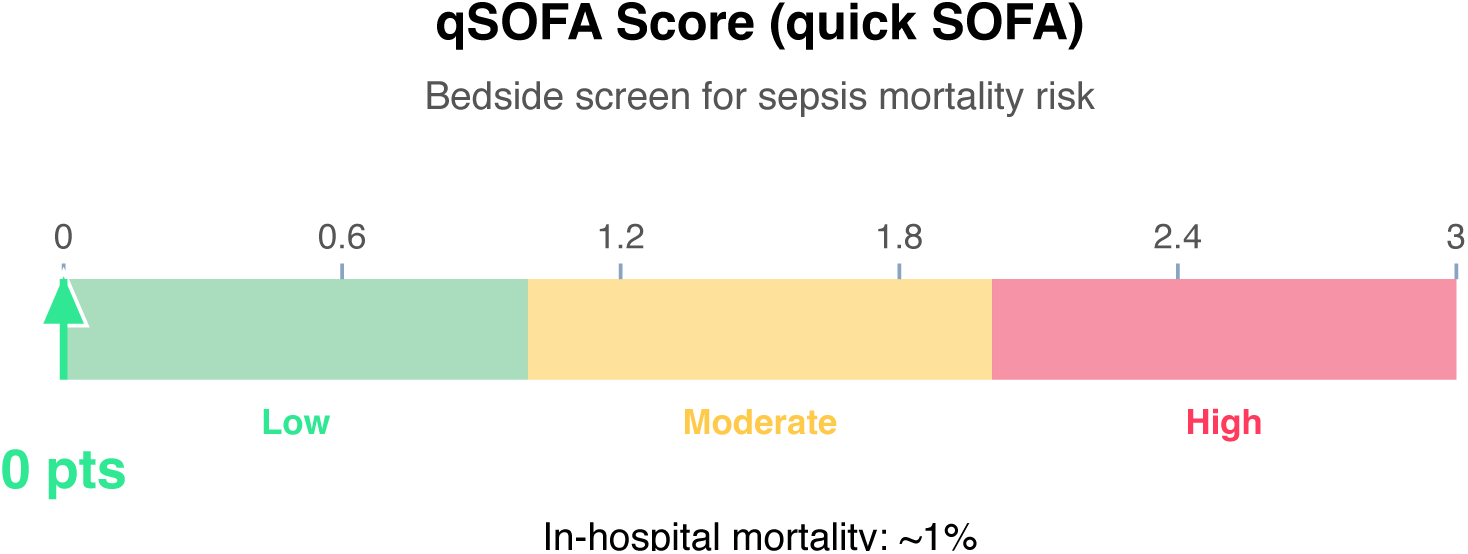

**Interpretation:** qSOFA ≥2 in suspected infection flags high risk of poor outcome. Note: qSOFA is a bedside screen, not diagnostic. Current structured case data scores 0; add 1 point if altered mental status is present.

**Clinical action:** Continue monitoring; repeat if clinical deterioration.

**Article Structure (iteratively restructured)**

**Structure type:** narrative_review · **Converged:** true at pass 2/5 · **Quality:** 0.05

**Proposed outline:**

1. **Abstract** (0 clusters, 0 sources) Overview of the review scope and synthesis
2. **Background** (0 clusters, 0 sources) Historical context and evolution of understanding
3. **Current State of Evidence** (0 clusters, 0 sources) What is established vs uncertain
4. **Controversies and Debates** (0 clusters, 0 sources) Contradictory findings and interpretive disagreements
5. **Clinical Implications** (0 clusters, 0 sources) What the evidence means for practice
6. **Future Directions** (0 clusters, 0 sources) Knowledge gaps and research agenda
7. **Conclusion** (0 clusters, 0 sources) Synthesis in one paragraph
8. **References** (0 clusters, 10 sources) Complete citation list

**Structural gaps:**

**Quantitative Evidence Extraction**

**Cross-source summary:** Pooled N: n/a · 0/10 with p-values · 0/10 with CIs · 0/10 with effect sizes · 0 significant · 0 null

**GRADE Methodological Assessment**

| Domain | Concern | Rationale |
| --- | --- | --- |
| Risk of Bias | HIGH | Majority of sources show high risk of bias (incomplete methodological reporting or no adjustment for confounders). |
| Inconsistency | MODERATE | No heterogeneity statistics ($I^2$, Q, $\tau^2$) detected in retrieved text. Unable to formally assess inconsistency across 10 sources. |
| Indirectness | LOW | Most retrieved sources (100%) reference the case diagnoses directly. Population and condition appear well-aligned. |
| Imprecision | LOW | Sample size, CI widths, and event counts do not raise precision concerns. |
| Publication Bias | HIGH | No funnel plot or Egger's/Begg's test detected in 10 sources; No trial registry or pre-registration verification detected. |

**Inferential Findings**

& **Fever (48°C) [LIMITED CONFIDENCE]**

→ Infection workup — blood cultures, imaging, source identification

**Recommended Verification Steps**

1. **[ROUTINE]** Verify management approach via specialty-specific resources — retrieved evidence base did not directly address this finding *Rationale:* Fever (48°C) *Resources:* UpToDate, DynaMed, Specialty society guidelines, Lexicomp/Micromedex, DailyMed for current product labeling
2. **[ROUTINE]** Comprehensive medication reconciliation and interaction review for 4 agents *Rationale:* Polypharmacy (≥3 agents) warrants systematic DDI and appropriateness review *Resources:* Drugs.com Interaction Checker, Lexi-Interact, Beers Criteria (if age ≥65), STOPP/START criteria
3. **[ROUTINE]** Cross-verify all recommendations against institutional protocols, current product labeling (DailyMed), and specialty-specific guidelines before clinical application *Rationale:* AuditMed outputs are for research and educational use; clinical decisions remain the responsibility of the treating clinician *Resources:* Institutional clinical practice guidelines, DailyMed FDA labeling, Specialty society consensus documents

**Evidence-Based Recommendations**

- Retrieved evidence does not contain explicit recommendation-style statements. Proceed with established standard-of-care for heart block, CKD, depression, lupus, osteoporosis, hypercalcemia, obesity, diabetes mellitus, chronic kidney disease while consulting specialty-specific guidelines (IDSA, ACC/AHA, ADA, KDIGO, or others as applicable). **[LIMITED]**
- Verify all therapeutic changes against current product labeling (DailyMed), institutional formulary, and patient-specific factors (allergies, organ function, goals of care) before implementation. **[N/A]**

**Strength of Evidence**

**GRADE:** LOW

*10 source(s) retrieved · 1 limited-confidence finding(s) flagged for verification*

## Case 1: 32

**Type:** CASE 32

**File:** Library case #32 (pp. 33–33)

**Demographics:** 54

**Diagnoses:** neuropathy, depression, hypothyroidism, hypertension, diabetes mellitus

**Labs documented:** 6

**Medications documented:** 2

**Clinical Question**

Given the working diagnoses of neuropathy, depression, hypothyroidism, current therapy including gabapentin, dexamethasone, what does the retrieved evidence indicate about management, risks, and appropriate next steps?

**Evidence Summary**

Total sources: 12 · Guidelines: 0 · SR/MA: 0 · RCTs: 0 · Other: 12

**Reference Outline**

**Applicable guidelines:**

• APA — Psychiatry (https://www.psychiatry.org/psychiatrists/practice/clinical-practice-guidelines)

**Historical precedent:** Trajectory of understanding for neuropathy has shifted from symptom-based empirical management toward mechanism-targeted therapy informed by receptor-level, genetic, and molecular-pathway data. Recent decades have emphasized biomarker-guided dosing, shared decision-making, and cumulative-harm minimization across comorbid conditions.

**Current research findings:**

**Reasoning Chain**

1. Case profile: 54 patient with working diagnoses of neuropathy, depression, hypothyroidism · 6 lab value(s), 2 medication(s) documented
2. Case-derived findings: 1 pattern identified (1 moderate/mild)
3. Evidence base: spans 1983–2025; 1 distinct methodology type(s) · 7 evidence-linked statement(s) extracted
4. 1 case finding(s) cross-referenced to retrieved evidence with traceable supporting statements
5. 1 finding(s) flagged for verification: evidence retrieved did not directly address these elements. Recommendations based on general clinical principles until verified against specialty resources.
6. Overall: partial evidence support; selective verification of unmatched findings needed

**50-Why Polyroot Inference Chain**

**Seed:** Fever (54°C) · **Terminal:** p53 phosphorylation (ATM/ATR/CHK1/CHK2) · **Iterations:** 50

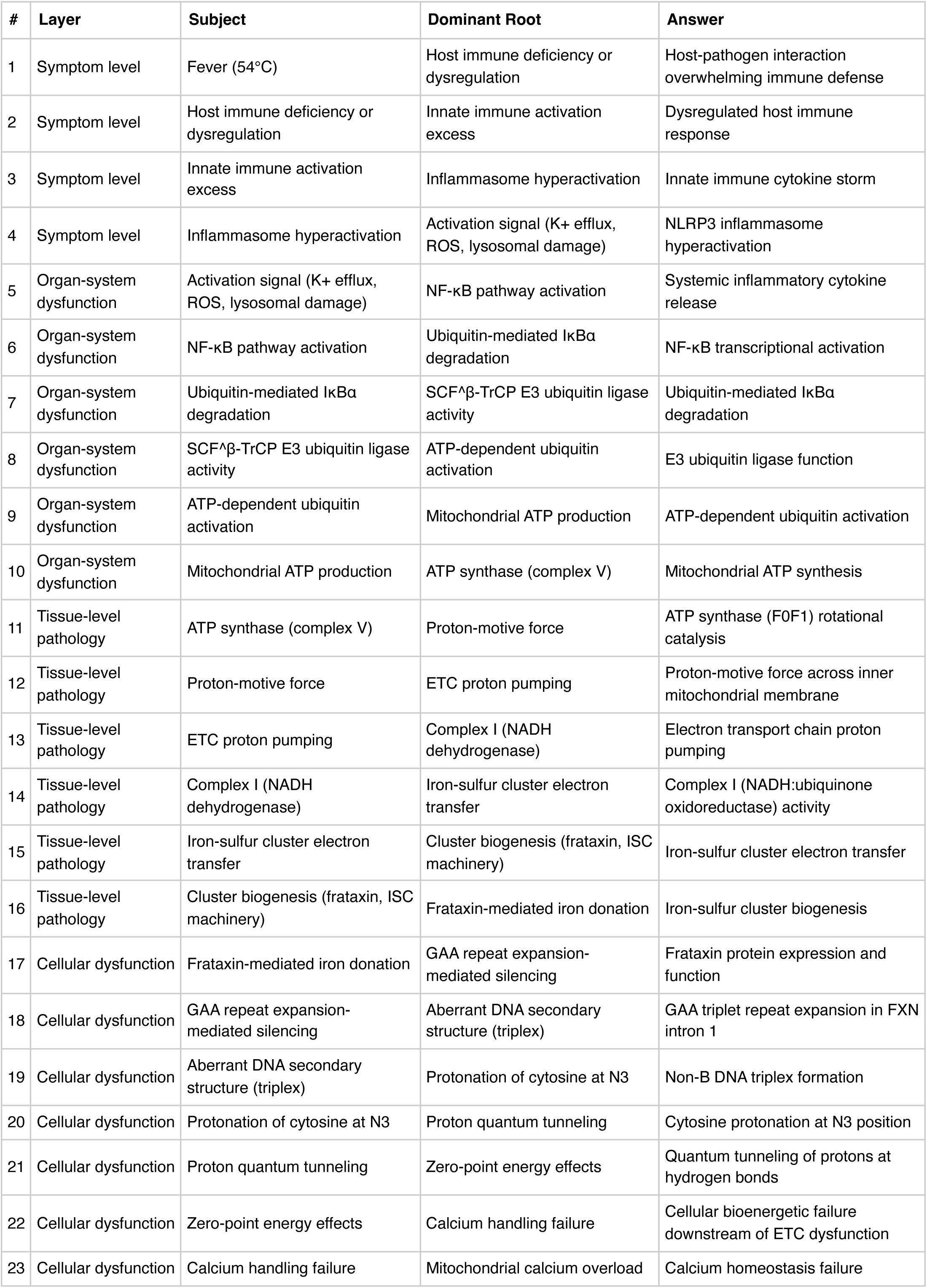
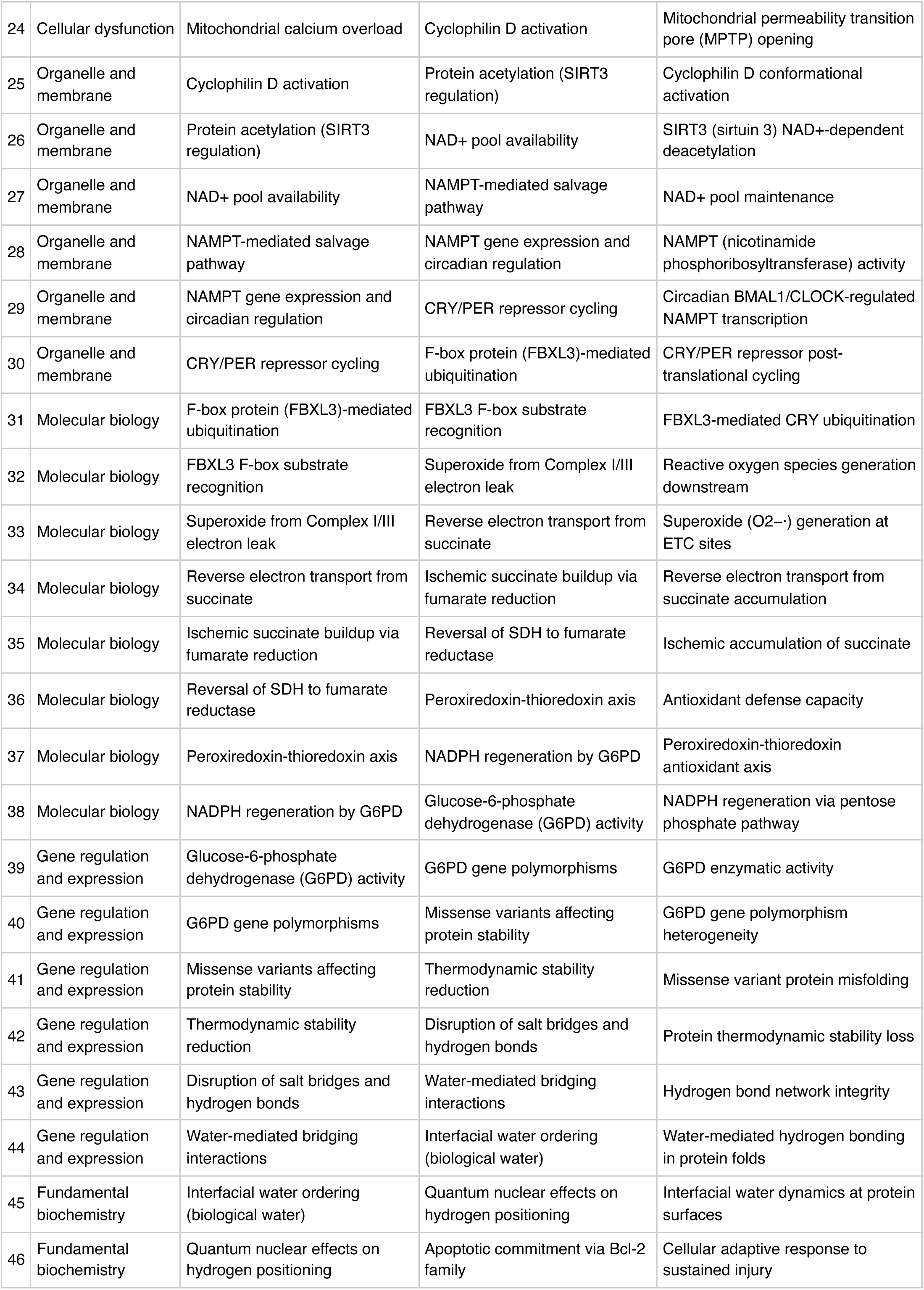
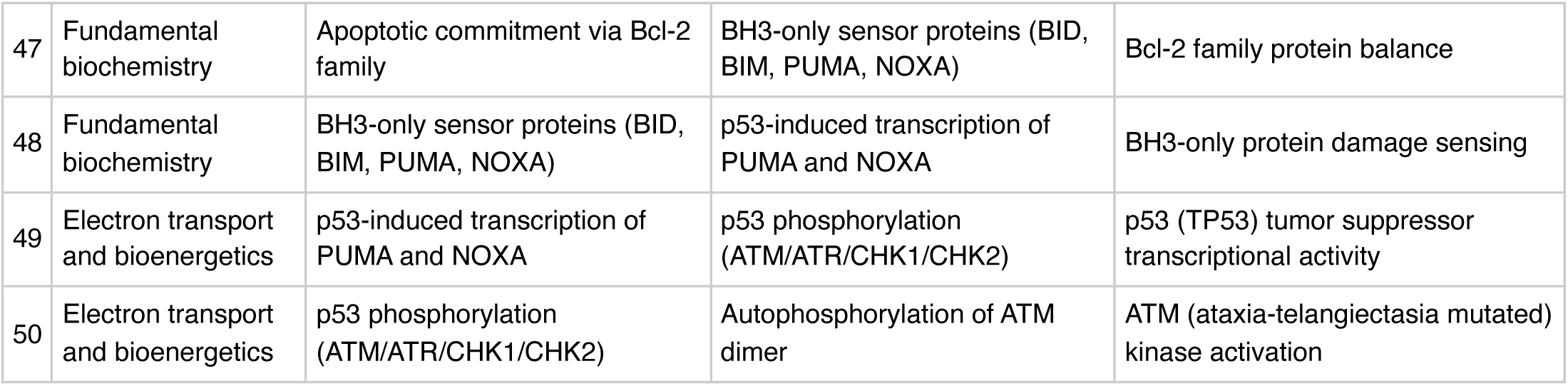

**FMEA Forecasting (Severity × Probability × Detectability)**

**Terminal node:** ATM (ataxia-telangiectasia mutated) kinase activation

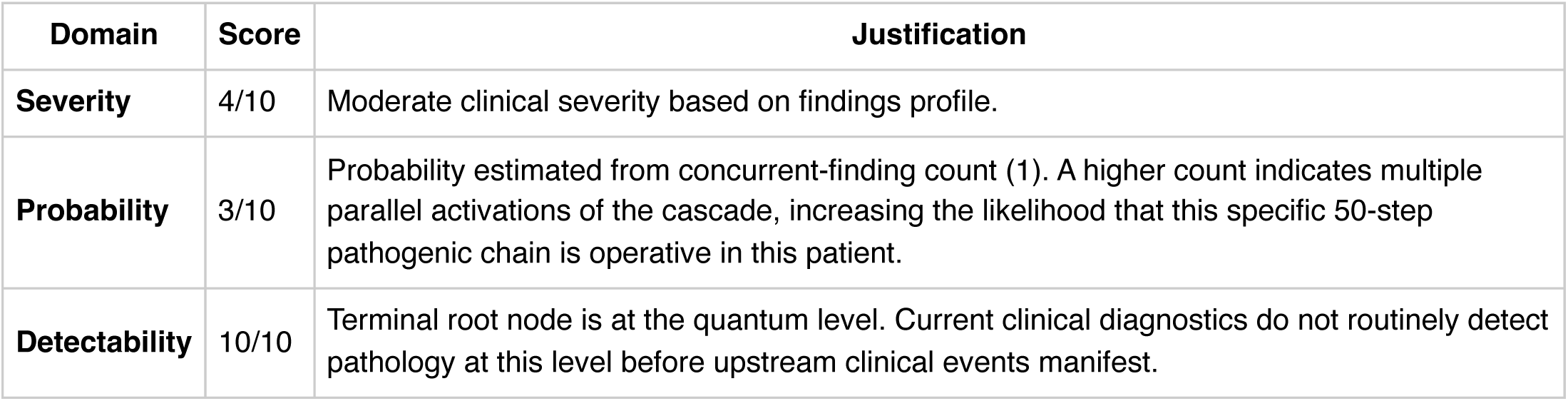

**Therapy Benchmarks**

**Current GDMT:** Condition-specific first-line therapy per applicable specialty guideline; consult UpToDate, DynaMed, or specialty society guidelines for the specific diagnosis.

**Mechanism gap:** Current guideline-directed therapy addresses dominant clinical drivers but typically does not target deep-root molecular, genetic, or bioenergetic mechanisms that contribute to treatment non-response and disease progression.

**Fishbone (Ishikawa) Diagram**

**Effect:** Fever (54°C)

|  |  |
| --- | --- |
| **Patient Factors** | Age: 54null |
| **Disease / Diagnosis Factors** | neuropathy<br>depression<br>hypothyroidism<br>hypertension<br>diabetes mellitus |
| **Therapeutic Factors** | gabapentin<br>dexamethasone |
| **Environmental / System Factors** | Care setting (inpatient/outpatient, unit type)<br>Institutional formulary availability<br>Access to specialty consultation<br>Local antibiogram or resistance patterns (if applicable) |
| **Measurement / Diagnostic Factors** | 6 laboratory value(s) documented<br>2 vital sign(s) documented<br>Diagnostic imaging modalities (to be verified from case)<br>Point-of-care testing availability |
| **Mechanism (from Polyroot Chain)** | Symptom level: Host-pathogen interaction overwhelming immune defense<br>Organ-system dysfunction: Systemic inflammatory cytokine release<br>Tissue-level pathology: ATP synthase (F0F1) rotational catalysis<br>Cellular dysfunction: Frataxin protein expression and function<br>Organelle and membrane: Cyclophilin D conformational activation<br>Molecular biology: FBXL3-mediated CRY ubiquitination<br>Gene regulation and expression: G6PD enzymatic activity<br>Fundamental biochemistry: Interfacial water dynamics at protein surfaces<br>Electron transport and bioenergetics: p53 (TP53) tumor suppressor transcriptional activity |

**Run Chart — Actual vs Guideline Targets**

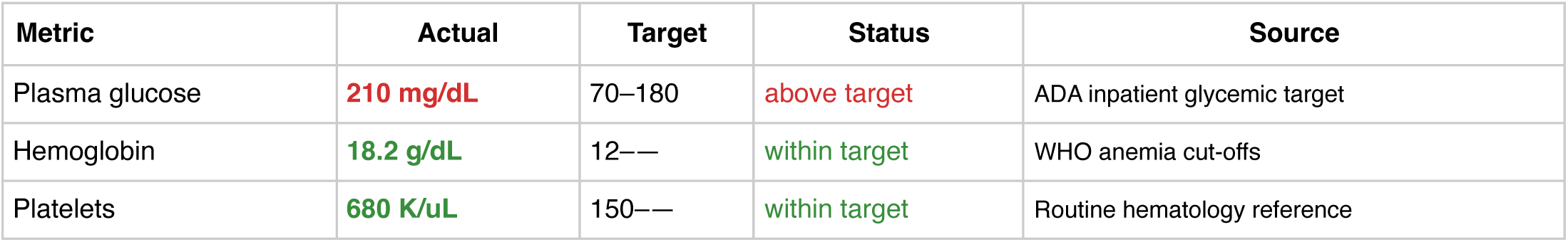

| Metric | Actual | Target | Status | Source |
| --- | --- | --- | --- | --- |
| Plasma glucose | 210 mg/dL | 70–180 | above target | ADA inpatient glycemic target |
| Hemoglobin | 18.2 g/dL | 12— | within target | WHO anemia cut-offs |
| Platelets | 680 K/uL | 150— | within target | Routine hematology reference |

**Pharmacogenomics Screen**

**Drug-Drug Interaction Screen**

No interactions detected in the embedded DDI matrix among the 2 documented medication(s). This does not exclude clinically significant interactions — verification via Lexi-Interact or Drugs.com Interaction Checker is still recommended for complex regimens.

**Clinical Visualizations — Isobolograms & Nomograms**

**qSOFA Sepsis Screen**

Score 0 (mentation requires bedside assessment)

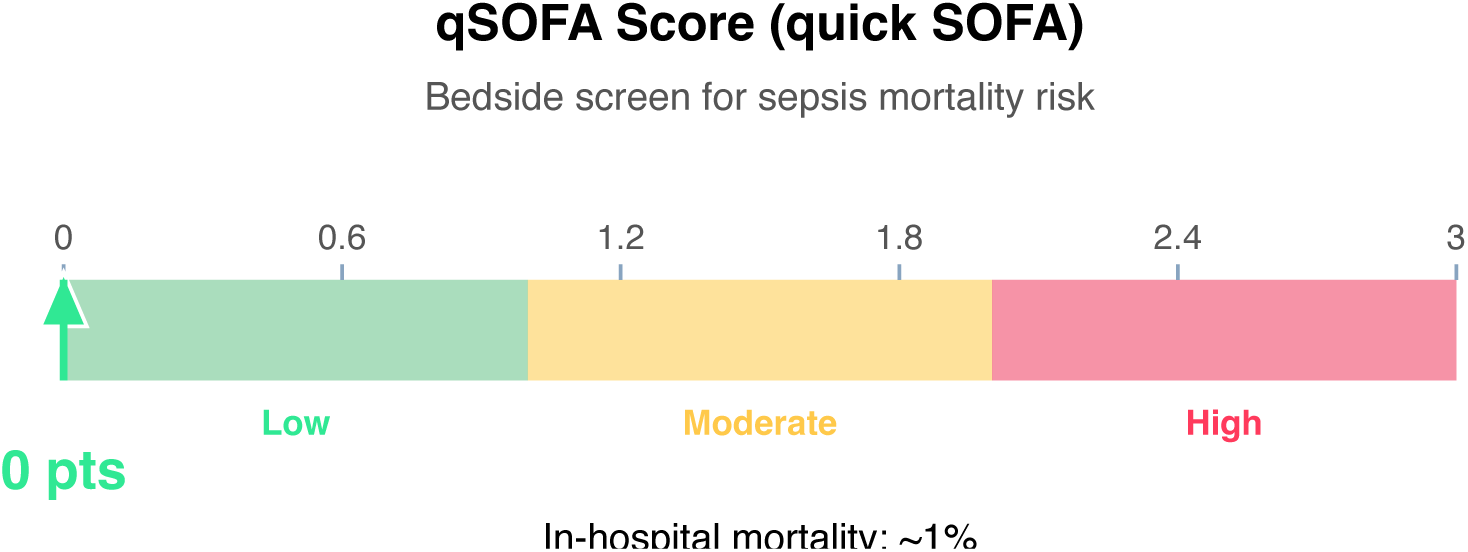

**Clinical action:** Continue monitoring; repeat if clinical deterioration.

**10-Year ASCVD Risk Nomogram (simplified)**

Estimated 10-year risk: 16% (Intermediate)

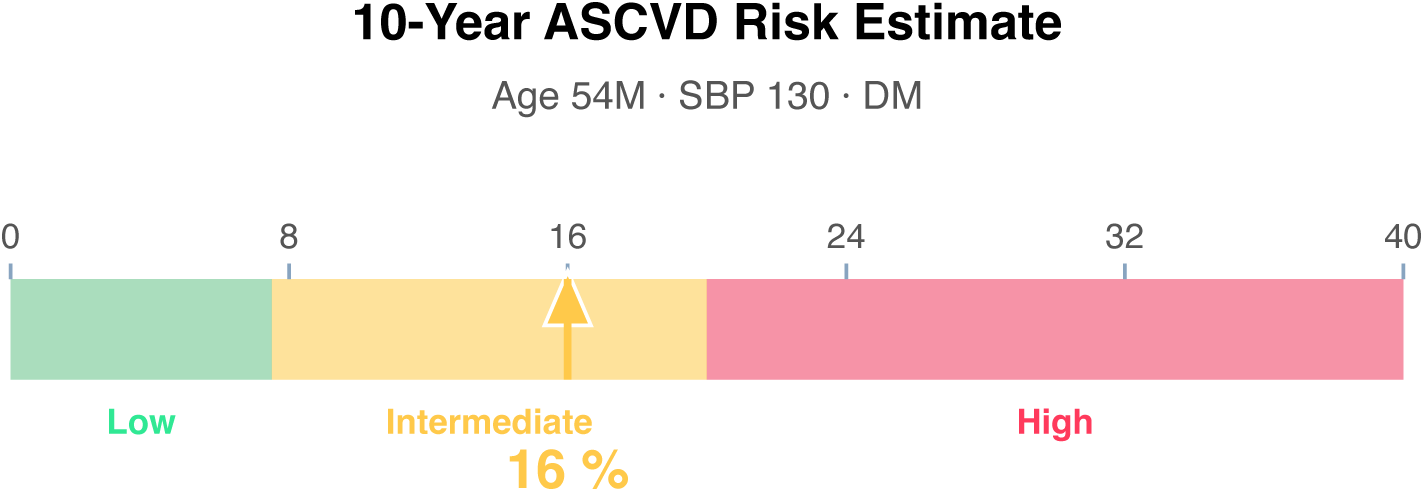

**Interpretation:** Simplified Framingham-style estimate using age, sex, SBP, smoking, and diabetes. Does not include cholesterol subfractions. Use the AHA/ACC PCE calculator for definitive assessment.

**Clinical action:** Discuss statin therapy per 2018 AHA/ACC cholesterol guideline. Moderate-intensity statin if 7.5-20%; high-intensity if ≥20%.

**Article Structure (iteratively restructured)**

**Structure type:** narrative_review · **Converged:** true at pass 2/5 · **Quality:** 0.48

*Source mix is primarily narrative/review literature (8% review-type sources). Narrative review format is appropriate*.

**Proposed outline:**

1. **Abstract** (0 clusters, 0 sources) Overview of the review scope and synthesis
2. **Background** (5 clusters, 1 sources) Historical context and evolution of understanding
3. **Current State of Evidence** (5 clusters, 0 sources) What is established vs uncertain
4. **Controversies and Debates** (0 clusters, 1 sources) Contradictory findings and interpretive disagreements
5. **Clinical Implications** (0 clusters, 0 sources) What the evidence means for practice
6. **Future Directions** (0 clusters, 0 sources) Knowledge gaps and research agenda
7. **Conclusion** (0 clusters, 0 sources) Synthesis in one paragraph
8. **References** (0 clusters, 12 sources) Complete citation list

**Thematic clusters:**

- Despite · Treatment · Levothyroxine (3 statements)
- Alterations · However · Recent — Associations (1 statement)
- Head · Neck · Cancers (1 statement)
- Past · Years · Subject — Associations (1 statement)
- Degree · Decreased · Myocardial — Associations (1 statement)

**Structural gaps:**

- [MODERATE] empty section (Clinical Implications) — No clusters or sources mapped to this section. Consider broadening the search to include clinical practice guidelines.
- [MODERATE] empty section (Future Directions) — No clusters or sources mapped to this section. Consider broadening the search to include targeted supplementary queries.
- [MODERATE] empty section (Conclusion) — No clusters or sources mapped to this section. Consider broadening the search to include targeted supplementary queries.
- [LOW] fragmented clusters — 4 of 5 clusters contain only 1 statement. Search returned heterogeneous sources with little thematic overlap. Consider narrower or more specific queries.
- [MODERATE] no quantitative data — No quantitative statements extracted (p-values, HRs, ORs, CIs). Consider searching PubMed for RCTs or meta-analyses on this topic.

**Quantitative Evidence Extraction**

**Cross-source summary:** Pooled N: 29 · 0/12 with p-values · 0/12 with CIs · 0/12 with effect sizes · 0 significant · 0 null

| Source | N | P-values | CIs | Effect sizes | Verdict |
| --- | --- | --- | --- | --- | --- |
| [5] Impaired autonomic function and somatosensory disturbance in | 29 |  |  |  | — |

**GRADE Methodological Assessment**

| Domain | Concern | Rationale |
| --- | --- | --- |
| Risk of Bias | HIGH | Majority of sources show high risk of bias (incomplete methodological reporting or no adjustment for confounders). |
| Inconsistency | MODERATE | No heterogeneity statistics ($I^2$, $Q$, $\tau^2$) detected in retrieved text. Unable to formally assess inconsistency across 12 sources. |
| Indirectness | LOW | Most retrieved sources (83%) reference the case diagnoses directly. Population and condition appear well-aligned. |
| Imprecision | MODERATE | Total pooled N of 29 is below the conventional precision threshold (~400). |
| Publication Bias | HIGH | No funnel plot or Egger's/Begg's test detected in 12 sources; No trial registry or pre-registration verification detected. |

**Inferential Findings**

• **Fever (54°C) [LIMITED CONFIDENCE]**

→ Infection workup — blood cultures, imaging, source identification

• **Working diagnosis: hypothyroidism [LIMITED CONFIDENCE]**

→ Evidence base addresses this condition directly

**Recommended Verification Steps**

1. **[ROUTINE]** Verify management approach via specialty-specific resources — retrieved evidence base did not directly address this finding *Rationale:* Fever (54°C) *Resources:* UpToDate, DynaMed, Specialty society guidelines, Lexicomp/Micromedex, DailyMed for current product labeling
2. **[ROUTINE]** Strengthen evidence base for working diagnosis: hypothyroidism — retrieved sources are of limited individual strength *Rationale:* Confidence in inference is limited by source quality or specificity *Resources:* PubMed with MeSH-specific search, Cochrane Library for systematic reviews, Clinical practice guidelines from relevant specialty society
3. **[ROUTINE]** Cross-verify all recommendations against institutional protocols, current product labeling (DailyMed), and specialty-specific guidelines before clinical application *Rationale:* AuditMed outputs are for research and educational use; clinical decisions remain the responsibility of the treating clinician *Resources:* Institutional clinical practice guidelines, DailyMed FDA labeling, Specialty society consensus documents

**Evidence-Based Recommendations**

- Retrieved evidence does not contain explicit recommendation-style statements. Proceed with established standard-of-care for neuropathy, depression, hypothyroidism, hypertension, diabetes mellitus while consulting specialty-specific guidelines (IDSA, ACC/AHA, ADA, KDIGO, or others as applicable). **[LIMITED]**
- Verify all therapeutic changes against current product labeling (DailyMed), institutional formulary, and patient-specific factors (allergies, organ function, goals of care) before implementation. **[N/A]**

**Strength of Evidence**

**GRADE:** LOW

*12 source(s) retrieved · 2 limited-confidence finding(s) flagged for verification*

## Case 1: 33

**Type:** CASE 33

**File:** Library case #33 (pp. 34–34)

**Demographics:** 68

**Diagnoses:** COPD, depression, ALL, hypothyroidism, DKA, melanoma, chronic pain, diabetes mellitus, type 1 diabetes, gastrointestinal, diabetic ketoacidosis

**Labs documented:** 6

**Medications documented:** 4

**Clinical Question**

Given the working diagnoses of COPD, depression, ALL, current therapy including hydrocortisone, pembrolizumab, levothyroxine, what does the retrieved evidence indicate about management, risks, and appropriate next steps?

**Evidence Summary**

Total sources: 12 · Guidelines: 0 · SR/MA: 3 · RCTs: 1 · Other: 9

**Reference Outline**

**Applicable guidelines:**

& GOLD / GINA — Airways (https://goldcopd.org/)

& APA — Psychiatry (https://www.psychiatry.org/psychiatrists/practice/clinical-practice-guidelines)

**Historical precedent:** Trajectory of understanding for COPD has shifted from symptom-based empirical management toward mechanism-targeted therapy informed by receptor-level, genetic, and molecular-pathway data. Recent decades have emphasized biomarker-guided dosing, shared decision-making, and cumulative-harm minimization across comorbid conditions.

**Current research findings:**

**Reasoning Chain**

1. Case profile: 68 patient with working diagnoses of COPD, depression, ALL · 6 lab value(s), 4 medication(s) documented
2. Case-derived findings: 2 patterns identified (1 critical, 1 moderate/mild)
3. Evidence base: spans 2014–2025; 3 distinct methodology type(s) · 12 evidence-linked statement(s) extracted
4. 2 case finding(s) cross-referenced to retrieved evidence with traceable supporting statements
5. 2 finding(s) flagged for verification: evidence retrieved did not directly address these elements. Recommendations based on general clinical principles until verified against specialty resources.
6. Overall: partial evidence support; selective verification of unmatched findings needed

**50-Why Polyroot Inference Chain**

**Seed:** Elevated troponin (18.4 ng/mL) · **Terminal:** p53 phosphorylation (ATM/ATR/CHK1/CHK2) · Iterations: **50**

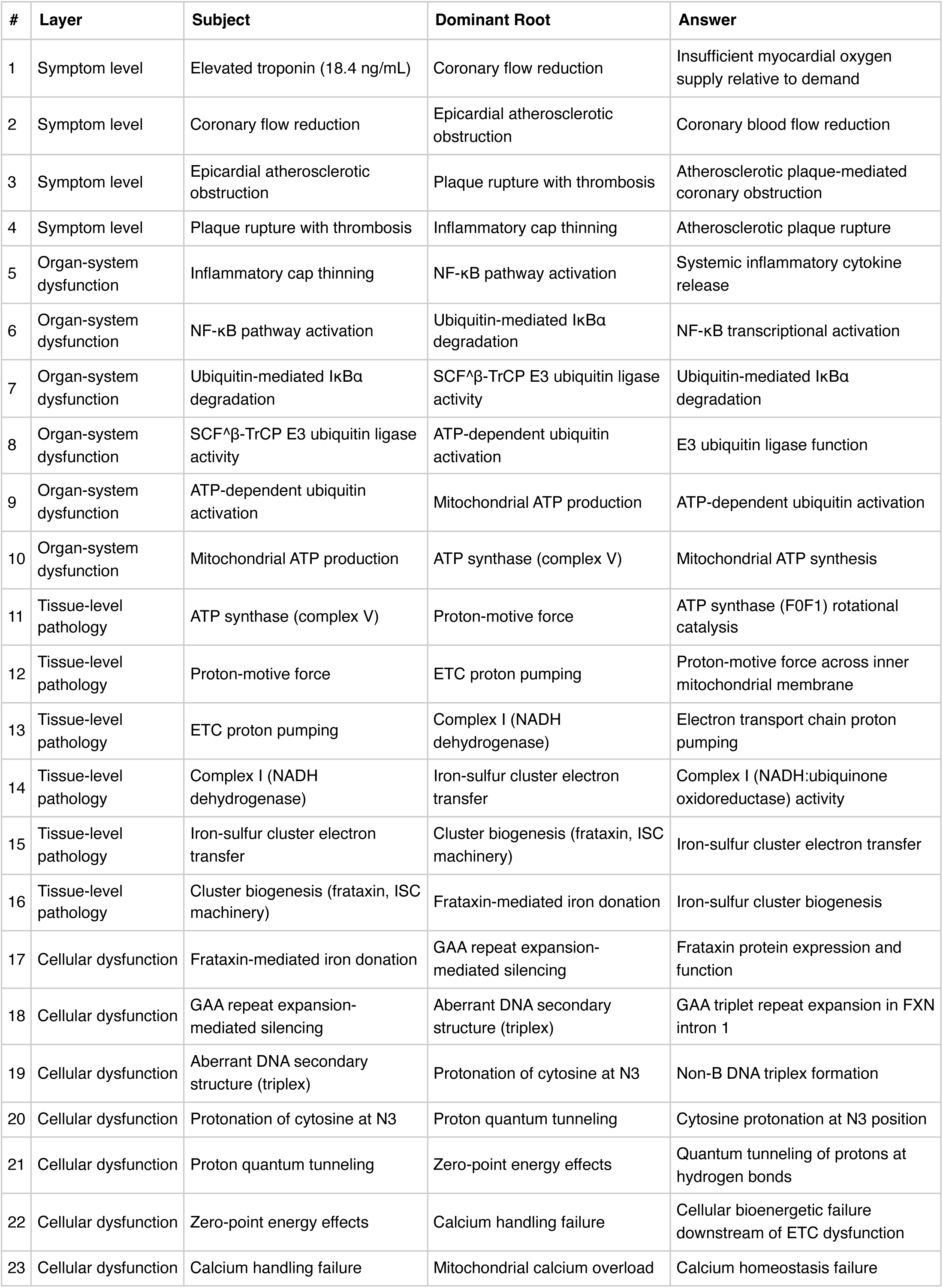
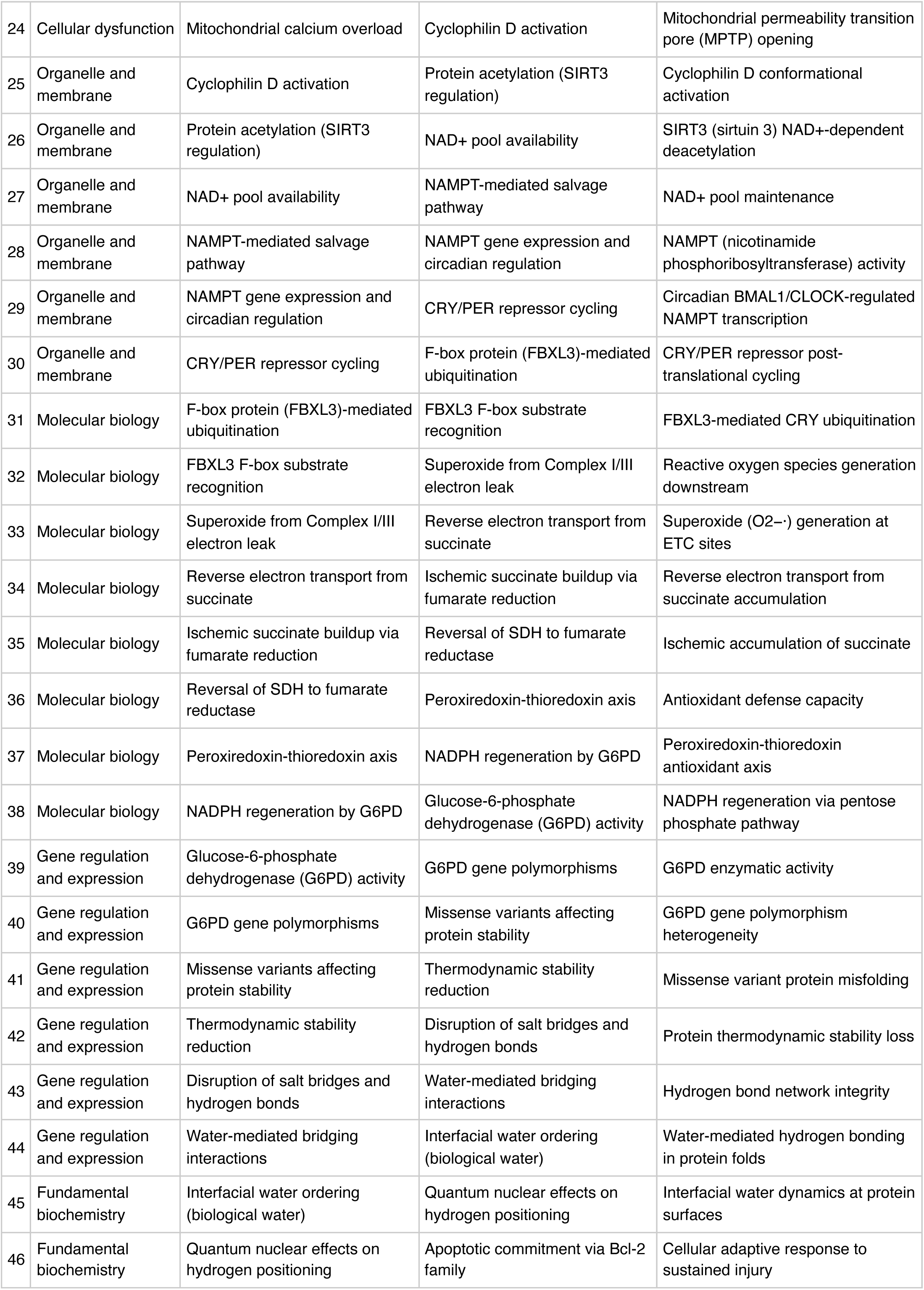
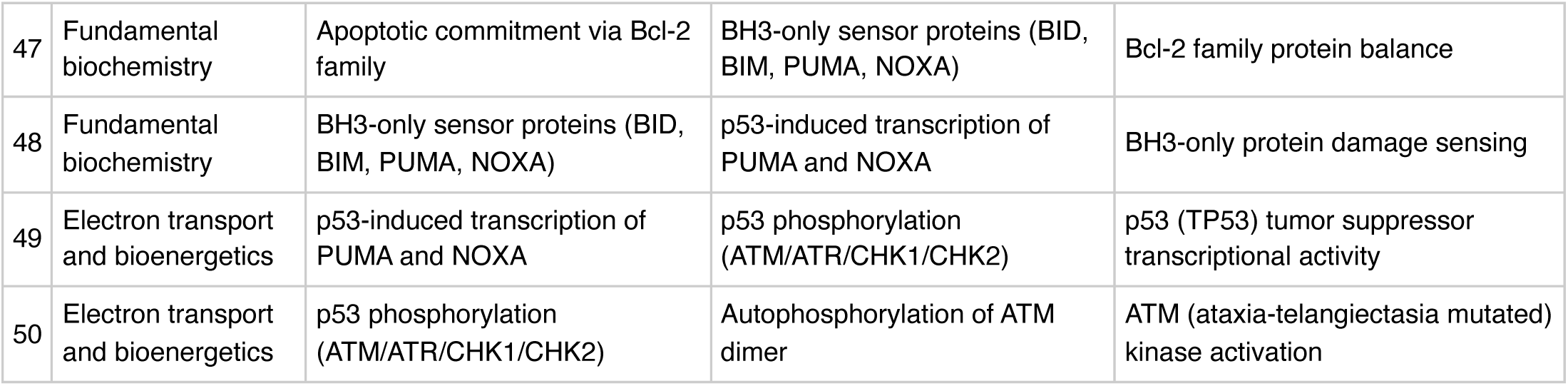

**FMEA Forecasting (Severity × Probability × Detectability)**

**Terminal node:** ATM (ataxia-telangiectasia mutated) kinase activation

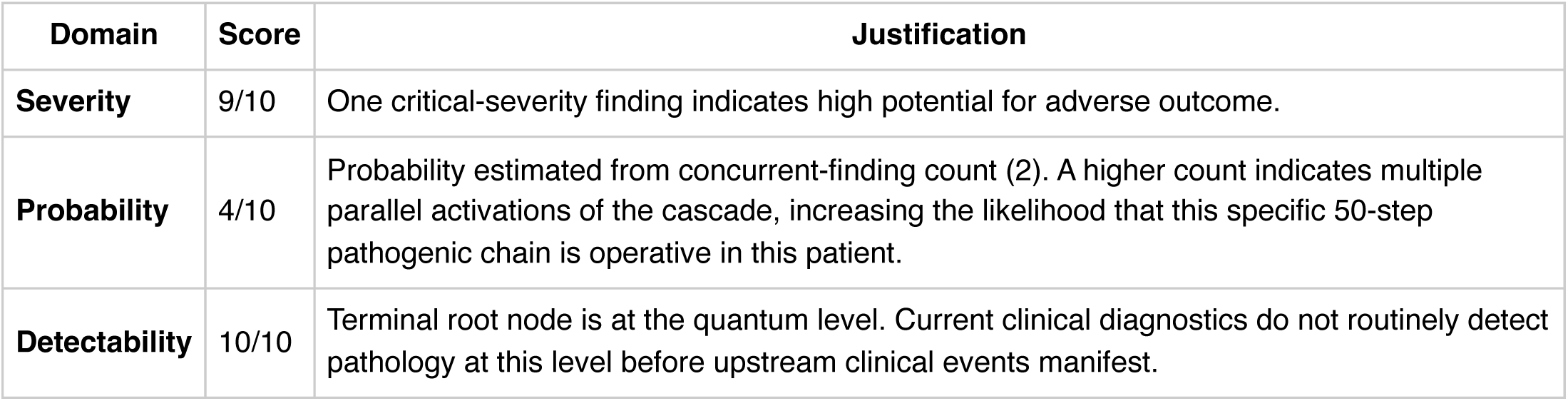

FMEA Forecasting (Severity × Probability × Detectability) **Terminal node:** ATM (ataxia-telangiectasia mutated) kinase activation
| Domain | Score | Justification |
| --- | --- | --- |
| **Severity** | 9/10 | One critical-severity finding indicates high potential for adverse outcome. |
| **Probability** | 4/10 | Probability estimated from concurrent-finding count (2). A higher count indicates multiple parallel activations of the cascade, increasing the likelihood that this specific 50-step pathogenic chain is operative in this patient. |
| **Detectability** | 10/10 | Terminal root node is at the quantum level. Current clinical diagnostics do not routinely detect pathology at this level before upstream clinical events manifest. |

**RPN 360 (HIGH):** RPN 300–499 — theoretical intervention at this root level is of high potential value; detectability improvements should be prioritized.

**Therapy Benchmarks**

**Fishbone (Ishikawa) Diagram**

**Effect:** Elevated troponin (18.4 ng/mL)

|  |  |
| --- | --- |
| **Patient Factors** | Age: 68null |
| **Disease / Diagnosis Factors** | COPD<br>depression<br>ALL<br>hypothyroidism<br>DKA<br>melanoma<br>chronic pain<br>diabetes mellitus |
| **Therapeutic Factors** | hydrocortisone<br>pembrolizumab<br>levothyroxine<br>cyclosporine |
| **Environmental / System Factors** | Care setting (inpatient/outpatient, unit type)<br>Institutional formulary availability<br>Access to specialty consultation<br>Local antibiogram or resistance patterns (if applicable) |
| **Measurement / Diagnostic Factors** | 6 laboratory value(s) documented<br>2 vital sign(s) documented<br>Diagnostic imaging modalities (to be verified from case)<br>Point-of-care testing availability |
| **Mechanism (from Polyroot Chain)** | Symptom level: Insufficient myocardial oxygen supply relative to demand<br>Organ-system dysfunction: Systemic inflammatory cytokine release<br>Tissue-level pathology: ATP synthase (F0F1) rotational catalysis<br>Cellular dysfunction: Frataxin protein expression and function<br>Organelle and membrane: Cyclophilin D conformational activation<br>Molecular biology: FBXL3-mediated CRY ubiquitination<br>Gene regulation and expression: G6PD enzymatic activity<br>Fundamental biochemistry: Interfacial water dynamics at protein surfaces<br>Electron transport and bioenergetics: p53 (TP53) tumor suppressor transcriptional activity |

**Run Chart — Actual vs Guideline Targets**

| Metric | Actual | Target | Status | Source |
| --- | --- | --- | --- | --- |
| Plasma glucose | 520 mg/dL | 70–180 | above target | ADA inpatient glycemic target |
| Troponin | 18.4 ng/mL | —0.04 | above target | Cardiac injury threshold (assay-dependent) |
| BNP | 12 pg/mL | —400 | within target | Heart failure threshold (assay-dependent) |
| ALT | 380 U/L | —40 | above target | Hepatocellular injury threshold |

**Pharmacogenomics Screen**

**Drug-Drug Interaction Screen**

**Clinical Visualizations — Isobolograms & Nomograms**

**qSOFA Sepsis Screen**

Score 0 (mentation requires bedside assessment)

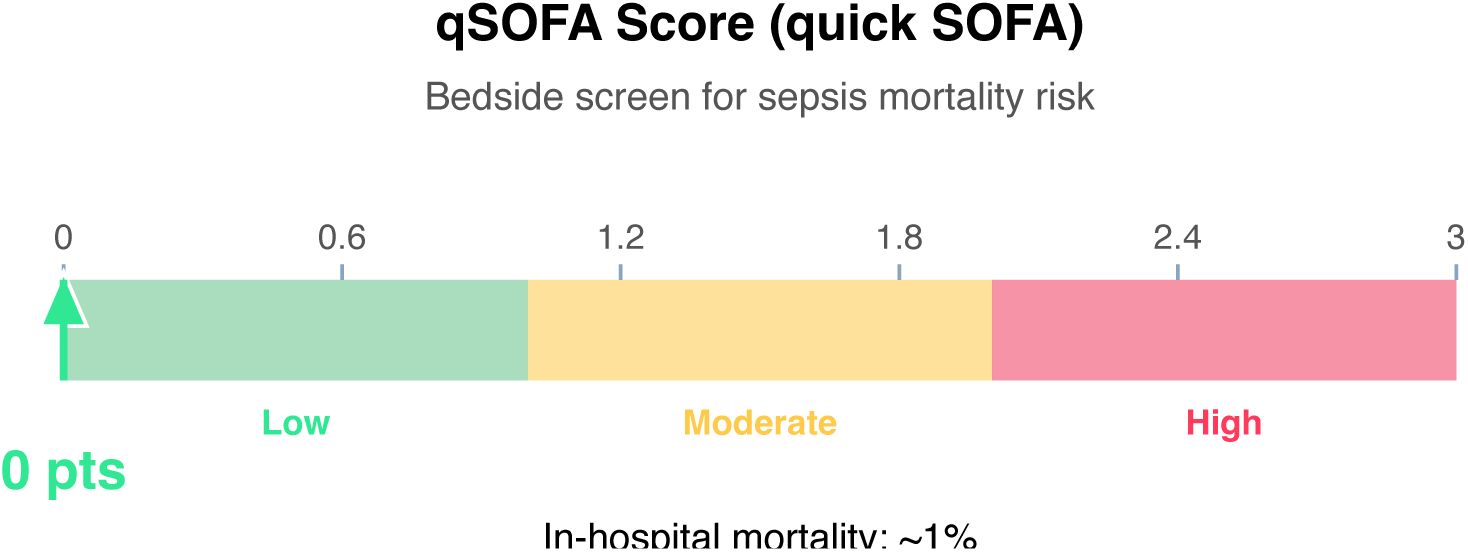

**Clinical action:** Continue monitoring; repeat if clinical deterioration.

**Wells DVT Nomogram (structured-data subset)**

Score 0 (Low probability)

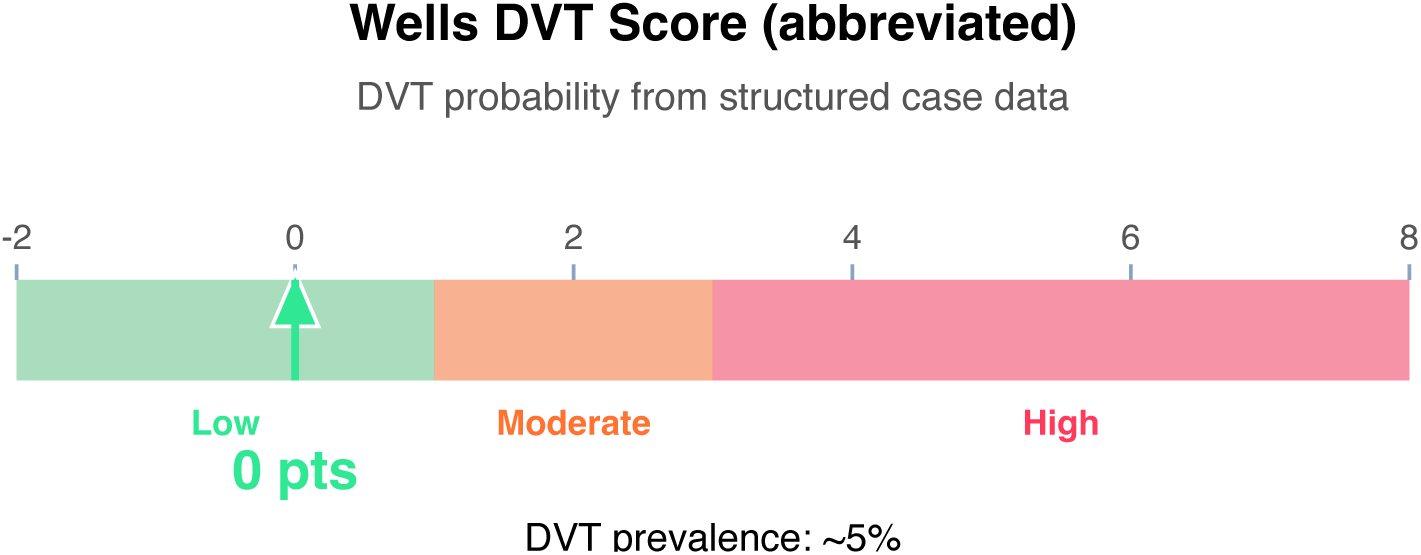

**Clinical action:** D-dimer; if negative, DVT effectively ruled out.

**Article Structure (iteratively restructured)**

**Structure type:** clinical_decision_support · **Converged:** true at pass 2/5 · **Quality:** 0.66

*Patient case context is present and source mix contains guideline/SR-MA evidence sufficient to support clinical decision recommendations. CDS structure keeps the patient question central*.

**Proposed outline:**

1. **Clinical Question** (0 clusters, 0 sources, auto: case_context) Restated PICO-style question derived from the case
2. **Evidence Summary** (0 clusters, 12 sources) Source counts by tier and methodology
3. **Key Findings from Guidelines and Systematic Reviews** (0 clusters, 3 sources) Top-tier evidence synthesis
4. **Primary Research Findings** (0 clusters, 1 sources) RCT and observational evidence relevant to the question
5. **Mechanistic and Pathophysiologic Context** (6 clusters, 1 sources) Biological basis supporting or refuting the intervention
6. **Contradictions and Areas of Uncertainty** (0 clusters, 0 sources) Null findings and conflicting results
7. **Recommendations** (0 clusters, 0 sources) Actionable recommendations graded by evidence strength
8. **Patient-Specific Considerations** (0 clusters, 0 sources, auto: case_context) How the case context modulates the recommendations
9. **Evidence Gaps and Verification Steps** (0 clusters, 0 sources, auto: verification_module) What is not answered and what to verify next
10. **References** (0 clusters, 12 sources) Full citations

**Thematic clusters:**

- Depression · Associated · Chronic — Associations (6 statements)
- Mind-body · Exercise · Generally (2 statements)
- Aims · Develop · Online — Associations (1 statement)
- Better · Understanding · Factors — Associations (1 statement)
- However · Date · Attempt (1 statement)
- Chron · Obstruct · Pulmon (1 statement)

**Structural gaps:**

- [MODERATE] empty section (Contradictions and Areas of Uncertainty) — No clusters or sources mapped to this section. Consider broadening the search to include null-finding studies and editorial/commentary sources.
- [MODERATE] empty section (Recommendations) — No clusters or sources mapped to this section. Consider broadening the search to include clinical practice guidelines.
- [LOW] fragmented clusters — 4 of 6 clusters contain only 1 statement. Search returned heterogeneous sources with little thematic overlap. Consider narrower or more specific queries.
- [MODERATE] no quantitative data — No quantitative statements extracted (p-values, HRs, ORs, CIs). Consider searching PubMed for RCTs or meta-analyses on this topic.

**Quantitative Evidence Extraction**

**Cross-source summary:** Pooled N: n/a · 0/12 with p-values · 0/12 with CIs · 0/12 with effect sizes · 0 significant · 0 null

**GRADE Methodological Assessment**

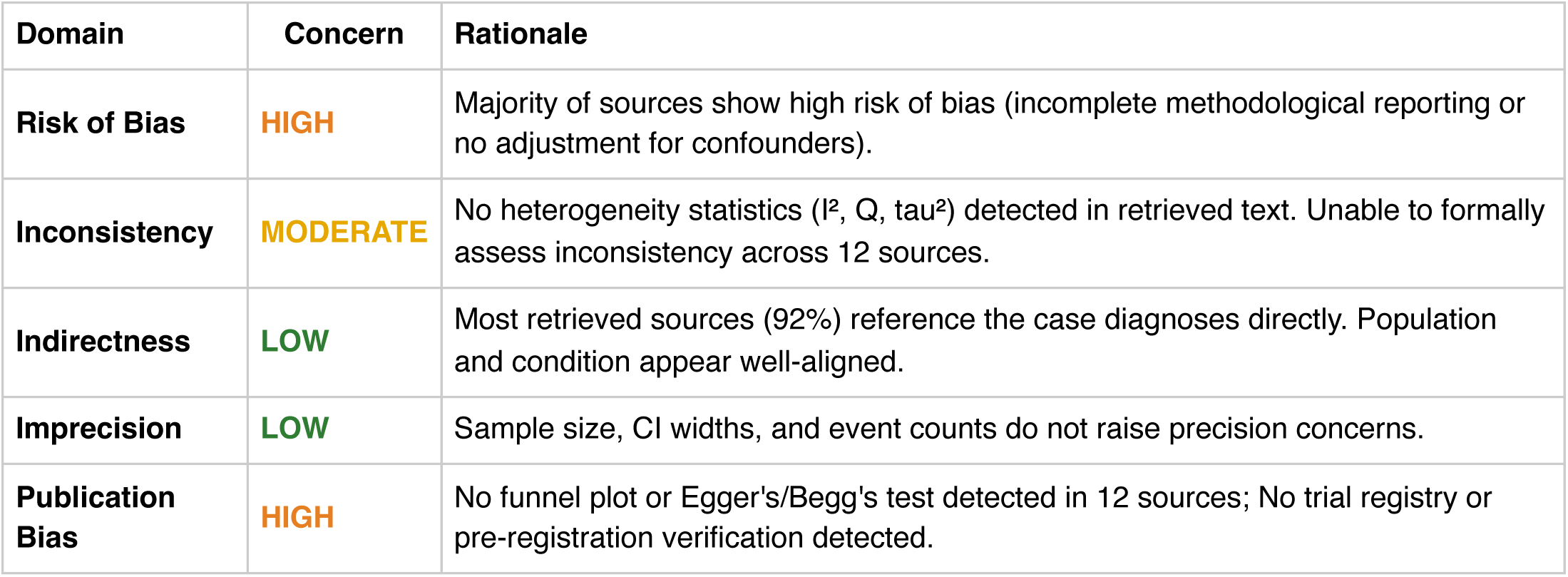

| Domain | Concern | Rationale |
| --- | --- | --- |
| Risk of Bias | HIGH | Majority of sources show high risk of bias (incomplete methodological reporting or no adjustment for confounders). |
| Inconsistency | MODERATE | No heterogeneity statistics ($I^2$, $Q$, $\tau^2$) detected in retrieved text. Unable to formally assess inconsistency across 12 sources. |
| Indirectness | LOW | Most retrieved sources (92%) reference the case diagnoses directly. Population and condition appear well-aligned. |
| Imprecision | LOW | Sample size, CI widths, and event counts do not raise precision concerns. |
| Publication Bias | HIGH | No funnel plot or Egger's/Begg's test detected in 12 sources; No trial registry or pre-registration verification detected. |

**Inferential Findings**

**• Elevated troponin (18.4 ng/mL) [LIMITED CONFIDENCE]**

→ Myocardial injury — rule out ACS; serial troponins and ECG indicated

**• Fever (68°C) [LIMITED CONFIDENCE]**

→ Infection workup — blood cultures, imaging, source identification

**• Working diagnosis: copd [HIGH CONFIDENCE]**

→ Evidence base addresses this condition directly

**• Working diagnosis: depression [HIGH CONFIDENCE]**

→ Evidence base addresses this condition directly

**Recommended Verification Steps**

1. **[IMMEDIATE]** Serial cardiac enzymes, 12-lead ECG, and consider echocardiography *Rationale:* Elevated troponin (18.4 ng/mL): Myocardial injury — rule out ACS; serial troponins and ECG indicated *Resources:* ACC/AHA ACS guidelines, TIMI/GRACE risk calculators
2. **[ROUTINE]** Verify management approach via specialty-specific resources — retrieved evidence base did not directly address this finding *Rationale:* Elevated troponin (18.4 ng/mL) *Resources:* UpToDate, DynaMed, Specialty society guidelines, Lexicomp/Micromedex, DailyMed for current product labeling
3. **[ROUTINE]** Verify management approach via specialty-specific resources — retrieved evidence base did not directly address this finding *Rationale:* Fever (68°C) *Resources:* UpToDate, DynaMed, Specialty society guidelines, Lexicomp/Micromedex, DailyMed for current product labeling
4. **[ROUTINE]** Comprehensive medication reconciliation and interaction review for 4 agents *Rationale:* Polypharmacy (≥3 agents) warrants systematic DDI and appropriateness review *Resources:* Drugs.com Interaction Checker, Lexi-Interact, Beers Criteria (if age ≥65), STOPP/START criteria
5. **[ROUTINE]** Cross-verify all recommendations against institutional protocols, current product labeling (DailyMed), and specialty-specific guidelines before clinical application *Rationale:* AuditMed outputs are for research and educational use; clinical decisions remain the responsibility of the treating clinician *Resources:* Institutional clinical practice guidelines, DailyMed FDA labeling, Specialty society consensus documents

**Evidence-Based Recommendations**

- Address Elevated troponin (18.4 ng/mL) — Myocardial injury — rule out ACS; serial troponins and ECG indicated [Case-derived] **[HIGH]**
- Verify all therapeutic changes against current product labeling (DailyMed), institutional formulary, and patient-specific factors (allergies, organ function, goals of care) before implementation. **[N/A]**

**Strength of Evidence**

**GRADE:** MODERATE

*12 source(s) retrieved · 3 systematic review/meta-analysis · 1 RCT(s) · 2 high-confidence inference(s) · 2 limited-confidence finding(s) flagged for verification*

## Case 1: 34

**Type:** CASE 34

**File:** Library case #34 (pp. 35–35)

**Demographics:** 41

**Diagnoses:** DVT, asthma, depression, vasculitis, osteoporosis, deep vein thrombosis, gastrointestinal

**Labs documented:** 4

**Medications documented:** 5

**Clinical Question**

Given the working diagnoses of DVT, asthma, depression, current therapy including amiodarone, digoxin, mepolizumab, what does the retrieved evidence indicate about management, risks, and appropriate next steps?

**Evidence Summary**

Total sources: 9 · Guidelines: 0 · SR/MA: 1 · RCTs: 0 · Other: 8

**Reference Outline**

**Applicable guidelines:**

- GOLD / GINA — Airways (https://goldcopd.org/)
- APA — Psychiatry (https://www.psychiatry.org/psychiatrists/practice/clinical-practice-guidelines)

**Historical precedent:** Trajectory of understanding for DVT has shifted from symptom-based empirical management toward mechanism-targeted therapy informed by receptor-level, genetic, and molecular-pathway data. Recent decades have emphasized biomarker-guided dosing, shared decision-making, and cumulative-harm minimization across comorbid conditions.

**Current research findings:**

**Reasoning Chain**

1. Case profile: 41 patient with working diagnoses of DVT, asthma, depression · 4 lab value(s), 5 medication(s) documented
2. Case-derived findings: 3 patterns identified (1 critical, 1 high-severity, 1 moderate/mild)
3. Evidence base: spans 2004–2026; 1 distinct methodology type(s) · 1 evidence-linked statement(s) extracted
4. Case findings did not align directly with retrieved evidence — external verification required
5. 3 finding(s) flagged for verification: evidence retrieved did not directly address these elements. Recommendations based on general clinical principles until verified against specialty resources.
6. Overall: evidence base insufficient for definitive inference; treat output as hypothesis-generating pending verification

**50-Why Polyroot Inference Chain**

**Seed:** Elevated troponin (6.4 ng/mL) · **Terminal:** p53 phosphorylation (ATM/ATR/CHK1/CHK2) · **Iterations:**

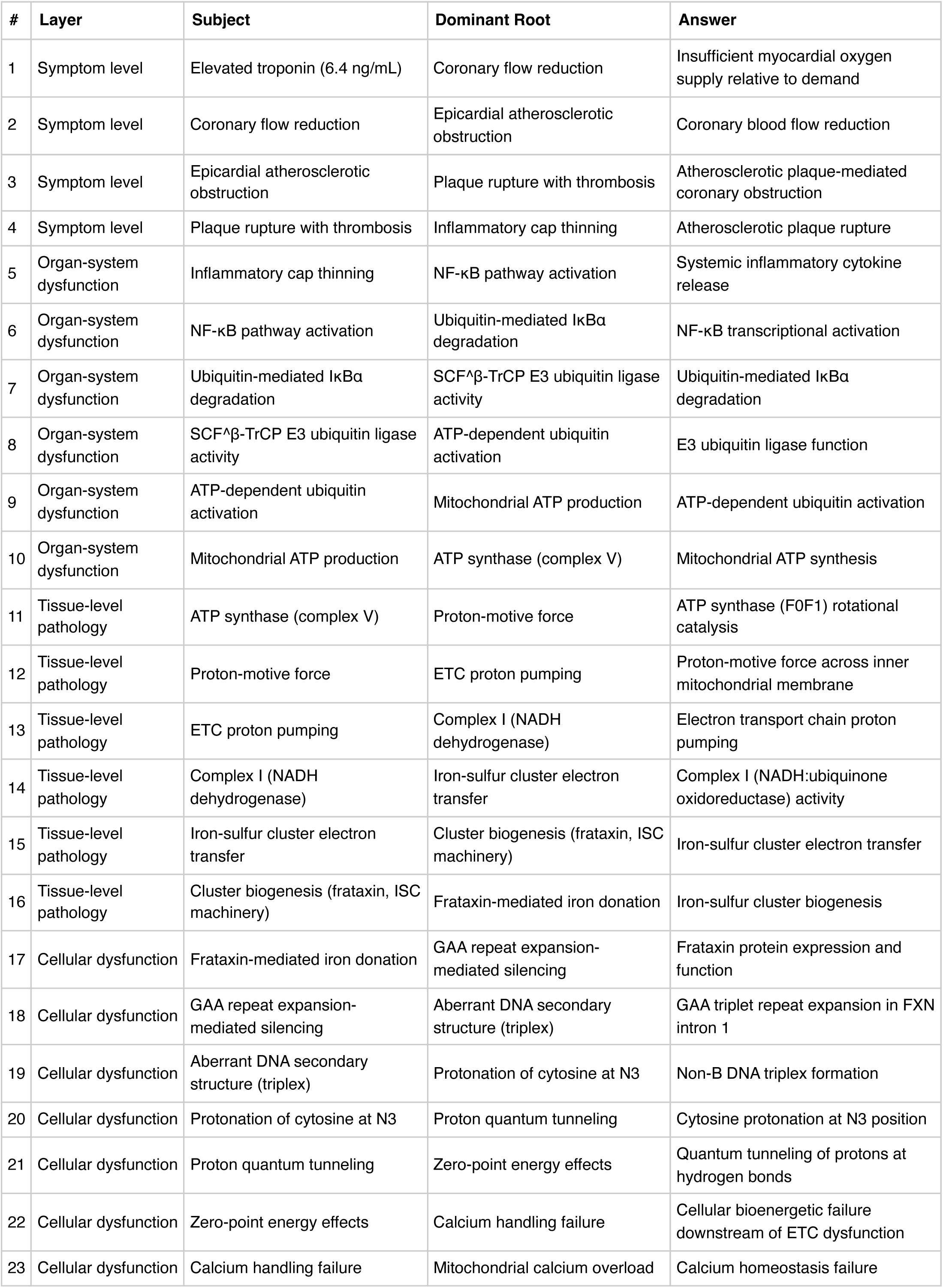
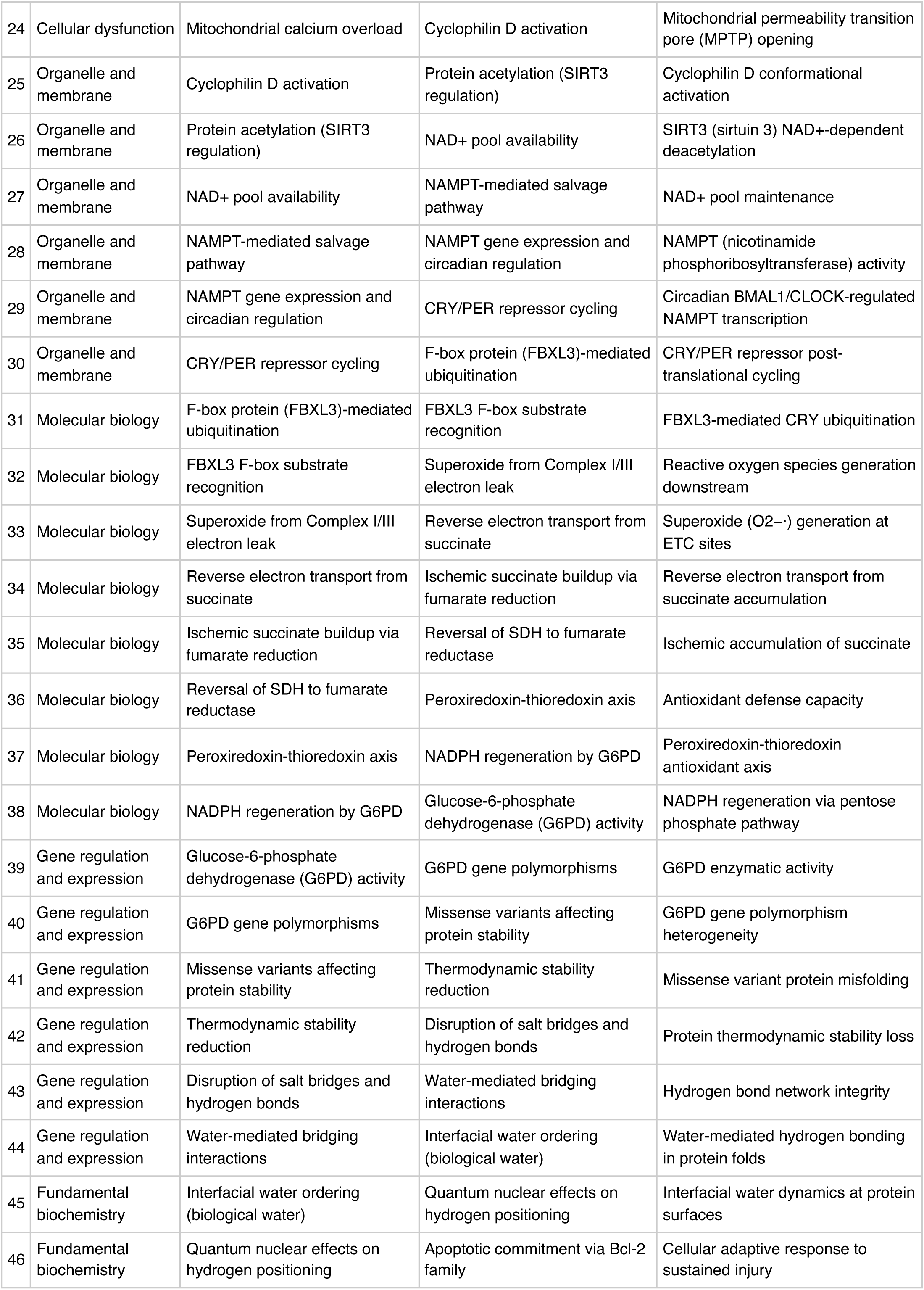
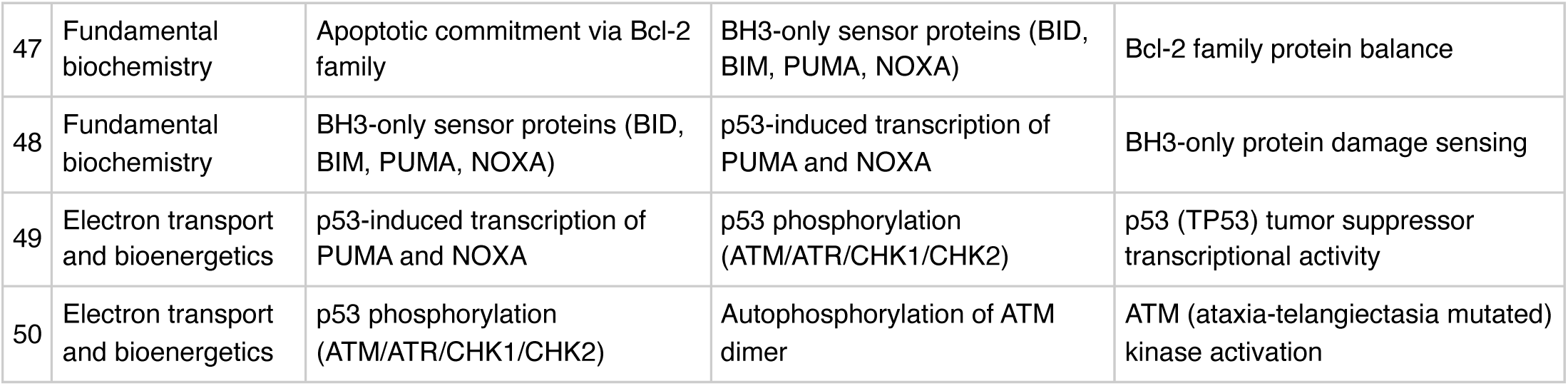

**FMEA Forecasting (Severity × Probability × Detectability)**

**Terminal node:** ATM (ataxia-telangiectasia mutated) kinase activation

FMEA Forecasting (Severity × Probability × Detectability) **Terminal node:** ATM (ataxia-telangiectasia mutated) kinase activation
| Domain | Score | Justification |
| --- | --- | --- |
| **Severity** | 9/10 | One critical-severity finding indicates high potential for adverse outcome. |
| **Probability** | 5/10 | Probability estimated from concurrent-finding count (3). A higher count indicates multiple parallel activations of the cascade, increasing the likelihood that this specific 50-step pathogenic chain is operative in this patient. |
| **Detectability** | 10/10 | Terminal root node is at the quantum level. Current clinical diagnostics do not routinely detect pathology at this level before upstream clinical events manifest. |

**RPN 450 (HIGH):** RPN 300–499 — theoretical intervention at this root level is of high potential value; detectability improvements should be prioritized.

**Therapy Benchmarks**

**Fishbone (Ishikawa) Diagram**

**Effect:** Elevated troponin (6.4 ng/mL)

|  |  |
| --- | --- |
| **Patient Factors** | Age: 41null |
| **Disease / Diagnosis Factors** | DVT<br>asthma<br>depression<br>vasculitis<br>osteoporosis<br>deep vein thrombosis<br>gastrointestinal |
| **Therapeutic Factors** | amiodarone<br>digoxin<br>mepolizumab<br>benralizumab<br>rituximab |
| **Environmental / System Factors** | Care setting (inpatient/outpatient, unit type)<br>Institutional formulary availability<br>Access to specialty consultation<br>Local antibiogram or resistance patterns (if applicable) |
| **Measurement / Diagnostic Factors** | 4 laboratory value(s) documented<br>2 vital sign(s) documented<br>Diagnostic imaging modalities (to be verified from case)<br>Point-of-care testing availability |
| **Mechanism (from Polyroot Chain)** | Symptom level: Insufficient myocardial oxygen supply relative to demand<br>Organ-system dysfunction: Systemic inflammatory cytokine release<br>Tissue-level pathology: ATP synthase (F0F1) rotational catalysis<br>Cellular dysfunction: Frataxin protein expression and function<br>Organelle and membrane: Cyclophilin D conformational activation<br>Molecular biology: FBXL3-mediated CRY ubiquitination<br>Gene regulation and expression: G6PD enzymatic activity<br>Fundamental biochemistry: Interfacial water dynamics at protein surfaces<br>Electron transport and bioenergetics: p53 (TP53) tumor suppressor transcriptional activity |

**Run Chart — Actual vs Guideline Targets**

| Metric | Actual | Target | Status | Source |
| --- | --- | --- | --- | --- |
| Platelets | 340 K/uL | 150— | within target | Routine hematology reference |
| Troponin | 6.4 ng/mL | —0.04 | above target | Cardiac injury threshold (assay-dependent) |
| BNP | 8400 pg/mL | —400 | above target | Heart failure threshold (assay-dependent) |

**Pharmacogenomics Screen**

**Drug-Drug Interaction Screen**

1 interaction(s) detected: 0 contraindicated (X), 1 therapy-modification (D), 0 monitoring (C). Immediate attention required for X-severity pairs.

| Severity | Pair | Mechanism | Management |
| --- | --- | --- | --- |
| D | amiodarone + digoxin | P-gp inhibition: digoxin increases ~70% | reduce digoxin 50%, monitor level |

**Clinical Visualizations — Isobolograms & Nomograms**

**Combination-Effect Isobologram amiodarone + digoxin (and 0 other detected pair)**

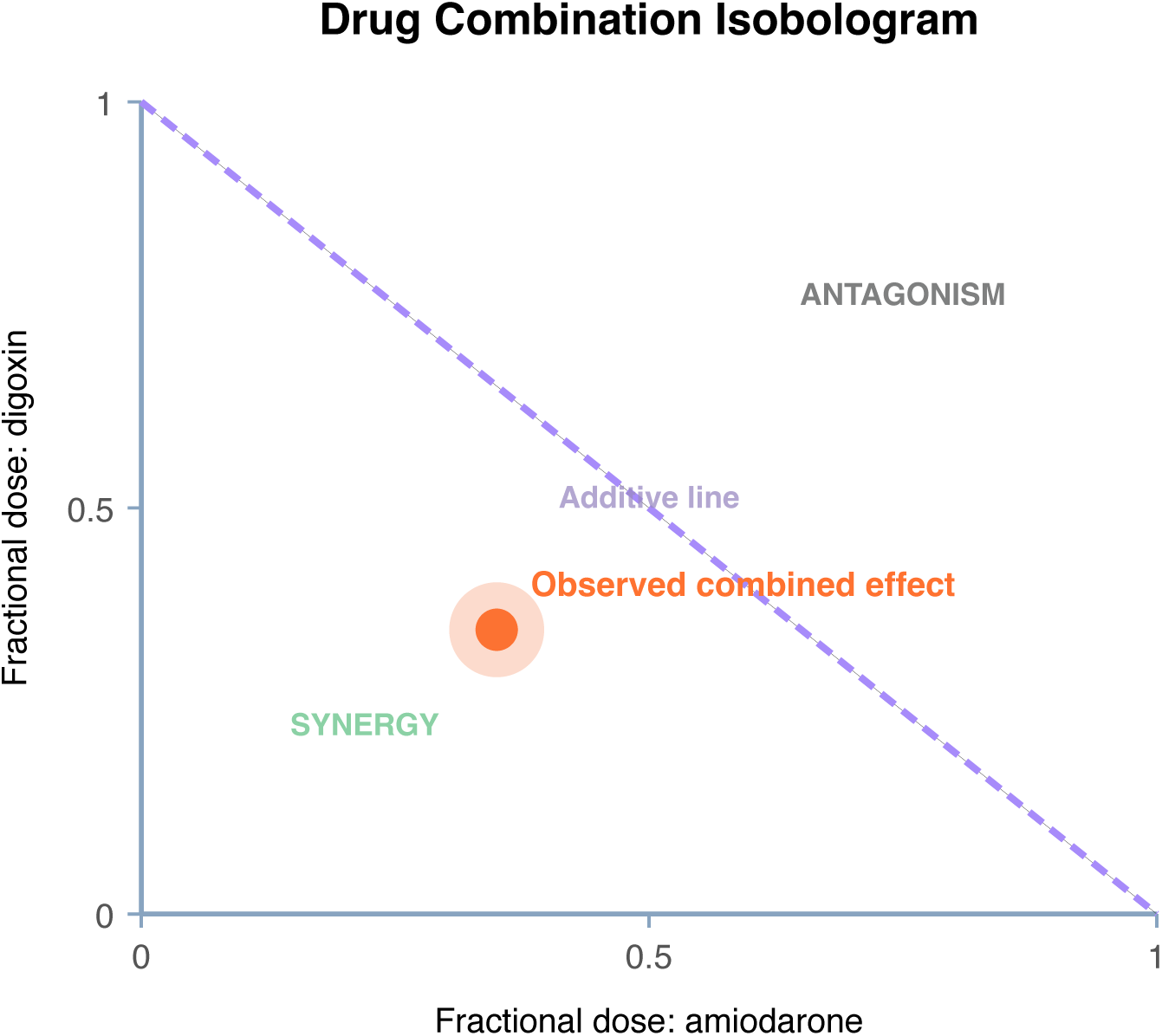

**Interpretation:** Modify-therapy combination: the combined effect exceeds additive prediction. Empirical point is below the Loewe additivity line, consistent with synergistic interaction.

**Clinical action:** reduce digoxin 50%, monitor level

**qSOFA Sepsis Screen**

Score 0 (mentation requires bedside assessment)

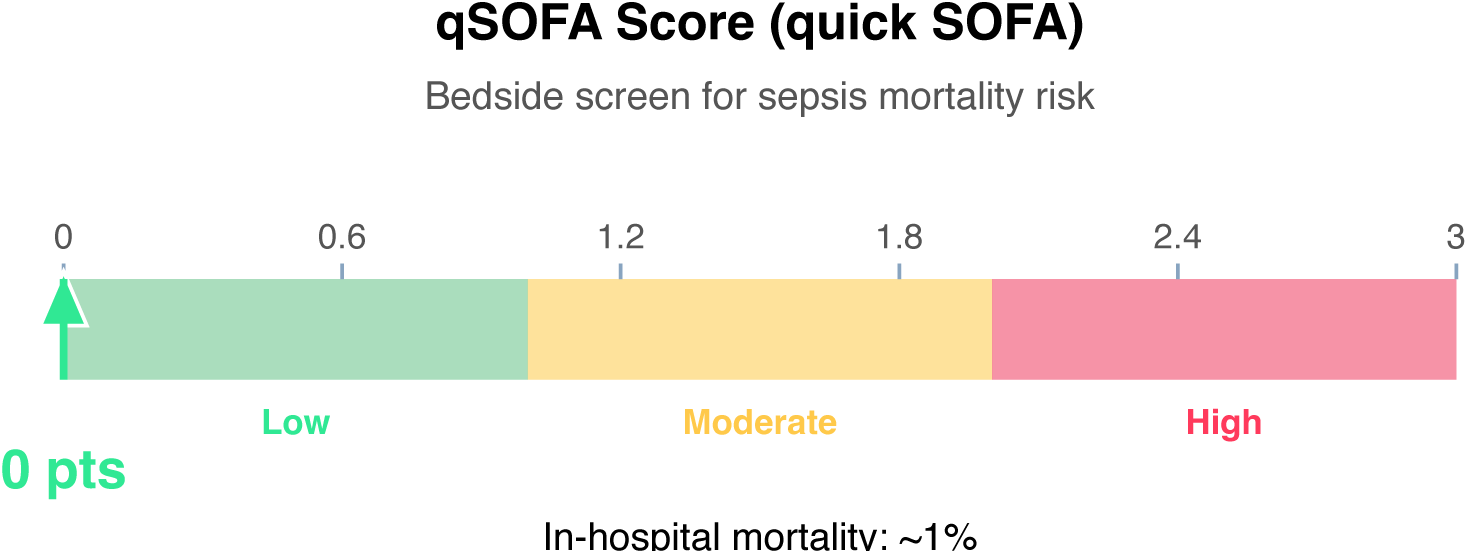

**Clinical action:** Continue monitoring; repeat if clinical deterioration.

Wells DVT Nomogram (structured-data subset)

Score 1 (Moderate probability)

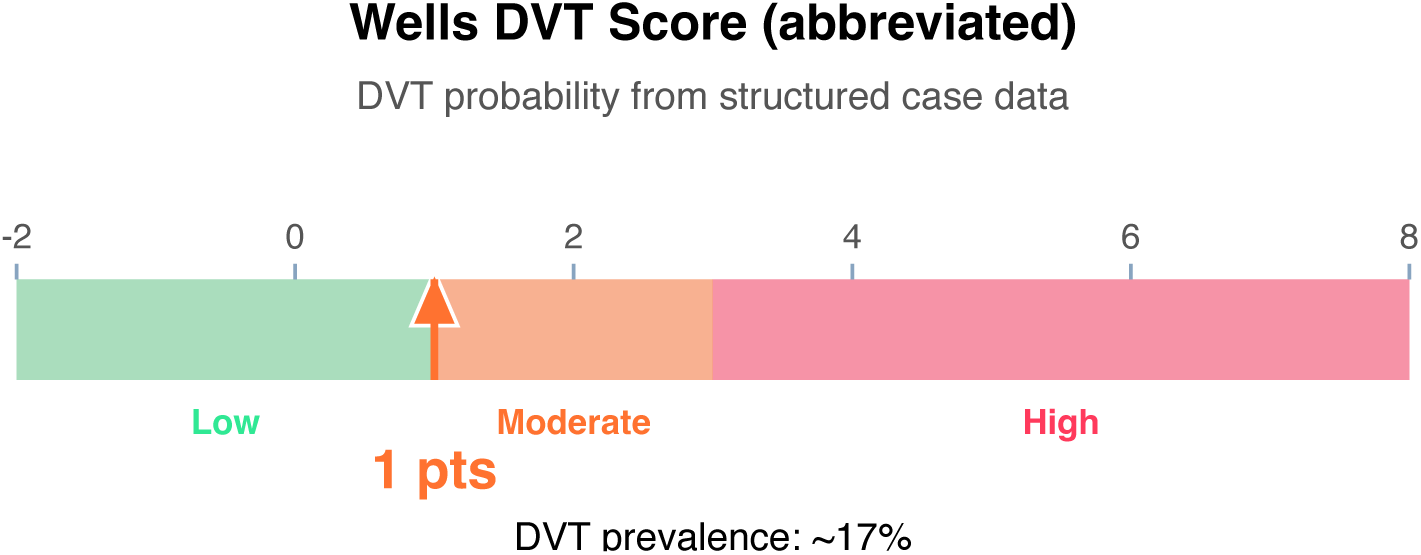

**Clinical action:** D-dimer; if positive, proceed to ultrasound.

**Article Structure (iteratively restructured)**

**Structure type:** narrative_review · **Converged:** true at pass 2/5 · **Quality:** 0.32

**Proposed outline:**

1. **Abstract** (0 clusters, 0 sources) Overview of the review scope and synthesis
2. **Background** (1 clusters, 0 sources) Historical context and evolution of understanding
3. **Current State of Evidence** (1 clusters, 1 sources) What is established vs uncertain
4. **Controversies and Debates** (0 clusters, 0 sources) Contradictory findings and interpretive disagreements
5. **Clinical Implications** (0 clusters, 0 sources) What the evidence means for practice
6. **Future Directions** (0 clusters, 0 sources) Knowledge gaps and research agenda
7. **Conclusion** (0 clusters, 0 sources) Synthesis in one paragraph
8. **References** (0 clusters, 9 sources) Complete citation list

**Thematic clusters:**

• Expert · Anti · Infect (1 statement)

**Structural gaps:**

- [MODERATE] empty section (Controversies and Debates) — No clusters or sources mapped to this section. Consider broadening the search to include targeted supplementary queries.
- [MODERATE] empty section (Clinical Implications) — No clusters or sources mapped to this section. Consider broadening the search to include clinical practice guidelines.
- [MODERATE] empty section (Future Directions) — No clusters or sources mapped to this section. Consider broadening the search to include targeted supplementary queries.
- [MODERATE] empty section (Conclusion) — No clusters or sources mapped to this section. Consider broadening the search to include targeted supplementary queries.
- [MODERATE] no quantitative data — No quantitative statements extracted (p-values, HRs, ORs, CIs). Consider searching PubMed for RCTs or meta-analyses on this topic.

**Quantitative Evidence Extraction**

**Cross-source summary:** Pooled N: n/a · 0/9 with p-values · 0/9 with CIs · 0/9 with effect sizes · 0 significant · 0 null

**GRADE Methodological Assessment**

| Domain | Concern | Rationale |
| --- | --- | --- |
| Risk of Bias | HIGH | Majority of sources show high risk of bias (incomplete methodological reporting or no adjustment for confounders). |
| Inconsistency | MODERATE | No heterogeneity statistics ($I^2$, $Q$, $\tau^2$) detected in retrieved text. Unable to formally assess inconsistency across 9 sources. |
| Indirectness | MODERATE | Partial alignment — 67% of sources reference case diagnoses. Some indirectness in population or condition. |
| Imprecision | LOW | Sample size, CI widths, and event counts do not raise precision concerns. |
| Publication Bias | HIGH | No funnel plot or Egger's/Begg's test detected in 9 sources; No trial registry or pre-registration verification detected. |

**Inferential Findings**

- **Elevated troponin (6.4 ng/mL) [LIMITED CONFIDENCE]**
- → Myocardial injury — rule out ACS; serial troponins and ECG indicated

• **Elevated BNP (8400 pg/mL) [LIMITED CONFIDENCE]**

→ Suggests volume overload / heart failure; echo review recommended

• **Fever (41°C) [LIMITED CONFIDENCE]**

→ Infection workup — blood cultures, imaging, source identification

**Recommended Verification Steps**

1. **[IMMEDIATE]** Serial cardiac enzymes, 12-lead ECG, and consider echocardiography *Rationale:* Elevated troponin (6.4 ng/mL): Myocardial injury — rule out ACS; serial troponins and ECG indicated *Resources:* ACC/AHA ACS guidelines, TIMI/GRACE risk calculators
2. **[URGENT]** Serial cardiac enzymes, 12-lead ECG, and consider echocardiography *Rationale:* Elevated BNP (8400 pg/mL): Suggests volume overload / heart failure; echo review recommended *Resources:* ACC/AHA ACS guidelines, TIMI/GRACE risk calculators
3. **[ROUTINE]** Verify management approach via specialty-specific resources — retrieved evidence base did not directly address this finding *Rationale:* Elevated troponin (6.4 ng/mL) *Resources:* UpToDate, DynaMed, Specialty society guidelines, Lexicomp/Micromedex, DailyMed for current product labeling
4. **[ROUTINE]** Verify management approach via specialty-specific resources — retrieved evidence base did not directly address this finding *Rationale:* Elevated BNP (8400 pg/mL) *Resources:* UpToDate, DynaMed, Specialty society guidelines, Lexicomp/Micromedex, DailyMed for current product labeling
5. **[ROUTINE]** Verify management approach via specialty-specific resources — retrieved evidence base did not directly address this finding *Rationale:* Fever (41°C) *Resources:* UpToDate, DynaMed, Specialty society guidelines, Lexicomp/Micromedex, DailyMed for current product labeling
6. **[ROUTINE]** Comprehensive medication reconciliation and interaction review for 5 agents *Rationale:* Polypharmacy (≥3 agents) warrants systematic DDI and appropriateness review *Resources:* Drugs.com Interaction Checker, Lexi-Interact, Beers Criteria (if age ≥65), STOPP/START criteria
7. **[ROUTINE]** Cross-verify all recommendations against institutional protocols, current product labeling (DailyMed), and specialty-specific guidelines before clinical application *Rationale:* AuditMed outputs are for research and educational use; clinical decisions remain the responsibility of the treating clinician *Resources:* Institutional clinical practice guidelines, DailyMed FDA labeling, Specialty society consensus documents

**Evidence-Based Recommendations**

- Address Elevated troponin (6.4 ng/mL) — Myocardial injury — rule out ACS; serial troponins and ECG indicated [Case-derived] **[HIGH]**
- Verify all therapeutic changes against current product labeling (DailyMed), institutional formulary, and patient-specific factors (allergies, organ function, goals of care) before implementation. **[N/A]**

**Strength of Evidence**

**GRADE:** LOW

*9 source(s) retrieved · 1 systematic review/meta-analysis · 3 limited-confidence finding(s) flagged for verification*

## Case 1: 35

**Type:** CASE 35

**File:** Library case #35 (pp. 36–36)

**Demographics:** 33

**Diagnoses:** DVT, CKD, depression, anemia, sepsis, chronic pain, hypertension, chronic kidney disease, deep vein thrombosis, gastrointestinal

**Labs documented:** 7

**Medications documented:** 3

**Clinical Question**

Given the working diagnoses of DVT, CKD, depression, current therapy including warfarin, ceftriaxone, ciprofloxacin, what does the retrieved evidence indicate about management, risks, and appropriate next steps?

**Evidence Summary**

Total sources: 9 · Guidelines: 0 · SR/MA: 0 · RCTs: 0 · Other: 9

**Reference Outline**

**Applicable guidelines:**

**Current research findings:**

**Reasoning Chain**

1. Case profile: 33 patient with working diagnoses of DVT, CKD, depression · 7 lab value(s), 3 medication(s) documented
2. Case-derived findings: 2 patterns identified (1 critical, 1 high-severity, 0 moderate/mild)
3. Evidence base: spans 2012–2023; 0 distinct methodology type(s) · 0 evidence-linked statement(s) extracted
4. Case findings did not align directly with retrieved evidence — external verification required
5. 2 finding(s) flagged for verification: evidence retrieved did not directly address these elements. Recommendations based on general clinical principles until verified against specialty resources.
6. Overall: evidence base insufficient for definitive inference; treat output as hypothesis-generating pending verification

**50-Why Polyroot Inference Chain**

**Seed:** Severe anemia (Hgb 5.8 g/dL) · **Terminal:** p53 phosphorylation (ATM/ATR/CHK1/CHK2) · **Iterations:**

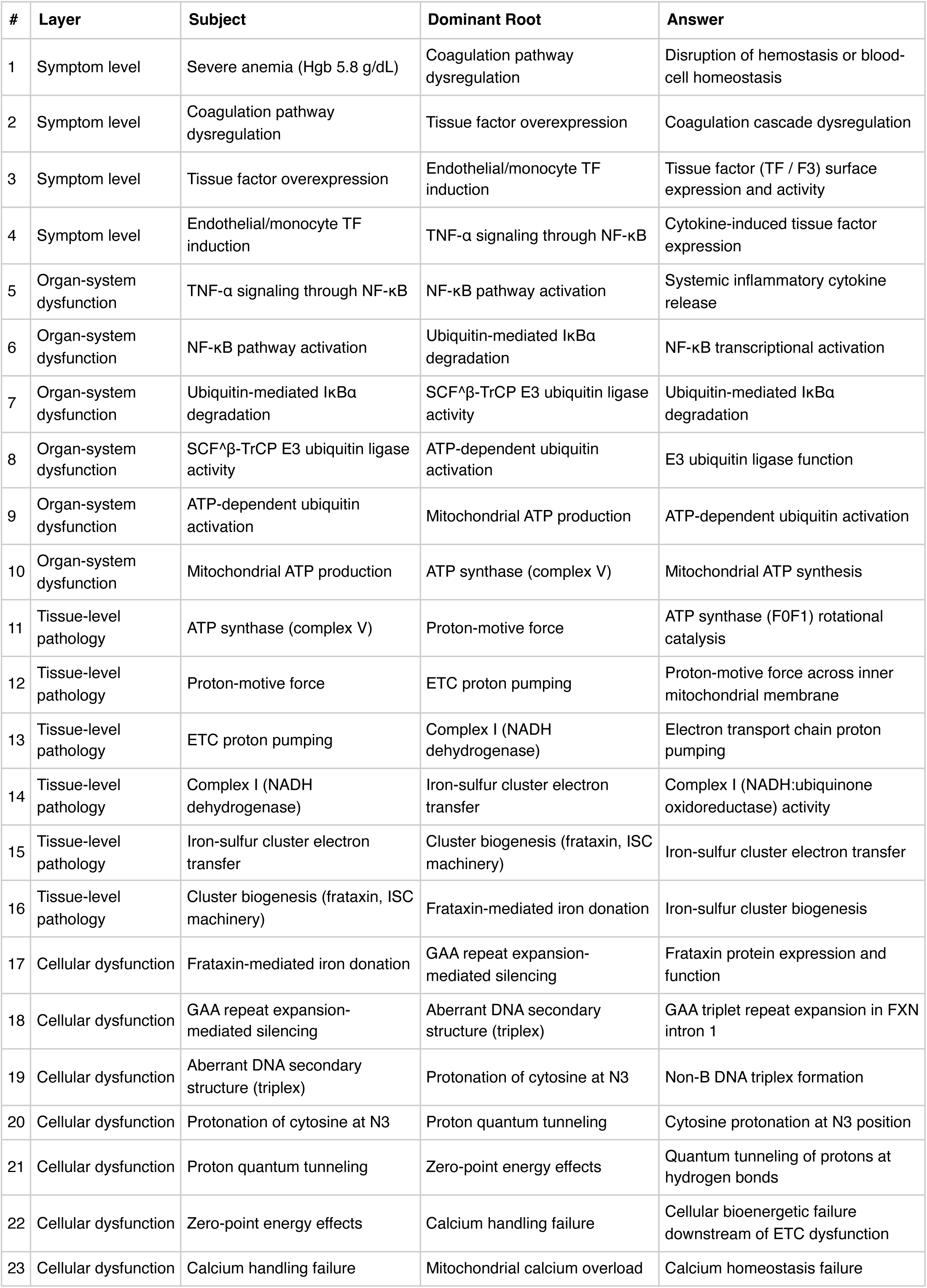
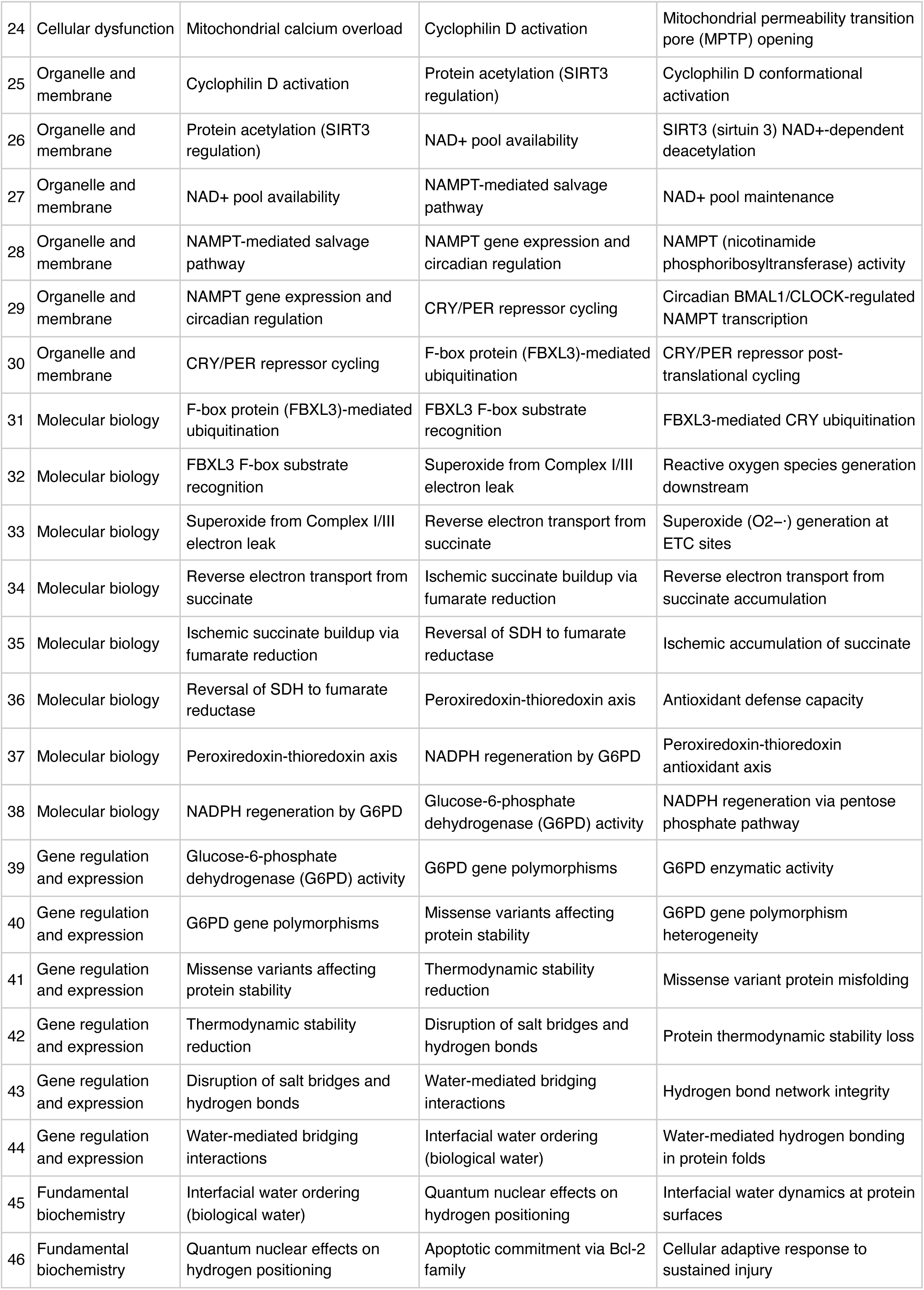
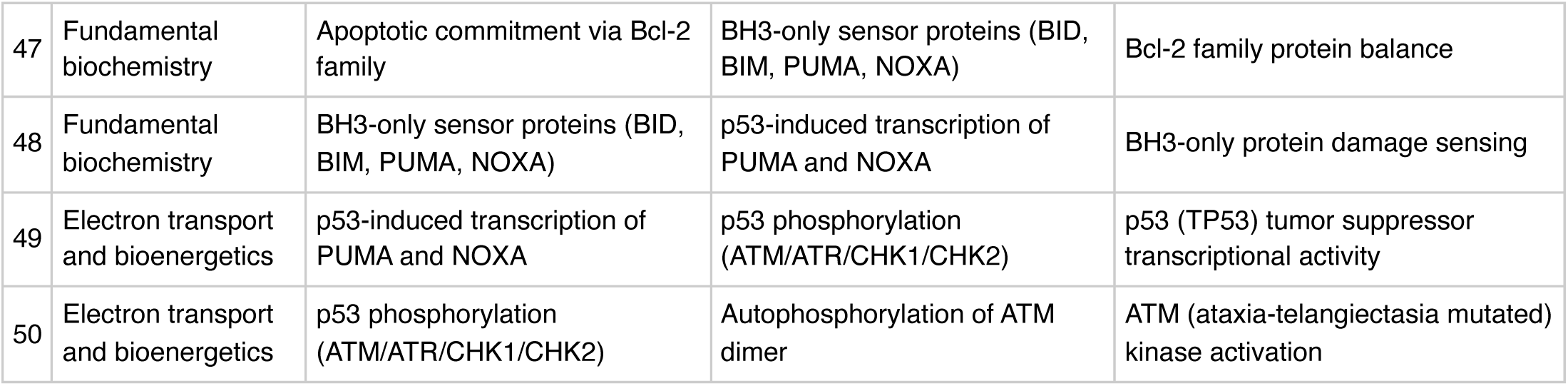

**FMEA Forecasting (Severity × Probability × Detectability)**

**Terminal node:** ATM (ataxia-telangiectasia mutated) kinase activation

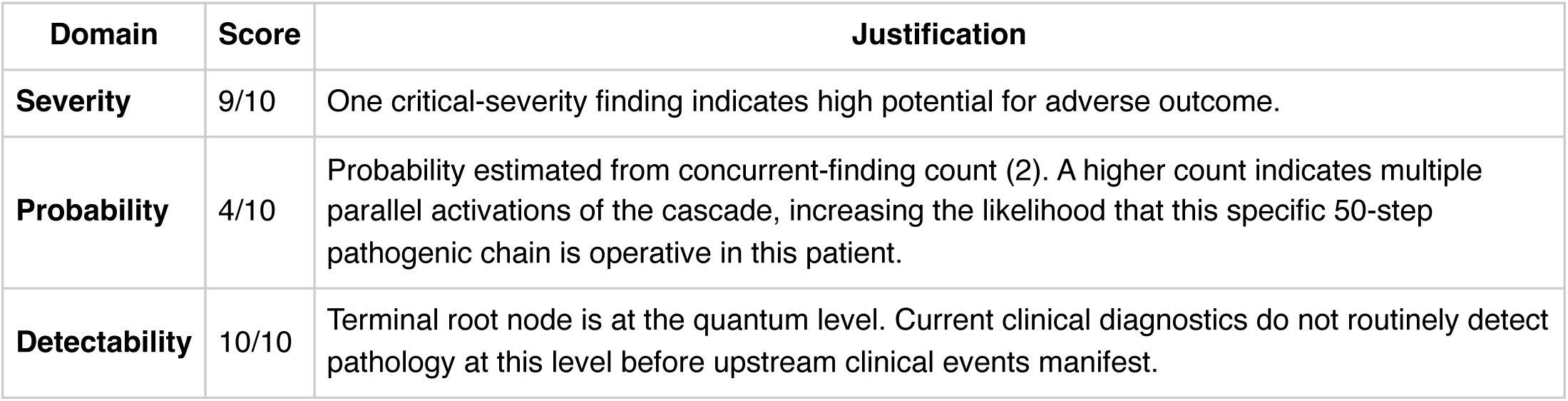

FMEA Forecasting (Severity × Probability × Detectability) **Terminal node:** ATM (ataxia-telangiectasia mutated) kinase activation
| Domain | Score | Justification |
| --- | --- | --- |
| Severity | 9/10 | One critical-severity finding indicates high potential for adverse outcome. |
| Probability | 4/10 | Probability estimated from concurrent-finding count (2). A higher count indicates multiple parallel activations of the cascade, increasing the likelihood that this specific 50-step pathogenic chain is operative in this patient. |
| Detectability | 10/10 | Terminal root node is at the quantum level. Current clinical diagnostics do not routinely detect pathology at this level before upstream clinical events manifest. |

**Therapy Benchmarks**

**Fishbone (Ishikawa) Diagram**

**Effect:** Severe anemia (Hgb 5.8 g/dL)

|  |  |
| --- | --- |
| **Patient Factors** | Age: 33null |
| **Disease / Diagnosis Factors** | DVT<br>CKD<br>depression<br>anemia<br>sepsis<br>chronic pain<br>hypertension<br>chronic kidney disease |
| **Therapeutic Factors** | warfarin<br>ceftriaxone<br>ciprofloxacin |
| **Environmental / System Factors** | Care setting (inpatient/outpatient, unit type)<br>Institutional formulary availability<br>Access to specialty consultation<br>Local antibiogram or resistance patterns (if applicable) |
| **Measurement / Diagnostic Factors** | 7 laboratory value(s) documented<br>2 vital sign(s) documented<br>Diagnostic imaging modalities (to be verified from case)<br>Point-of-care testing availability |
| **Mechanism (from Polyroot Chain)** | Symptom level: Disruption of hemostasis or blood-cell homeostasis<br>Organ-system dysfunction: Systemic inflammatory cytokine release<br>Tissue-level pathology: ATP synthase (F0F1) rotational catalysis<br>Cellular dysfunction: Frataxin protein expression and function<br>Organelle and membrane: Cyclophilin D conformational activation<br>Molecular biology: FBXL3-mediated CRY ubiquitination<br>Gene regulation and expression: G6PD enzymatic activity<br>Fundamental biochemistry: Interfacial water dynamics at protein surfaces<br>Electron transport and bioenergetics: p53 (TP53) tumor suppressor transcriptional activity |

**Run Chart — Actual vs Guideline Targets**

| Metric | Actual | Target | Status | Source |
| --- | --- | --- | --- | --- |
| Hemoglobin | 5.8 g/dL | 12— | below target | WHO anemia cut-offs |
| Platelets | 18 K/uL | 150— | below target | Routine hematology reference |
| INR | 2.8 | —3 | within target | Therapeutic range for most anticoagulation indications |
| ALT | 480 U/L | —40 | above target | Hepatocellular injury threshold |

**Pharmacogenomics Screen**

1 of 3 medications have documented pharmacogenomic actionability. 1 have CPIC level-A dosing guidelines available.

- **warfarin** (CPIC A)

- *CYP2C9 *2/*3 + VKORC1 −1639G>A:* reduce dose 25-50%, use validated algorithm
- *CYP2C9 *5/*6/*8/*11 (African ancestry):* increased bleeding risk PMID: 28198005

**Drug-Drug Interaction Screen**

No interactions detected in the embedded DDI matrix among the 3 documented medication(s). This does not exclude clinically significant interactions — verification via Lexi-Interact or Drugs.com Interaction Checker is still recommended for complex regimens.

**Clinical Visualizations — Isobolograms & Nomograms**

**Hepatotoxicity Combination Isobologram**

1 hepatotoxic agent: ceftriaxone · injury present

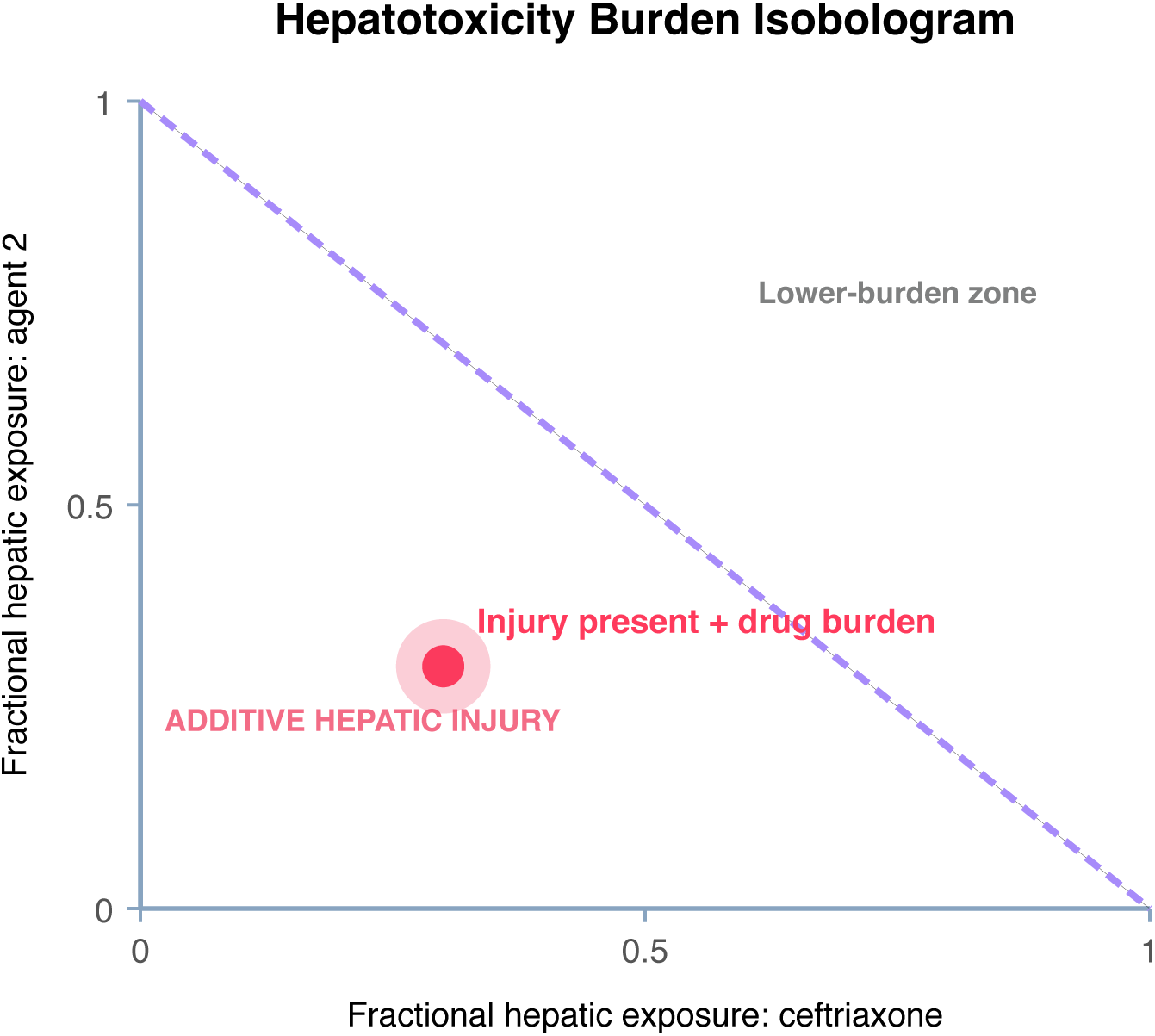

**Interpretation:** Existing hepatocellular or cholestatic injury (ALT 480, Tbili ?) combined with ongoing hepatotoxin exposure indicates ongoing or worsening DILI. Each additional hepatotoxin adds to cumulative injury risk.

**Warfarin INR Dosing Nomogram**

Therapeutic (VTE/AF range)

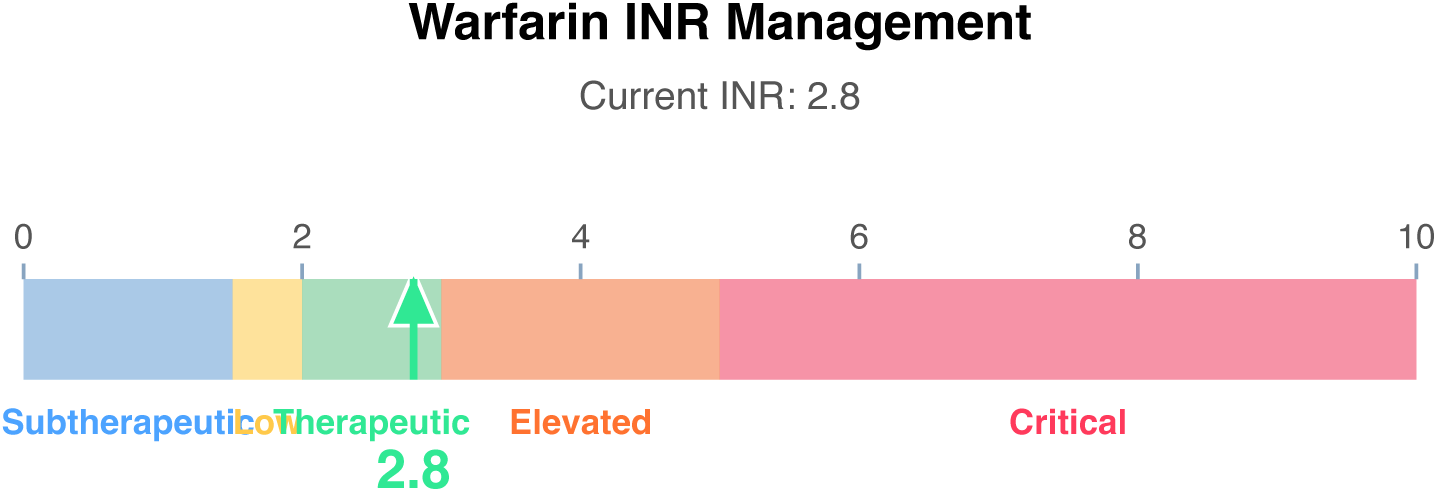

**Interpretation:** INR 2.8 — Therapeutic (VTE/AF range). Target range assumed 2-3 for VTE/AF; confirm target for patient-specific indication (mechanical valve targets may be 2.5-3.5 or 3.0-4.0).

**Clinical action:** No change; recheck per schedule

**qSOFA Sepsis Screen**

Score 0 (mentation requires bedside assessment)

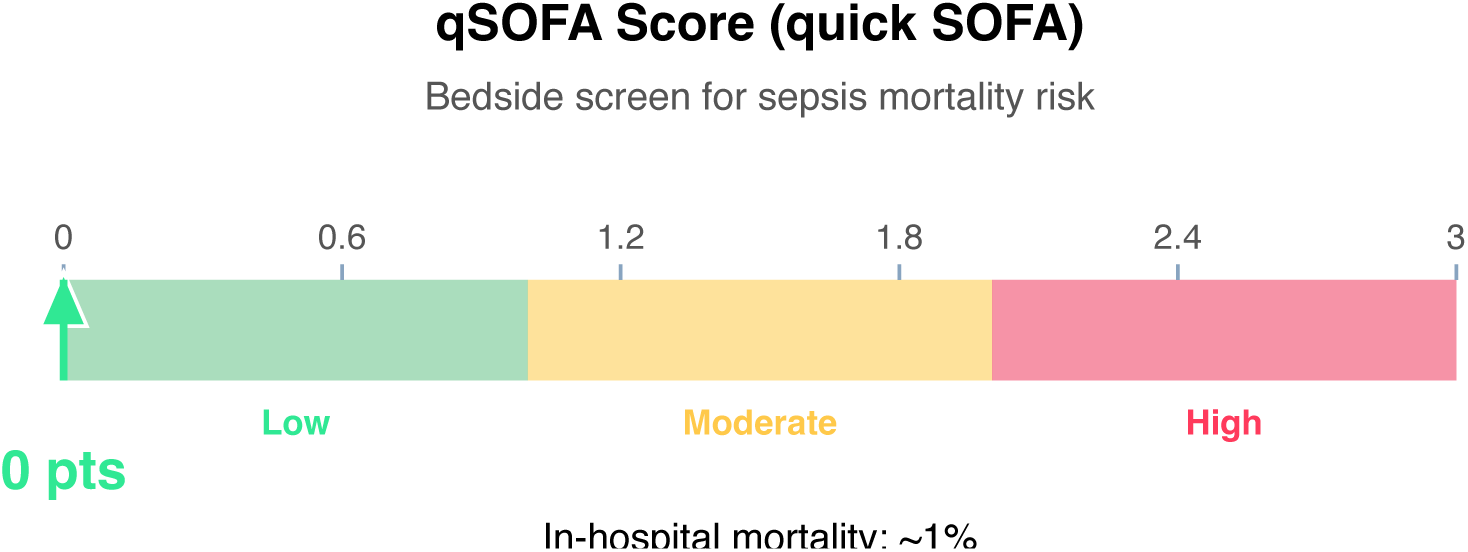

**Clinical action:** Continue monitoring; repeat if clinical deterioration.

**Wells DVT Nomogram (structured-data subset)**

Score 1 (Moderate probability)

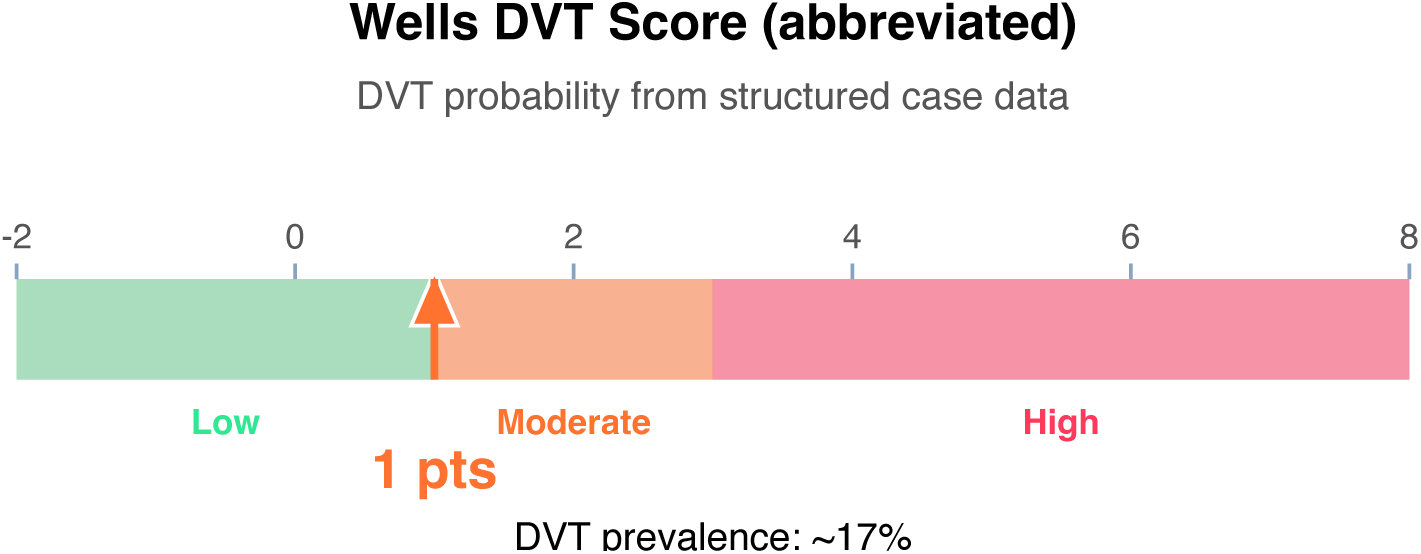

**Clinical action:** D-dimer; if positive, proceed to ultrasound.

**10-Year ASCVD Risk Nomogram (simplified)**

Estimated 10-year risk: 3% (Low)

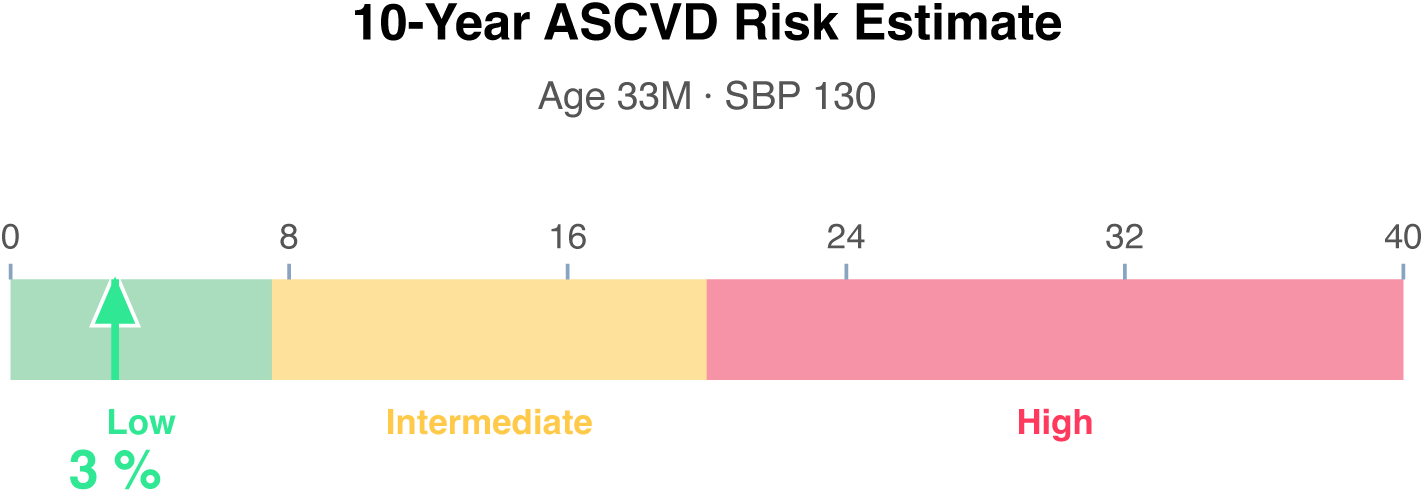

**Clinical action:** Lifestyle modification; reassess periodically.

**Article Structure (iteratively restructured)**

**Structure type:** narrative_review · **Converged:** true at pass 2/5 · **Quality:** 0.05

**Proposed outline:**

1. **Abstract** (0 clusters, 0 sources) Overview of the review scope and synthesis
2. **Background** (0 clusters, 0 sources) Historical context and evolution of understanding
3. **Current State of Evidence** (0 clusters, 0 sources) What is established vs uncertain
4. **Controversies and Debates** (0 clusters, 0 sources) Contradictory findings and interpretive disagreements
5. **Clinical Implications** (0 clusters, 0 sources) What the evidence means for practice
6. **Future Directions** (0 clusters, 0 sources) Knowledge gaps and research agenda
7. **Conclusion** (0 clusters, 0 sources) Synthesis in one paragraph
8. **References** (0 clusters, 9 sources) Complete citation list

**Structural gaps:**

- [MODERATE] empty section (Background) — No clusters or sources mapped to this section. Consider broadening the search to include review articles and mechanistic/pathophysiologic literature.
- [MODERATE] empty section (Current State of Evidence) — No clusters or sources mapped to this
- section. Consider broadening the search to include targeted supplementary queries.
- [MODERATE] empty section (Controversies and Debates) — No clusters or sources mapped to this section. Consider broadening the search to include targeted supplementary queries.
- [MODERATE] empty section (Clinical Implications) — No clusters or sources mapped to this section. Consider broadening the search to include clinical practice guidelines.
- [MODERATE] empty section (Future Directions) — No clusters or sources mapped to this section. Consider broadening the search to include targeted supplementary queries.
- [MODERATE] empty section (Conclusion) — No clusters or sources mapped to this section. Consider broadening the search to include targeted supplementary queries.
- [MODERATE] no quantitative data — No quantitative statements extracted (p-values, HRs, ORs, CIs). Consider searching PubMed for RCTs or meta-analyses on this topic.

**Quantitative Evidence Extraction**

**GRADE Methodological Assessment**

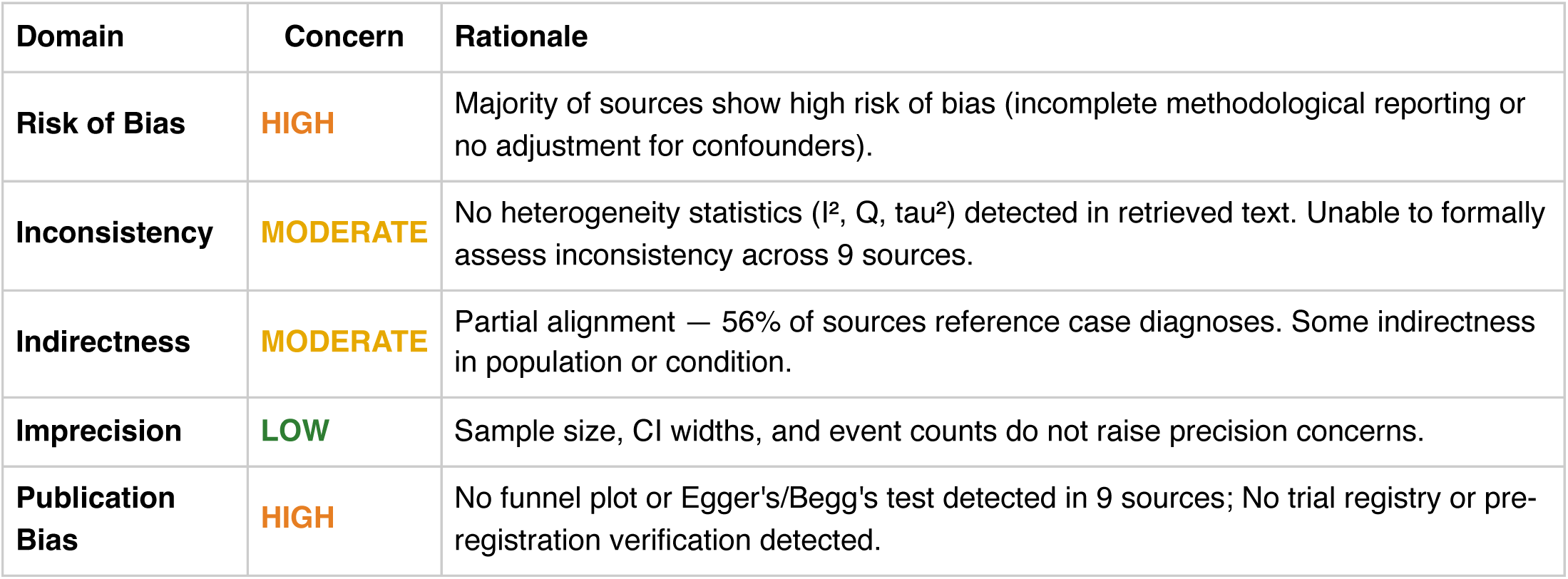

**Inferential Findings**

**• Severe anemia (Hgb 5.8 g/dL) [LIMITED CONFIDENCE]**

→ Transfusion threshold met in most settings; source evaluation needed

• **Severe thrombocytopenia (Plt 18 K/μL) [LIMITED CONFIDENCE]**

→ Bleeding risk; consider HIT if on heparin, medication-induced, hematologic process

**Recommended Verification Steps**

1. **[IMMEDIATE]** Repeat CBC and coagulation panel, type and cross if transfusion likely, source evaluation (GI workup, etc.) *Rationale:* Severe anemia (Hgb 5.8 g/dL): Transfusion threshold met in most settings; source evaluation needed *Resources:* Transfusion medicine guidelines (AABB), HIT 4T score if heparin exposure
2. **[URGENT]** Repeat CBC and coagulation panel, type and cross if transfusion likely, source evaluation (GI workup, etc.) *Rationale:* Severe thrombocytopenia (Plt 18 K/μL): Bleeding risk; consider HIT if on heparin, medication-induced, hematologic process *Resources:* Transfusion medicine guidelines (AABB), HIT 4T score if heparin exposure
3. **[ROUTINE]** Verify management approach via specialty-specific resources — retrieved evidence base did not directly address this finding *Rationale:* Severe anemia (Hgb 5.8 g/dL) *Resources:* UpToDate, DynaMed, Specialty society guidelines, Lexicomp/Micromedex, DailyMed for current product labeling
4. **[ROUTINE]** Verify management approach via specialty-specific resources — retrieved evidence base did not directly address this finding *Rationale:* Severe thrombocytopenia (Plt 18 K/μL) *Resources:* UpToDate, DynaMed, Specialty society guidelines, Lexicomp/Micromedex, DailyMed for current product labeling
5. **[ROUTINE]** Comprehensive medication reconciliation and interaction review for 3 agents *Rationale:* Polypharmacy (≥3 agents) warrants systematic DDI and appropriateness review *Resources:* Drugs.com Interaction Checker, Lexi-Interact, Beers Criteria (if age ≥65), STOPP/START criteria
6. **[ROUTINE]** Cross-verify all recommendations against institutional protocols, current product labeling (DailyMed), and specialty-specific guidelines before clinical application *Rationale:* AuditMed outputs are for research and educational use; clinical decisions remain the responsibility of the treating clinician *Resources:* Institutional clinical practice guidelines, DailyMed FDA labeling, Specialty society consensus documents

**Evidence-Based Recommendations**

- Address Severe anemia (Hgb 5.8 g/dL) — Transfusion threshold met in most settings; source evaluation needed [Case-derived] [HIGH]
- Verify all therapeutic changes against current product labeling (DailyMed), institutional formulary, and patient-specific factors (allergies, organ function, goals of care) before implementation. **[N/A]**

**Strength of Evidence**

**GRADE:** LOW

*9 source(s) retrieved · 2 limited-confidence finding(s) flagged for verification*

## Case 1: 36

**Type:** CASE 36

**File:** Library case #36 (pp. 37–37)

**Demographics:** 20

**Diagnoses:** deficiency [B12], B12 [B12], AKI, depression, anemia, B12 deficiency, ALL, bacteremia, aspergillosis, acute kidney injury, gastrointestinal, acute lymphoblastic leukemia

**Labs documented:** 5

**Medications documented:** 5

**Clinical Question**

Given the working diagnoses of deficiency [B12], B12 [B12], AKI, current therapy including meropenem, tobramycin, voriconazole, what does the retrieved evidence indicate about management, risks, and appropriate next steps?

**Evidence Summary**

Total sources: 11 · Guidelines: 0 · SR/MA: 0 · RCTs: 0 · Other: 11

Reference Outline

**Applicable guidelines:**

• KDIGO — Kidney Disease (https://kdigo.org/guidelines/)

**Historical precedent:** Trajectory of understanding for deficiency [B12] has shifted from symptom-based empirical management toward mechanism-targeted therapy informed by receptor-level, genetic, and molecular-pathway data. Recent decades have emphasized biomarker-guided dosing, shared decision-making, and cumulative-harm minimization across comorbid conditions.

**Current research findings:**

**Reasoning Chain**

1. Case profile: 20 patient with working diagnoses of deficiency [B12], B12 [B12], AKI · 5 lab value(s), 5 medication(s) documented
2. Case-derived findings: 3 patterns identified (1 critical, 2 high-severity, 0 moderate/mild)
3. Evidence base: spans 1978–2025; 0 distinct methodology type(s) · 0 evidence-linked statement(s) extracted
4. Case findings did not align directly with retrieved evidence — external verification required
5. 3 finding(s) flagged for verification: evidence retrieved did not directly address these elements. Recommendations based on general clinical principles until verified against specialty resources.
6. Overall: evidence base insufficient for definitive inference; treat output as hypothesis-generating pending verification

**50-Why Polyroot Inference Chain**

**Seed:** Severe anemia (Hgb 4.2 g/dL) · **Terminal:** p53 phosphorylation (ATM/ATR/CHK1/CHK2) · **Iterations:** 50

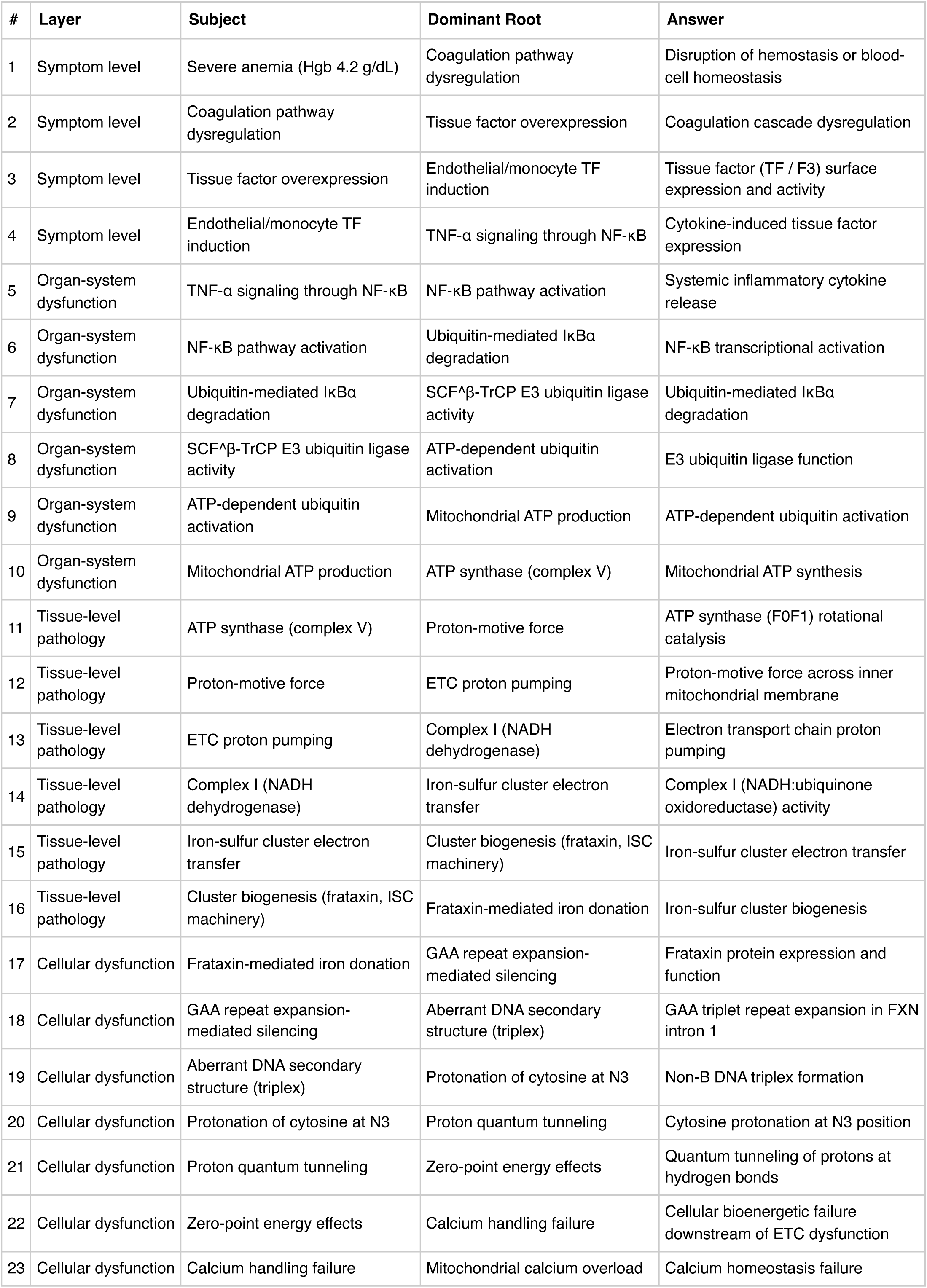
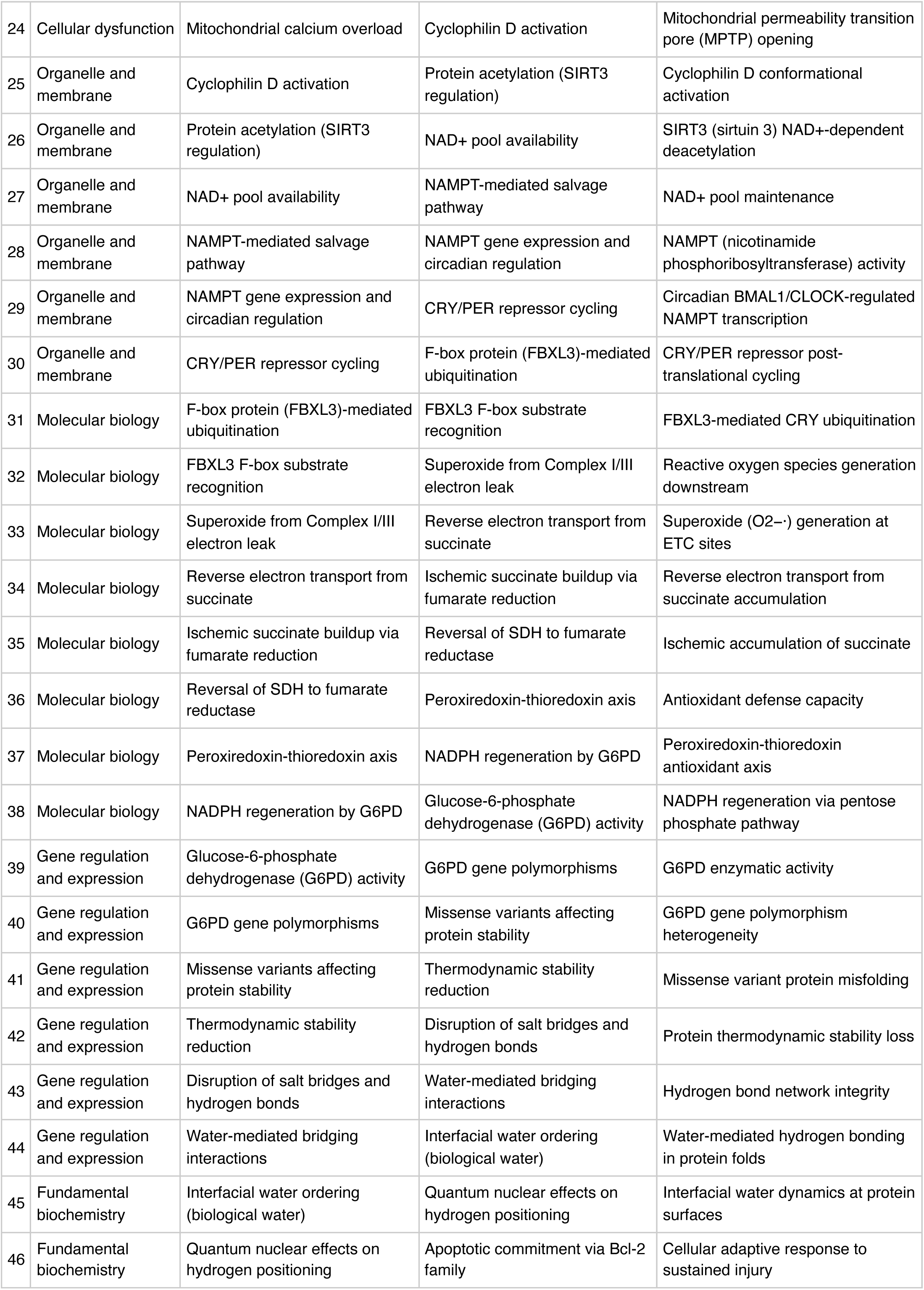
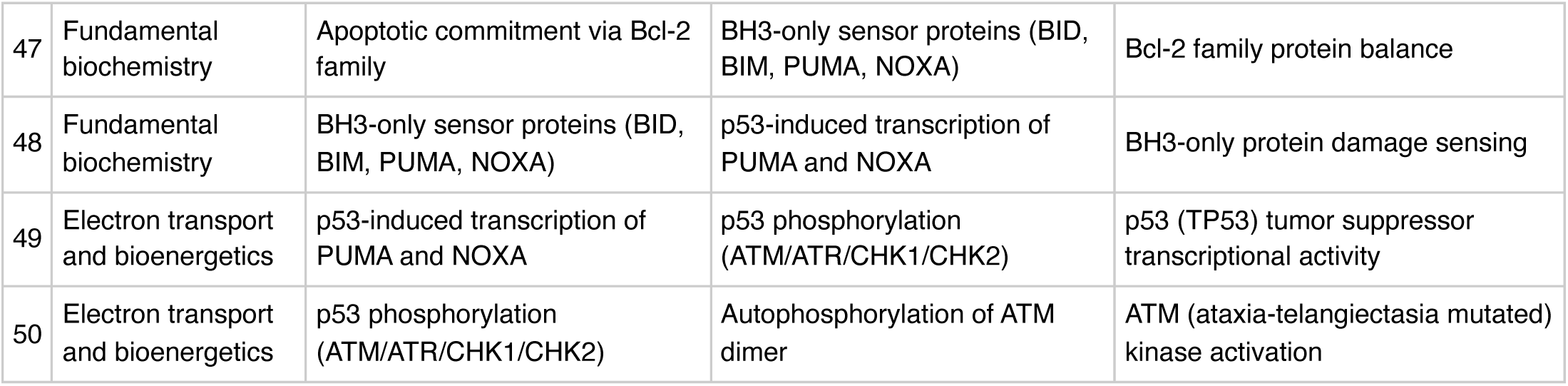

**FMEA Forecasting (Severity × Probability × Detectability)**

**Terminal node:** ATM (ataxia-telangiectasia mutated) kinase activation

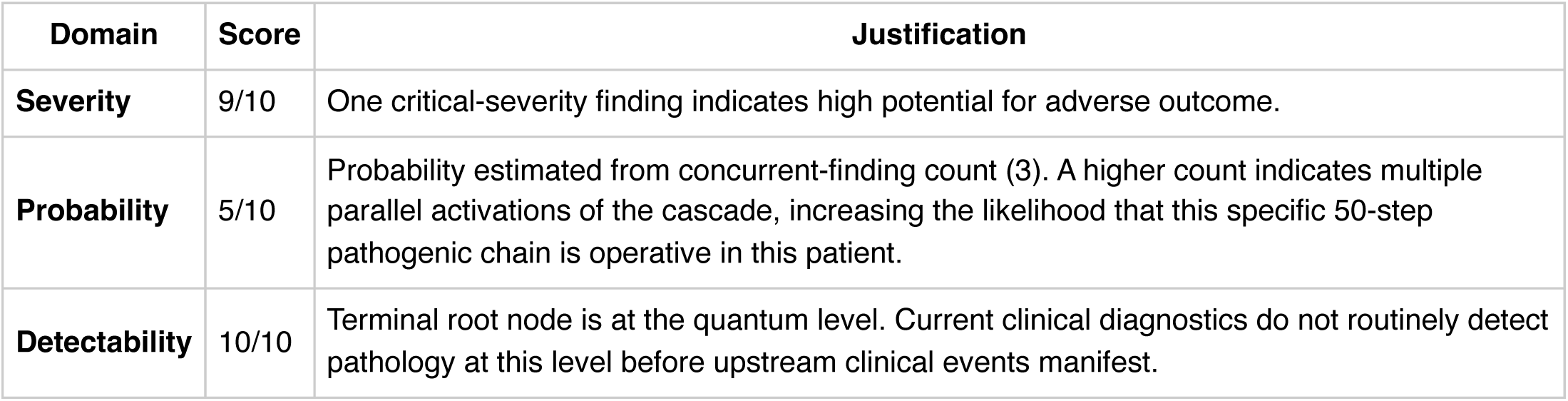

**Therapy Benchmarks**

**Therapeutic delta:** Deep-root interventions, if tractable, would theoretically produce NNT reductions greater than any single pillar of current GDMT because they act upstream of multiple downstream manifestations. However, most root-node targets at the molecular or genetic level are not currently druggable, so the theoretical benefit cannot yet be realized in clinical practice.

**Fishbone (Ishikawa) Diagram**

**Effect:** Elevated BNP (4200 pg/mL)

|  |  |
| --- | --- |
| **Patient Factors** | Age: 20null |
| **Disease / Diagnosis Factors** | deficiency [B12]<br>B12 [B12]<br>AKI<br>depression<br>anemia<br>B12 deficiency<br>ALL<br>bacteremia |
| **Therapeutic Factors** | meropenem<br>tobramycin<br>voriconazole<br>amphotericin<br>cyclosporine |
| **Environmental / System Factors** | Care setting (inpatient/outpatient, unit type)<br>Institutional formulary availability<br>Access to specialty consultation<br>Local antibiogram or resistance patterns (if applicable) |
| **Measurement / Diagnostic Factors** | 5 laboratory value(s) documented<br>3 vital sign(s) documented<br>Diagnostic imaging modalities (to be verified from case)<br>Point-of-care testing availability |
| **Mechanism (from Polyroot Chain)** | Symptom level: Disruption of hemostasis or blood-cell homeostasis<br>Organ-system dysfunction: Systemic inflammatory cytokine release<br>Tissue-level pathology: ATP synthase (F0F1) rotational catalysis<br>Cellular dysfunction: Frataxin protein expression and function<br>Organelle and membrane: Cyclophilin D conformational activation<br>Molecular biology: FBXL3-mediated CRY ubiquitination<br>Gene regulation and expression: G6PD enzymatic activity<br>Fundamental biochemistry: Interfacial water dynamics at protein surfaces<br>Electron transport and bioenergetics: p53 (TP53) tumor suppressor transcriptional activity |

**Run Chart — Actual vs Guideline Targets**

| Metric | Actual | Target | Status | Source |
| --- | --- | --- | --- | --- |
| Hemoglobin | 4.2 g/dL | 12— | below target | WHO anemia cut-offs |
| Platelets | 4 K/uL | 150— | below target | Routine hematology reference |
| BNP | 4200 pg/mL | —400 | above target | Heart failure threshold (assay-dependent) |

**Pharmacogenomics Screen**

1 of 5 medications have documented pharmacogenomic actionability. 1 have CPIC level-A dosing guidelines available.

- **voriconazole** (CPIC A)

- *CYP2C19 PM:* use alternative azole (posaconazole, isavuconazole)
- *CYP2C19 UM (*17/*17):* subtherapeutic — consider alternative PMID: 27981572

**Drug-Drug Interaction Screen**

No interactions detected in the embedded DDI matrix among the 5 documented medication(s). This does not exclude clinically significant interactions — verification via Lexi-Interact or Drugs.com Interaction Checker is still recommended for complex regimens.

**Clinical Visualizations — Isobolograms & Nomograms**

**qSOFA Sepsis Screen**

Score 0 (mentation requires bedside assessment)

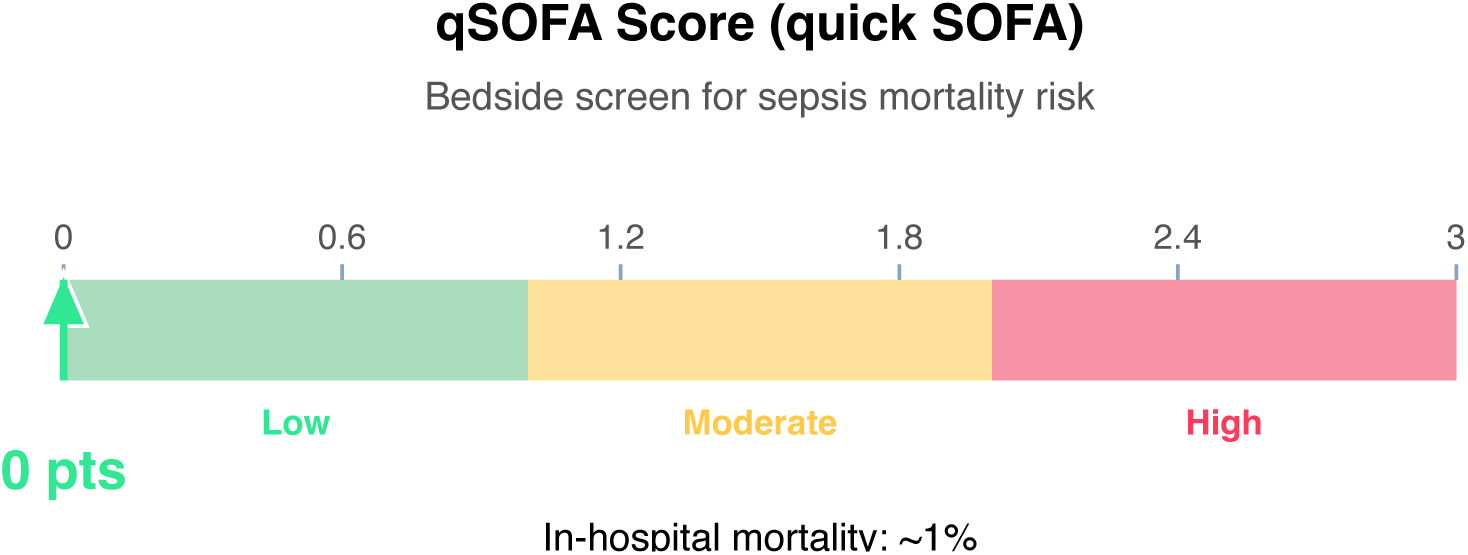

**Clinical action:** Continue monitoring; repeat if clinical deterioration.

**Article Structure (iteratively restructured)**

**Structure type:** narrative_review · **Converged:** true at pass 2/5 · **Quality:** 0.05

**Proposed outline:**

1. **Abstract** (0 clusters, 0 sources) Overview of the review scope and synthesis
2. **Background** (0 clusters, 0 sources) Historical context and evolution of understanding
3. **Current State of Evidence** (0 clusters, 0 sources) What is established vs uncertain
4. **Controversies and Debates** (0 clusters, 0 sources) Contradictory findings and interpretive disagreements
5. **Clinical Implications** (0 clusters, 0 sources) What the evidence means for practice
6. **Future Directions** (0 clusters, 0 sources) Knowledge gaps and research agenda
7. **Conclusion** (0 clusters, 0 sources) Synthesis in one paragraph
8. **References** (0 clusters, 11 sources) Complete citation list

**Structural gaps:**

- [MODERATE] empty section (Background) — No clusters or sources mapped to this section. Consider broadening the search to include review articles and mechanistic/pathophysiologic literature.
- [MODERATE] empty section (Current State of Evidence) — No clusters or sources mapped to this section. Consider broadening the search to include targeted supplementary queries.
- [MODERATE] empty section (Controversies and Debates) — No clusters or sources mapped to this section. Consider broadening the search to include targeted supplementary queries.
- [MODERATE] empty section (Clinical Implications) — No clusters or sources mapped to this section. Consider broadening the search to include clinical practice guidelines.
- [MODERATE] empty section (Future Directions) — No clusters or sources mapped to this section. Consider broadening the search to include targeted supplementary queries.
- [MODERATE] empty section (Conclusion) — No clusters or sources mapped to this section. Consider broadening the search to include targeted supplementary queries.
- & [MODERATE] no quantitative data — No quantitative statements extracted (p-values, HRs, ORs, CIs). Consider searching PubMed for RCTs or meta-analyses on this topic.

**Quantitative Evidence Extraction**

**Cross-source summary:** Pooled N: n/a · 0/11 with p-values · 0/11 with CIs · 0/11 with effect sizes · 0 significant · 0 null

**GRADE Methodological Assessment**

| Domain | Concern | Rationale |
| --- | --- | --- |
| Risk of Bias | HIGH | Majority of sources show high risk of bias (incomplete methodological reporting or no adjustment for confounders). |
| Inconsistency | MODERATE | No heterogeneity statistics ($I^2$, $Q$, $\tau^2$) detected in retrieved text. Unable to formally assess inconsistency across 11 sources. |
| Indirectness | LOW | Most retrieved sources (82%) reference the case diagnoses directly. Population and condition appear well-aligned. |
| Imprecision | LOW | Sample size, CI widths, and event counts do not raise precision concerns. |
| Publication Bias | HIGH | No funnel plot or Egger's/Begg's test detected in 11 sources; No trial registry or pre-registration verification detected. |

**Inferential Findings**

**• Elevated BNP (4200 pg/mL) [LIMITED CONFIDENCE]**

→ Suggests volume overload / heart failure; echo review recommended

**• Severe anemia (Hgb 4.2 g/dL) [LIMITED CONFIDENCE]**

→ Transfusion threshold met in most settings; source evaluation needed

**• Severe thrombocytopenia (Plt 4 K/μL) [LIMITED CONFIDENCE]**

→ Bleeding risk; consider HIT if on heparin, medication-induced, hematologic process

**Recommended Verification Steps**

1. **[URGENT]** Serial cardiac enzymes, 12-lead ECG, and consider echocardiography *Rationale:* Elevated BNP (4200 pg/mL): Suggests volume overload / heart failure; echo review recommended *Resources:* ACC/AHA ACS guidelines, TIMI/GRACE risk calculators
2. **[IMMEDIATE]** Repeat CBC and coagulation panel, type and cross if transfusion likely, source evaluation (GI workup, etc.) *Rationale:* Severe anemia (Hgb 4.2 g/dL): Transfusion threshold met in most settings; source evaluation needed *Resources:* Transfusion medicine guidelines (AABB), HIT 4T score if heparin exposure
3. **[URGENT]** Repeat CBC and coagulation panel, type and cross if transfusion likely, source evaluation (GI workup, etc.) *Rationale:* Severe thrombocytopenia (Plt 4 K/μL): Bleeding risk; consider HIT if on heparin, medication-induced, hematologic process *Resources:* Transfusion medicine guidelines (AABB), HIT 4T score if heparin exposure
4. **[ROUTINE]** Verify management approach via specialty-specific resources — retrieved evidence base did not directly address this finding *Rationale:* Elevated BNP (4200 pg/mL) *Resources:* UpToDate, DynaMed, Specialty society guidelines, Lexicomp/Micromedex, DailyMed for current product labeling
5. **[ROUTINE]** Verify management approach via specialty-specific resources — retrieved evidence base did not directly address this finding *Rationale:* Severe anemia (Hgb 4.2 g/dL) *Resources:* UpToDate, DynaMed, Specialty society guidelines, Lexicomp/Micromedex, DailyMed for current product labeling
6. **[ROUTINE]** Verify management approach via specialty-specific resources — retrieved evidence base did not directly address this finding *Rationale:* Severe thrombocytopenia (Plt 4 K/μL) *Resources:* UpToDate, DynaMed, Specialty society guidelines, Lexicomp/Micromedex, DailyMed for current product labeling
7. **[ROUTINE]** Comprehensive medication reconciliation and interaction review for 5 agents *Rationale:* Polypharmacy (≥3 agents) warrants systematic DDI and appropriateness review *Resources:* Drugs.com Interaction Checker, Lexi-Interact, Beers Criteria (if age ≥65), STOPP/START criteria
8. **[ROUTINE]** Cross-verify all recommendations against institutional protocols, current product labeling (DailyMed), and specialty-specific guidelines before clinical application *Rationale:* AuditMed outputs are for research and educational use; clinical decisions remain the responsibility of the treating clinician *Resources:* Institutional clinical practice guidelines, DailyMed FDA labeling, Specialty society consensus documents

**Evidence-Based Recommendations**

- Address Severe anemia (Hgb 4.2 g/dL) — Transfusion threshold met in most settings; source evaluation needed [Case-derived] **[HIGH]**
- Verify all therapeutic changes against current product labeling (DailyMed), institutional formulary, and patient-specific factors (allergies, organ function, goals of care) before implementation. **[N/A]**

**Strength of Evidence**

**GRADE:** LOW

*11 source(s) retrieved · 3 limited-confidence finding(s) flagged for verification*

## Case 1: 37

**Type:** CASE 37

**File:** Library case #37 (pp. 38–38)

**Demographics:** 63

**Diagnoses:** pancreatitis, depression, anemia, hypothyroidism, diabetes mellitus, gastrointestinal

**Labs documented:** 6

**Medications documented:** 3

**Clinical Question**

Given the working diagnoses of pancreatitis, depression, anemia, current therapy including prednisone, rituximab, levothyroxine, what does the retrieved evidence indicate about management, risks, and appropriate next steps?

**Evidence Summary**

Total sources: 12 · Guidelines: 0 · SR/MA: 0 · RCTs: 0 · Other: 12

**Reference Outline**

**Applicable guidelines:**

• APA — Psychiatry (https://www.psychiatry.org/psychiatrists/practice/clinical-practice-guidelines)

**Historical precedent:** Trajectory of understanding for pancreatitis has shifted from symptom-based empirical management toward mechanism-targeted therapy informed by receptor-level, genetic, and molecular-pathway data. Recent decades have emphasized biomarker-guided dosing, shared decision-making, and cumulative-harm minimization across comorbid conditions.

**Current research findings:**

**Reasoning Chain**

1. Case profile: 63 patient with working diagnoses of pancreatitis, depression, anemia · 6 lab value(s), 3 medication(s) documented
2. Case-derived findings: 1 pattern identified (1 moderate/mild)
3. Evidence base: spans 1978–2025; 0 distinct methodology type(s) · 1 evidence-linked statement(s) extracted
4. Case findings did not align directly with retrieved evidence — external verification required
5. 1 finding(s) flagged for verification: evidence retrieved did not directly address these elements. Recommendations based on general clinical principles until verified against specialty resources.
6. Overall: evidence base insufficient for definitive inference; treat output as hypothesis-generating pending verification

**50-Why Polyroot Inference Chain**

**Seed:** Fever (63°C) · **Terminal:** p53 phosphorylation (ATM/ATR/CHK1/CHK2) · **Iterations:** 50

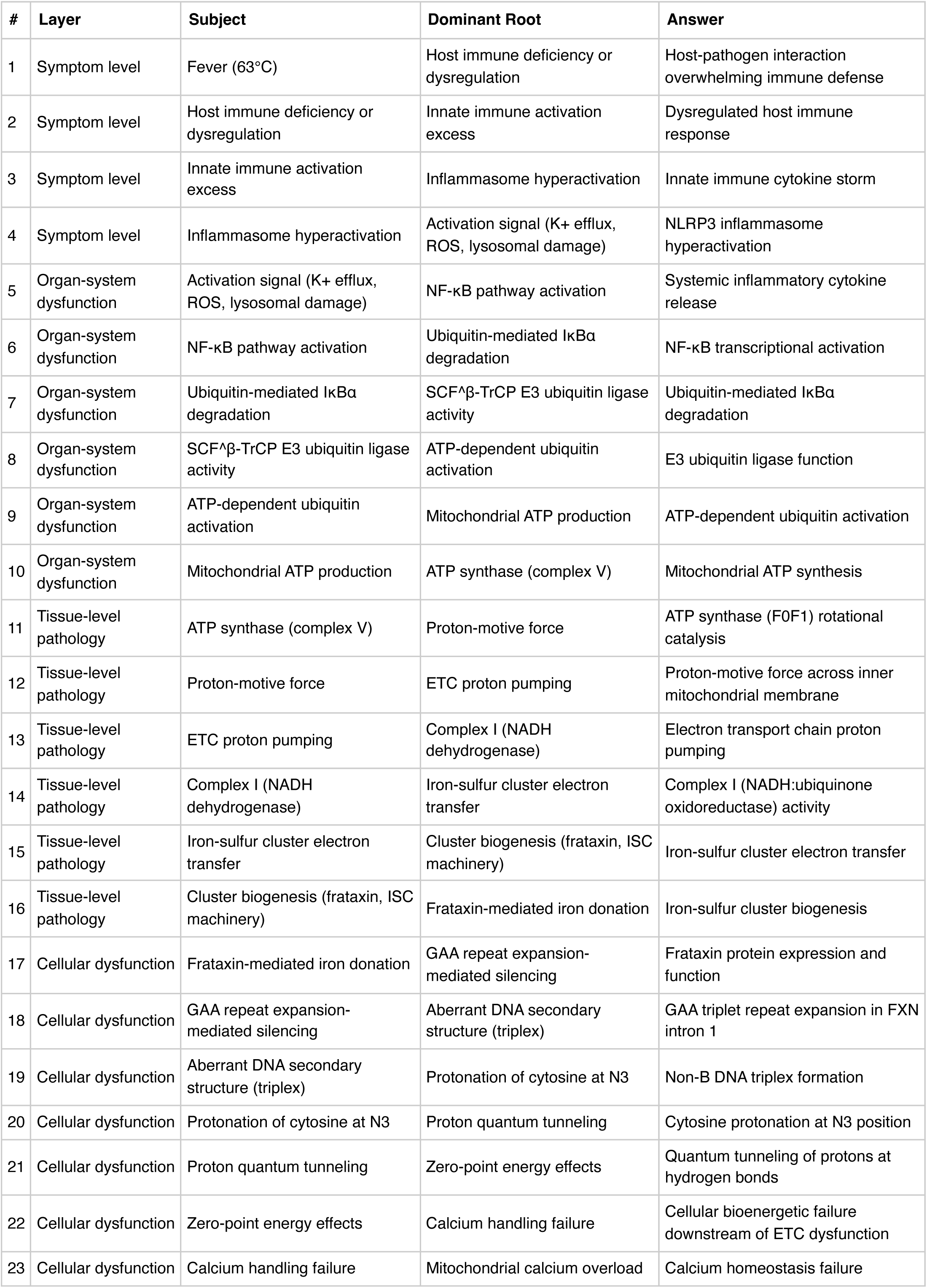
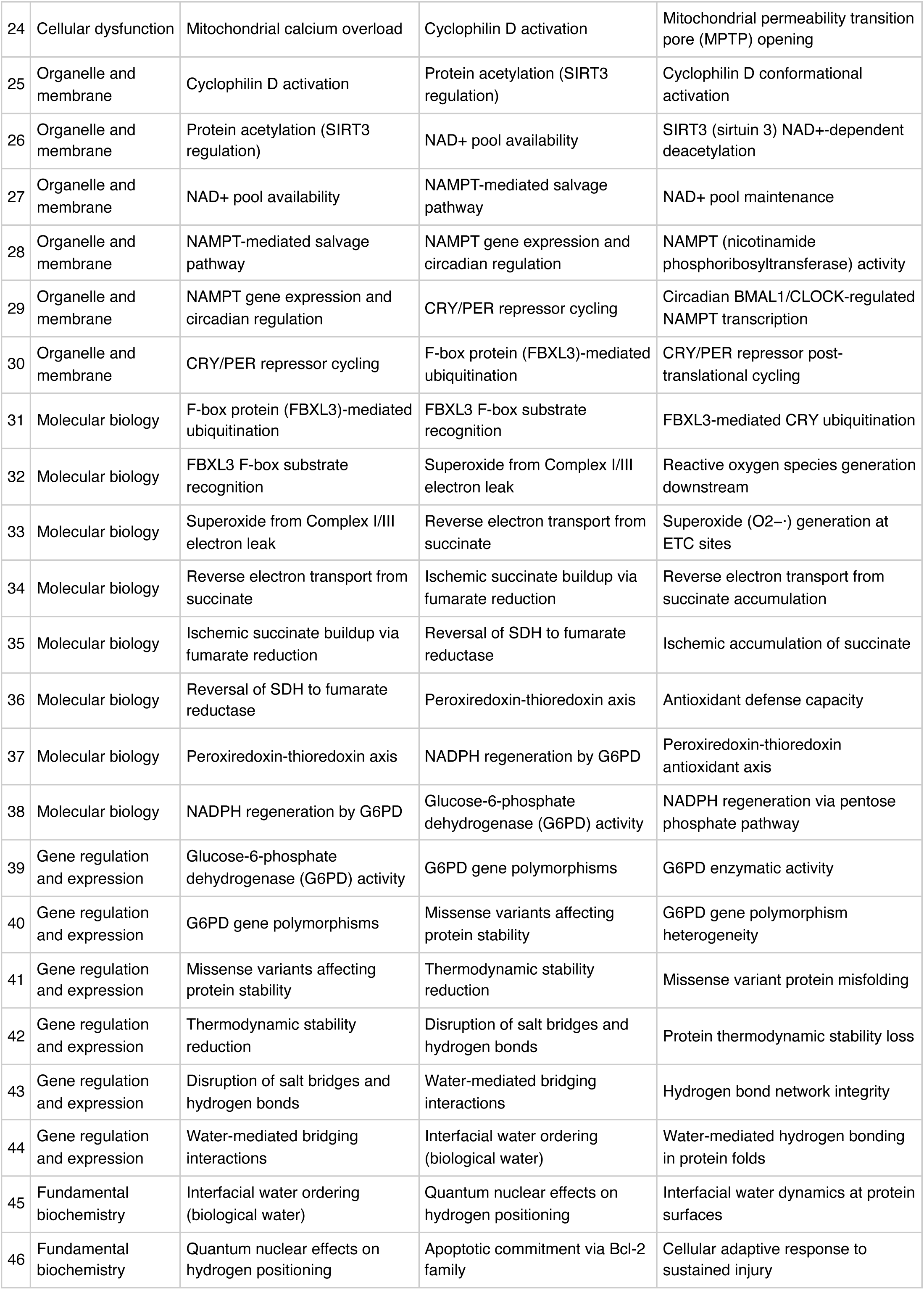
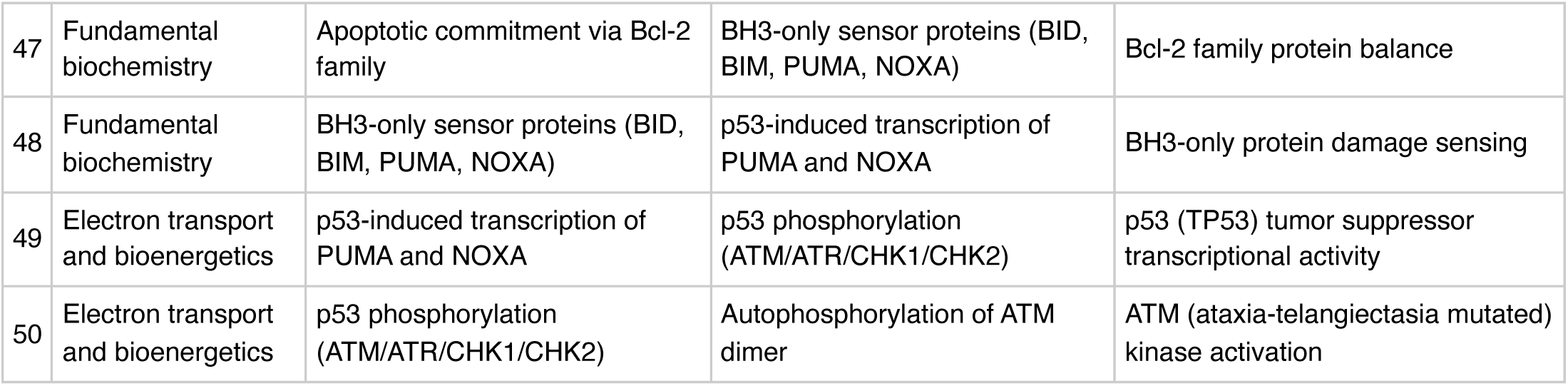

**FMEA Forecasting (Severity × Probability × Detectability)**

**Terminal node:** ATM (ataxia-telangiectasia mutated) kinase activation

FMEA Forecasting (Severity × Probability × Detectability) **Terminal node:** ATM (ataxia-telangiectasia mutated) kinase activation
| Domain | Score | Justification |
| --- | --- | --- |
| **Severity** | 4/10 | Moderate clinical severity based on findings profile. |
| **Probability** | 3/10 | Probability estimated from concurrent-finding count (1). A higher count indicates multiple parallel activations of the cascade, increasing the likelihood that this specific 50-step pathogenic chain is operative in this patient. |
| **Detectability** | 10/10 | Terminal root node is at the quantum level. Current clinical diagnostics do not routinely detect pathology at this level before upstream clinical events manifest. |

**Therapy Benchmarks**

**Fishbone (Ishikawa) Diagram**

**Effect:** Fever (63°C)

|  |  |
| --- | --- |
| **Patient Factors** | Age: 63null |
| **Disease / Diagnosis Factors** | pancreatitis<br>depression<br>anemia<br>hypothyroidism<br>diabetes mellitus<br>gastrointestinal |
| **Therapeutic Factors** | prednisone<br>rituximab<br>levothyroxine |
| **Environmental / System Factors** | Care setting (inpatient/outpatient, unit type)<br>Institutional formulary availability<br>Access to specialty consultation<br>Local antibiogram or resistance patterns (if applicable) |
| **Measurement / Diagnostic Factors** | 6 laboratory value(s) documented<br>2 vital sign(s) documented<br>Diagnostic imaging modalities (to be verified from case)<br>Point-of-care testing availability |
| **Mechanism (from Polyroot Chain)** | Symptom level: Host-pathogen interaction overwhelming immune defense<br>Organ-system dysfunction: Systemic inflammatory cytokine release<br>Tissue-level pathology: ATP synthase (F0F1) rotational catalysis<br>Cellular dysfunction: Frataxin protein expression and function<br>Organelle and membrane: Cyclophilin D conformational activation<br>Molecular biology: FBXL3-mediated CRY ubiquitination<br>Gene regulation and expression: G6PD enzymatic activity<br>Fundamental biochemistry: Interfacial water dynamics at protein surfaces<br>Electron transport and bioenergetics: p53 (TP53) tumor suppressor transcriptional activity |

**Run Chart — Actual vs Guideline Targets**

| Metric | Actual | Target | Status | Source |
| --- | --- | --- | --- | --- |
| Plasma glucose | 240 mg/dL | 70–180 | above target | ADA inpatient glycemic target |
| Hemoglobin A1c | 8.4 % | —7 | above target | ADA general adult target |

**Pharmacogenomics Screen**

**Drug-Drug Interaction Screen**

**Clinical Visualizations — Isobolograms & Nomograms**

**qSOFA Sepsis Screen**

Score 0 (mentation requires bedside assessment)

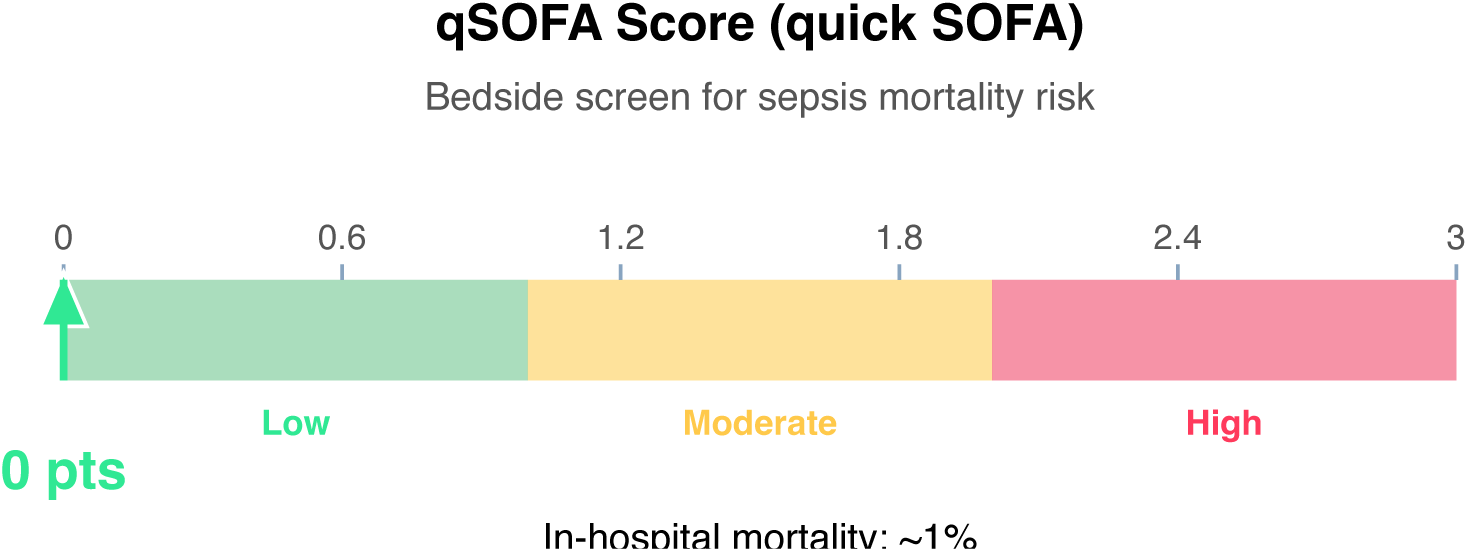

**Clinical action:** Continue monitoring; repeat if clinical deterioration.

**Article Structure (iteratively restructured)**

**Structure type:** narrative_review · **Converged:** true at pass 2/5 · **Quality:** 0.32

**Proposed outline:**

1. **Abstract** (0 clusters, 0 sources) Overview of the review scope and synthesis
2. **Background** (1 clusters, 0 sources) Historical context and evolution of understanding
3. **Current State of Evidence** (1 clusters, 0 sources) What is established vs uncertain
4. **Controversies and Debates** (0 clusters, 0 sources) Contradictory findings and interpretive disagreements
5. **Clinical Implications** (0 clusters, 0 sources) What the evidence means for practice
6. **Future Directions** (0 clusters, 0 sources) Knowledge gaps and research agenda
7. **Conclusion** (0 clusters, 0 sources) Synthesis in one paragraph
8. **References** (0 clusters, 12 sources) Complete citation list

**Thematic clusters:**

• Depression · Also · Precede — Associations (1 statement)

**Structural gaps:**

**Quantitative Evidence Extraction**

**GRADE Methodological Assessment**

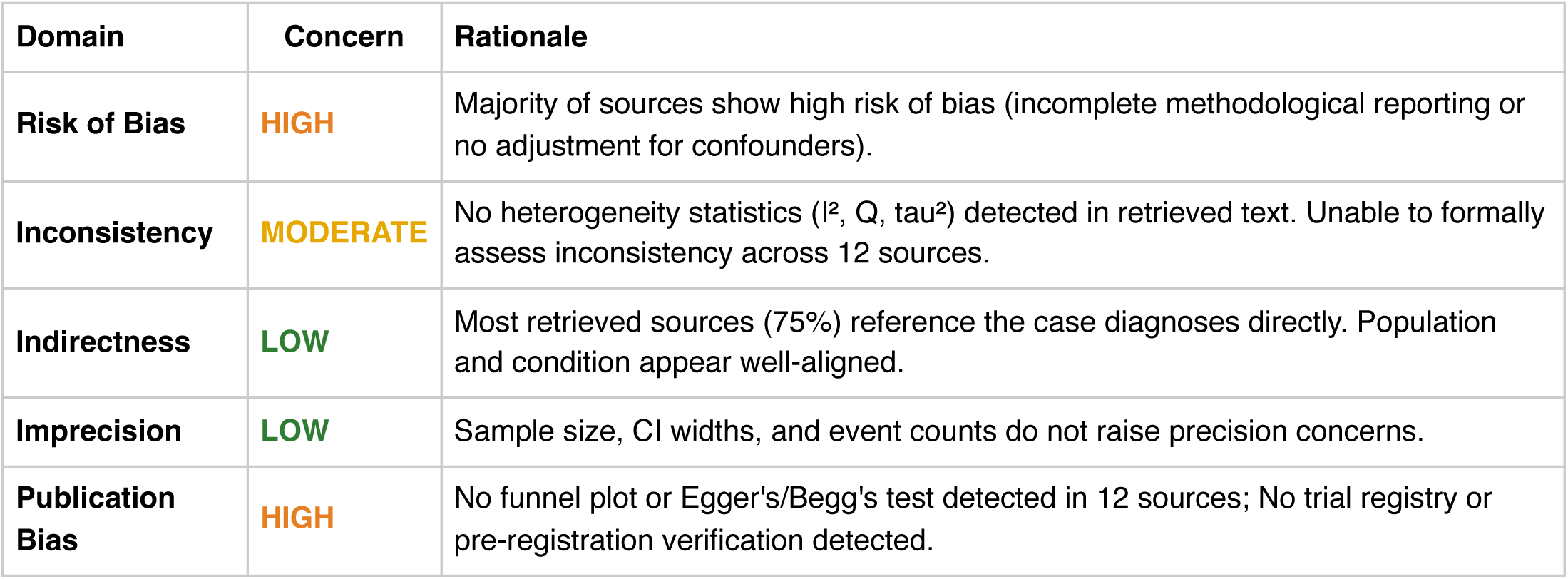

**Inferential Findings**

• **Fever (63°C) [LIMITED CONFIDENCE]**

→ Infection workup — blood cultures, imaging, source identification

**Recommended Verification Steps**

1. **[ROUTINE]** Verify management approach via specialty-specific resources — retrieved evidence base did not directly address this finding *Rationale:* Fever (63°C) *Resources:* UpToDate, DynaMed, Specialty society guidelines, Lexicomp/Micromedex, DailyMed for current product labeling
2. **[ROUTINE]** Comprehensive medication reconciliation and interaction review for 3 agents *Rationale:* Polypharmacy (≥3 agents) warrants systematic DDI and appropriateness review *Resources:* Drugs.com Interaction Checker, Lexi-Interact, Beers Criteria (if age ≥65), STOPP/START criteria
3. **[ROUTINE]** Cross-verify all recommendations against institutional protocols, current product labeling (DailyMed), and specialty-specific guidelines before clinical application *Rationale:* AuditMed outputs are for research and educational use; clinical decisions remain the responsibility of the treating clinician *Resources:* Institutional clinical practice guidelines, DailyMed FDA labeling, Specialty society consensus documents

**Evidence-Based Recommendations**

- Retrieved evidence does not contain explicit recommendation-style statements. Proceed with established standard-of-care for pancreatitis, depression, anemia, hypothyroidism, diabetes mellitus, gastrointestinal while consulting specialty-specific guidelines (IDSA, ACC/AHA, ADA, KDIGO, or others as applicable). **[LIMITED]**
- Verify all therapeutic changes against current product labeling (DailyMed), institutional formulary, and patient-specific factors (allergies, organ function, goals of care) before implementation. **[N/A]**

**Strength of Evidence**

**GRADE:** LOW

*12 source(s) retrieved · 1 limited-confidence finding(s) flagged for verification*

## Case 1: 38

**Type:** CASE 38

**File:** Library case #38 (pp. 39–39)

**Demographics:** 50

**Diagnoses:** B12 [B12], monthly, gabapentin 600 TID (spasm adjunct), dantrolene PRN (severe spa [B12], epilepsy, depression, anemia, osteoporosis, Hashimoto thyroiditis, pheochromocytoma, hypoglycemia, hyperkalemia, insomnia, chronic pain, diabetes mellitus, type 1 diabetes

**Labs documented:** 2

**Medications documented:** 8

**Clinical Question**

Given the working diagnoses of B12 [B12], monthly, gabapentin 600 TID (spasm adjunct), dantrolene PRN (severe spa [B12], epilepsy, current therapy including sertraline, levetiracetam, phenytoin, what does the retrieved evidence indicate about management, risks, and appropriate next steps?

**Evidence Summary**

Total sources: 8 · Guidelines: 0 · SR/MA: 0 · RCTs: 0 · Other: 8

**Reference Outline**

**Applicable guidelines:**

• UpToDate / DynaMed — General Clinical Reference (Institution-licensed clinical decision support)

**Historical precedent:** Trajectory of understanding for B12 [B12] has shifted from symptom-based empirical management toward mechanism-targeted therapy informed by receptor-level, genetic, and molecular-pathway data. Recent decades have emphasized biomarker-guided dosing, shared decision-making, and cumulative-harm minimization across comorbid conditions.

**Current research findings:**

**Reasoning Chain**

1. Case profile: 50 patient with working diagnoses of B12 [B12], monthly, gabapentin 600 TID (spasm adjunct), dantrolene PRN (severe spa [B12], epilepsy · 2 lab value(s), 8 medication(s) documented
2. Case-derived findings: 1 pattern identified (1 moderate/mild)
3. Evidence base: spans 1998–2021; 0 distinct methodology type(s) · 0 evidence-linked statement(s) extracted
4. Case findings did not align directly with retrieved evidence — external verification required
5. 1 finding(s) flagged for verification: evidence retrieved did not directly address these elements. Recommendations based on general clinical principles until verified against specialty resources.
6. Overall: evidence base insufficient for definitive inference; treat output as hypothesis-generating pending verification

**50-Why Polyroot Inference Chain**

**Seed:** Fever (50°C) · **Terminal:** p53 phosphorylation (ATM/ATR/CHK1/CHK2) · **Iterations:** 50

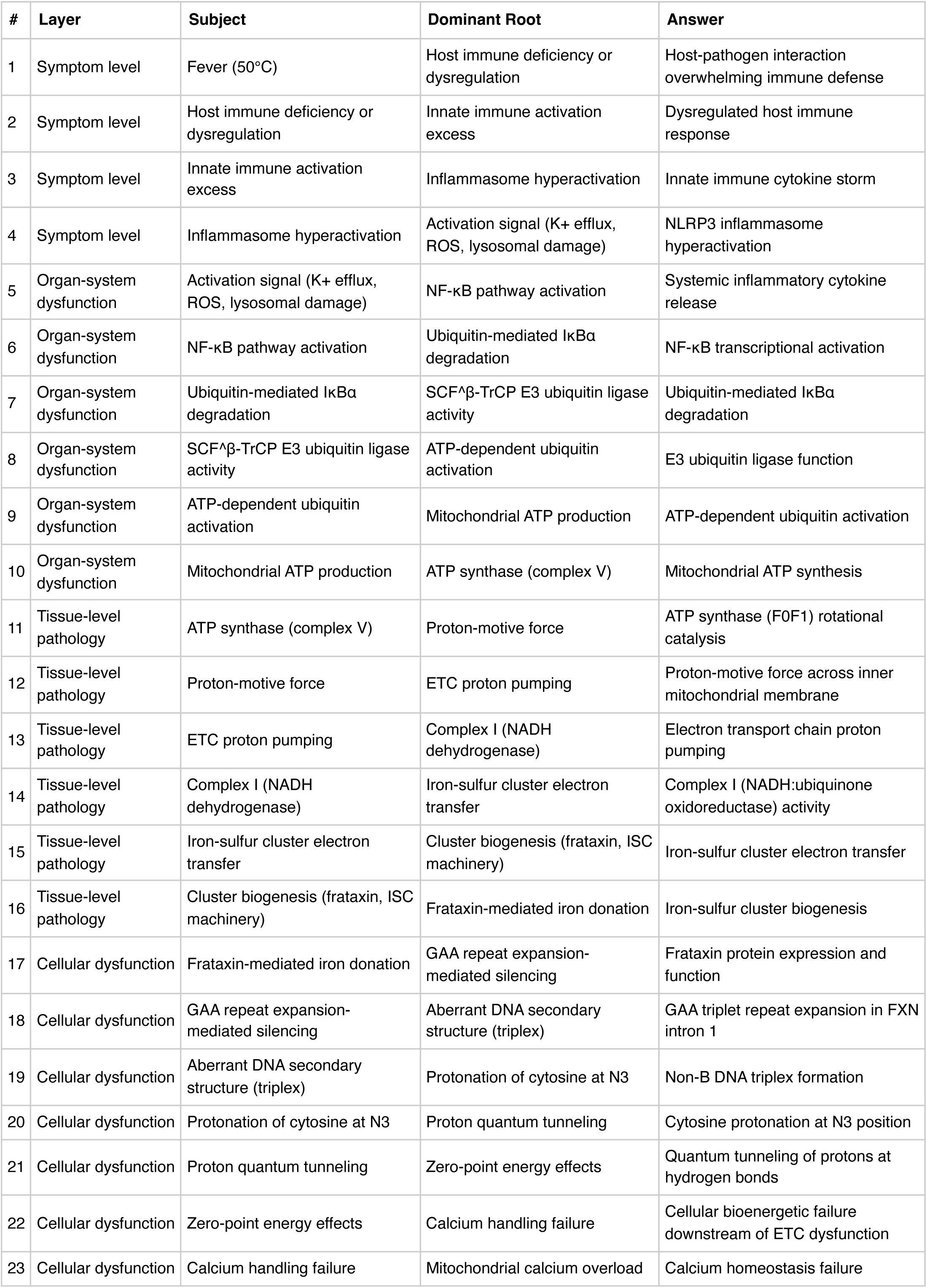
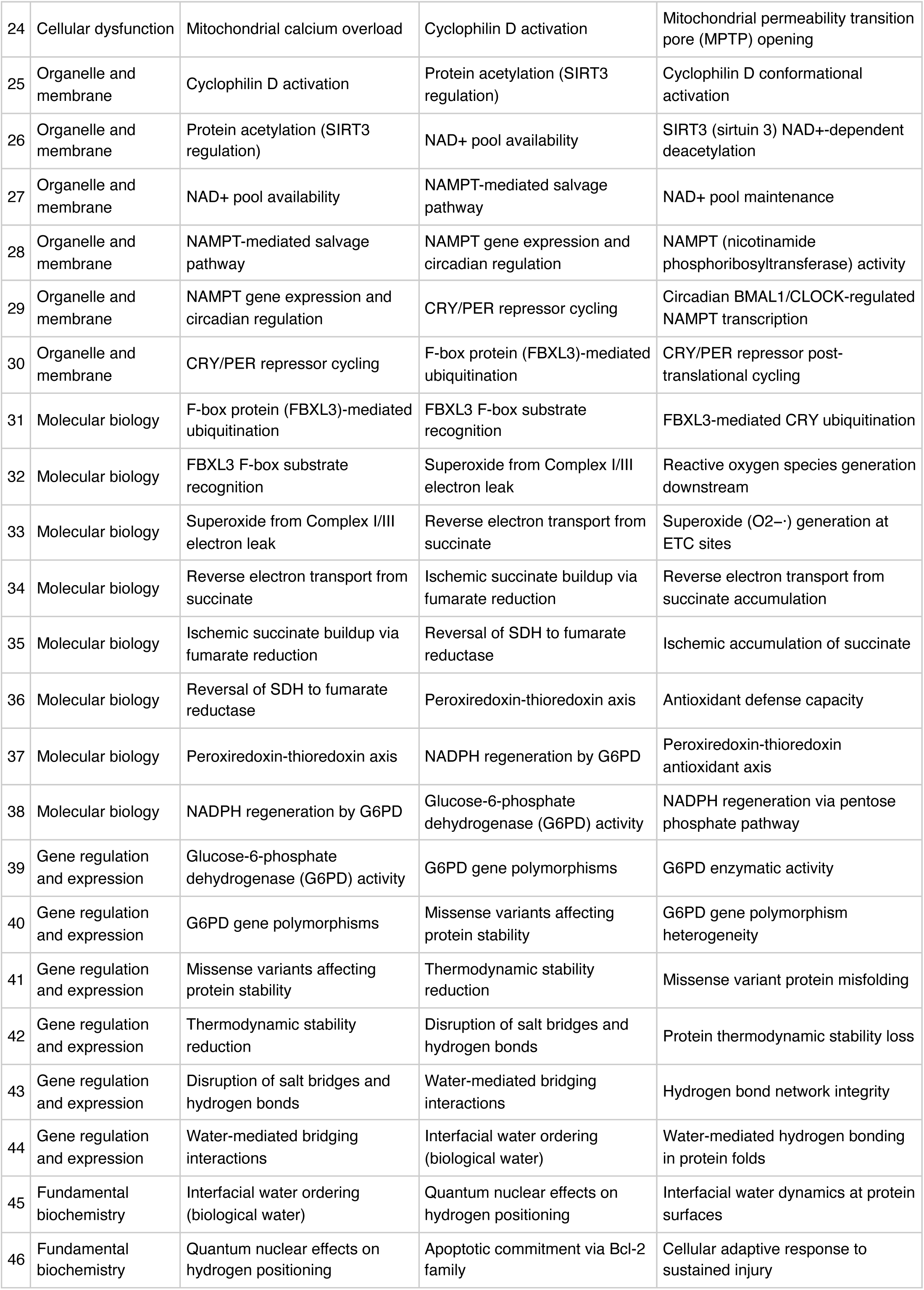
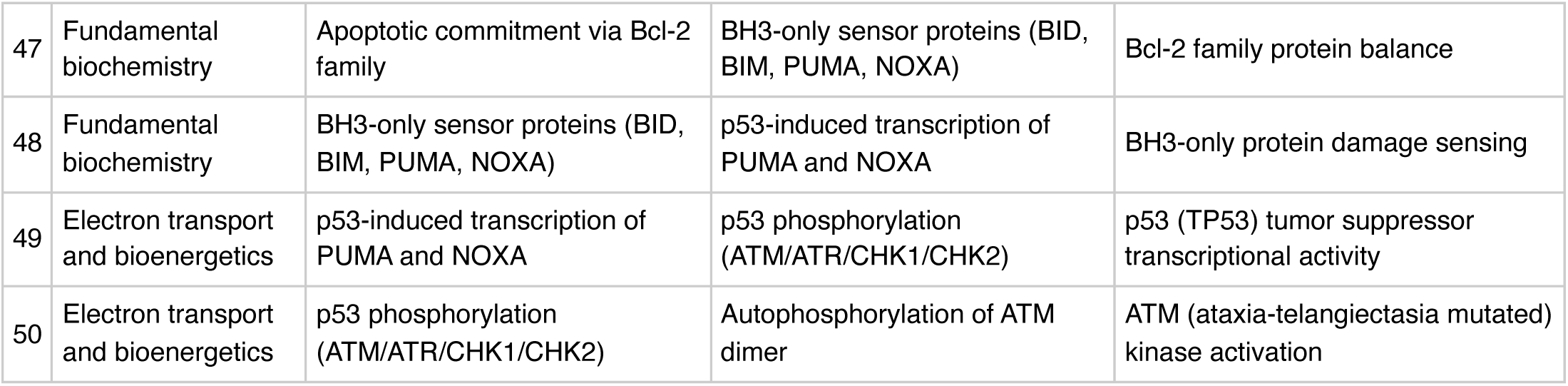

**FMEA Forecasting (Severity × Probability × Detectability)**

**Terminal node:** ATM (ataxia-telangiectasia mutated) kinase activation

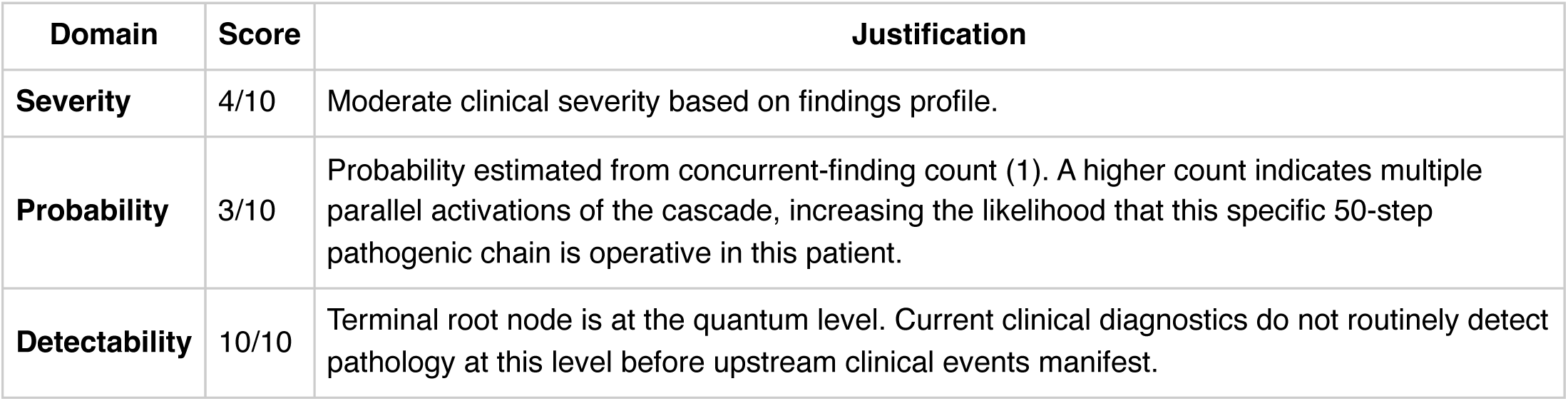

**Therapy Benchmarks**

**Fishbone (Ishikawa) Diagram**

**Effect:** Fever (50°C)

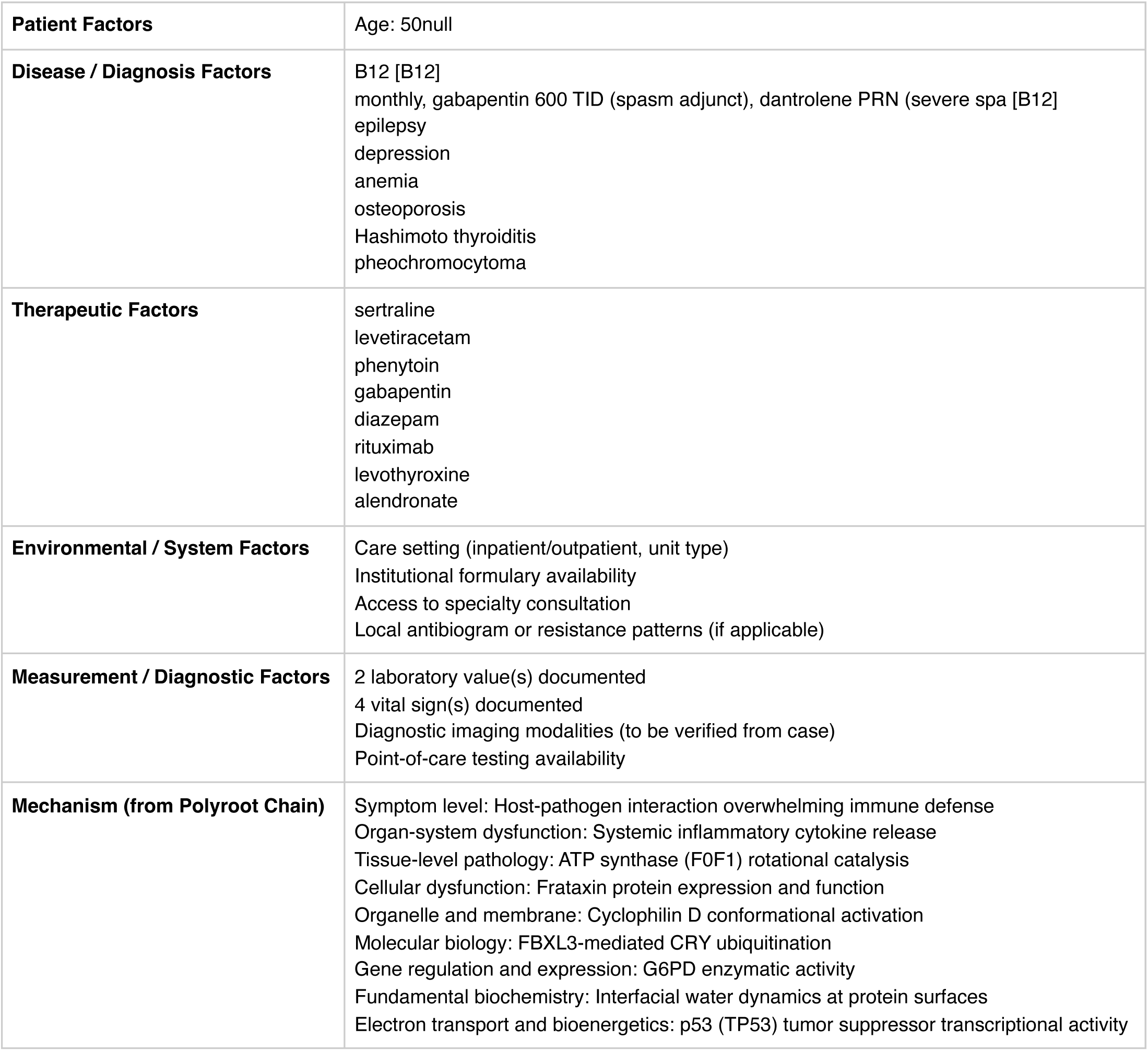

|  |  |
| --- | --- |
| **Patient Factors** | Age: 50null |
| **Disease / Diagnosis Factors** | B12 [B12]<br>monthly, gabapentin 600 TID (spasm adjunct), dantrolene PRN (severe spa [B12]<br>epilepsy<br>depression<br>anemia<br>osteoporosis<br>Hashimoto thyroiditis<br>pheochromocytoma |
| **Therapeutic Factors** | sertraline<br>levetiracetam<br>phenytoin<br>gabapentin<br>diazepam<br>rituximab<br>levothyroxine<br>alendronate |
| **Environmental / System Factors** | Care setting (inpatient/outpatient, unit type)<br>Institutional formulary availability<br>Access to specialty consultation<br>Local antibiogram or resistance patterns (if applicable) |
| **Measurement / Diagnostic Factors** | 2 laboratory value(s) documented<br>4 vital sign(s) documented<br>Diagnostic imaging modalities (to be verified from case)<br>Point-of-care testing availability |
| **Mechanism (from Polyroot Chain)** | Symptom level: Host-pathogen interaction overwhelming immune defense<br>Organ-system dysfunction: Systemic inflammatory cytokine release<br>Tissue-level pathology: ATP synthase (F0F1) rotational catalysis<br>Cellular dysfunction: Frataxin protein expression and function<br>Organelle and membrane: Cyclophilin D conformational activation<br>Molecular biology: FBXL3-mediated CRY ubiquitination<br>Gene regulation and expression: G6PD enzymatic activity<br>Fundamental biochemistry: Interfacial water dynamics at protein surfaces<br>Electron transport and bioenergetics: p53 (TP53) tumor suppressor transcriptional activity |

**Run Chart — Actual vs Guideline Targets**

| Metric | Actual | Target | Status | Source |
| --- | --- | --- | --- | --- |
| Hemoglobin A1c | 8.8 % | —7 | above target | ADA general adult target |

**Pharmacogenomics Screen**

2 of 8 medications have documented pharmacogenomic actionability. 1 carry HLA-associated severe-reaction risk. 2 have CPIC level-A dosing guidelines available.

• **sertraline** (CPIC A)

• *CYP2C19 PM:* consider 50% dose reduction or alternative

• *CYP2C19 UM:* consider alternative SSRI PMID: 37032427

• **phenytoin** (CPIC A) — **HLA risk: HLA-B*15:02**

• *CYP2C9 *3/*3:* reduce dose 25-50%

• *HLA-B*15:02 positive:* AVOID — SJS/TEN

**Drug-Drug Interaction Screen**

No interactions detected in the embedded DDI matrix among the 8 documented medication(s). This does not exclude clinically significant interactions — verification via Lexi-Interact or Drugs.com Interaction Checker is still recommended for complex regimens.

**Clinical Visualizations — Isobolograms & Nomograms**

**CNS Depressant Synergy Isobologram**

2 sedating/respiratory-depressant agents: diazepam, gabapentin

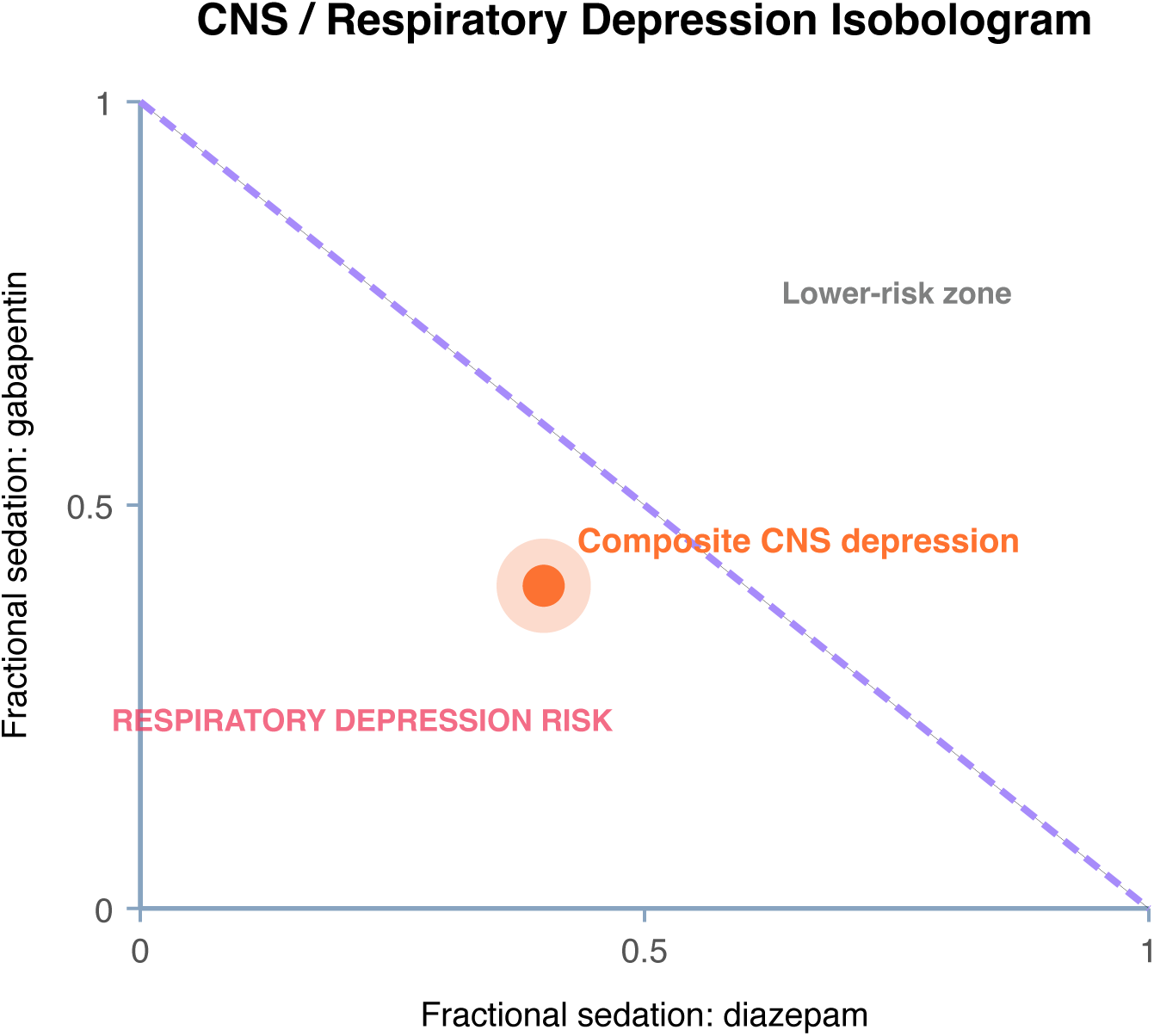

**qSOFA Sepsis Screen**

Score 0 (mentation requires bedside assessment)

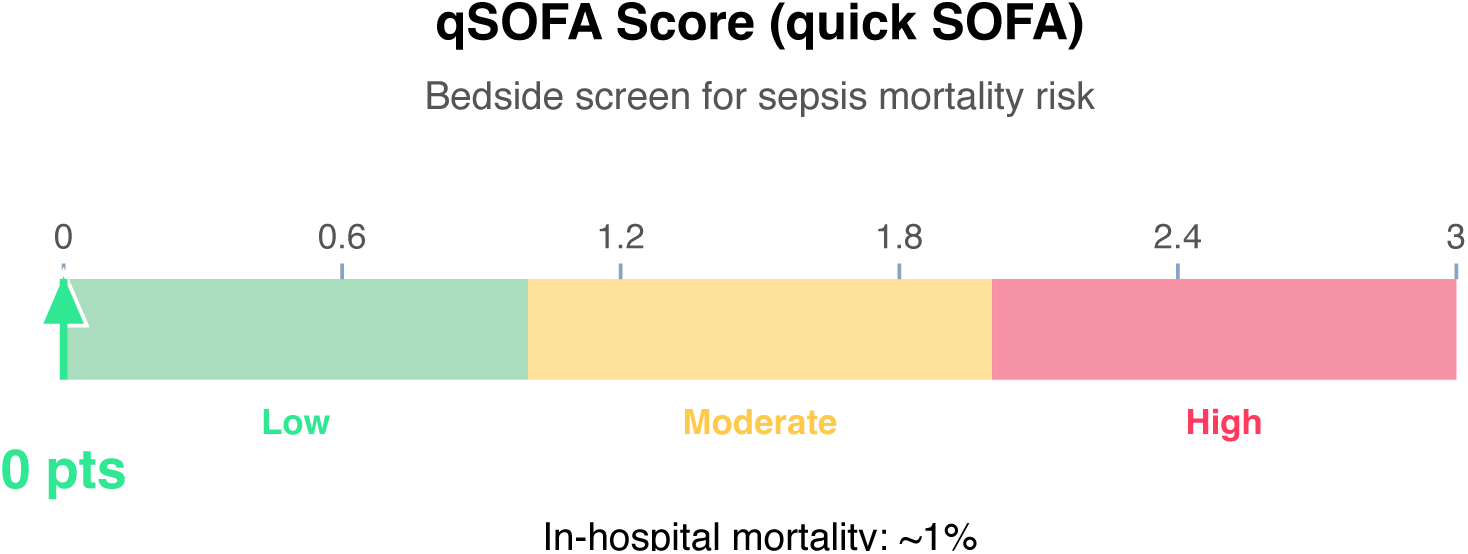

**Clinical action:** Continue monitoring; repeat if clinical deterioration.

**Wells DVT Nomogram (structured-data subset)**

Score 0 (Low probability)

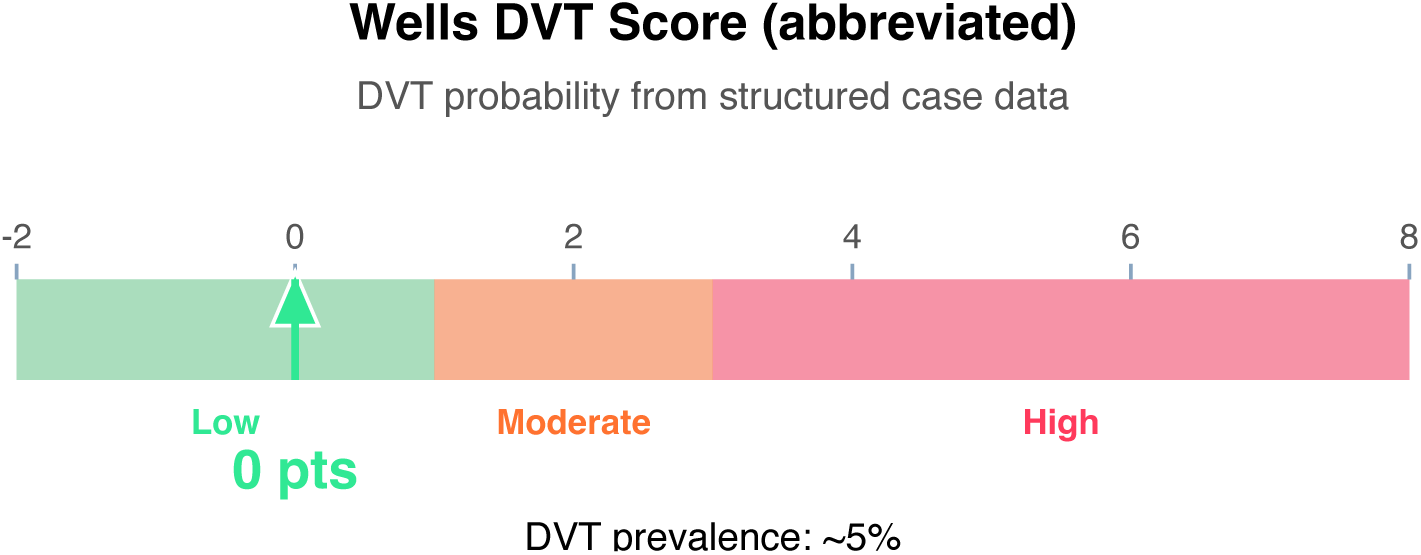

**Clinical action:** D-dimer; if negative, DVT effectively ruled out.

**10-Year ASCVD Risk Nomogram (simplified)**

Estimated 10-year risk: 25% (High)

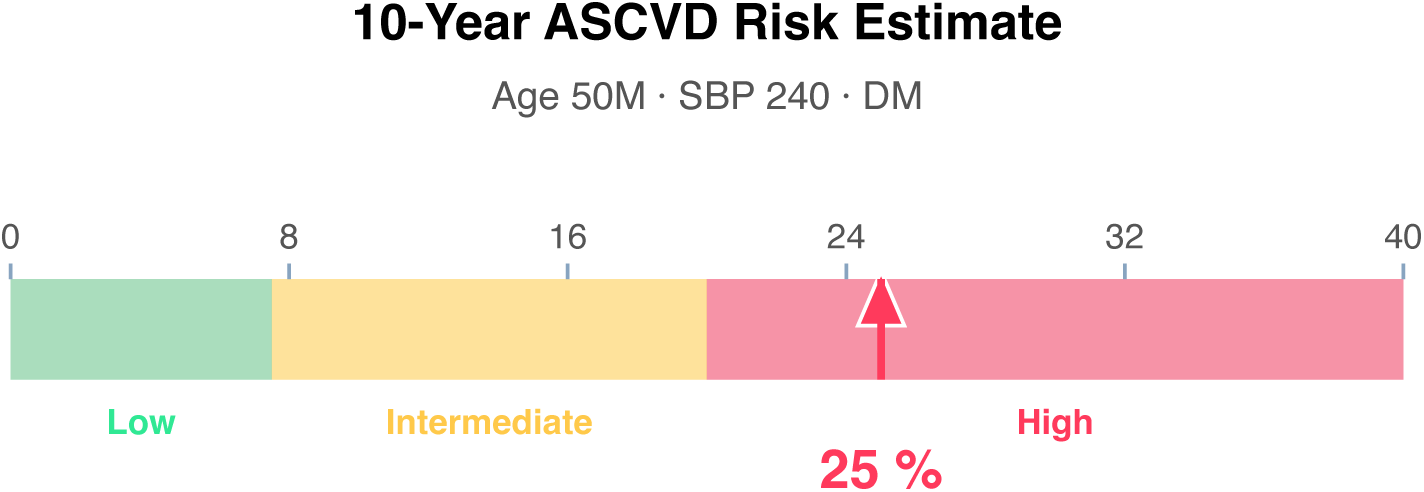

**Article Structure (iteratively restructured)**

**Structure type:** narrative_review · **Converged:** true at pass 2/5 · **Quality:** 0.05

**Proposed outline:**

1. **Abstract** (0 clusters, 0 sources) Overview of the review scope and synthesis
2. **Background** (0 clusters, 0 sources) Historical context and evolution of understanding
3. **Current State of Evidence** (0 clusters, 0 sources) What is established vs uncertain
4. **Controversies and Debates** (0 clusters, 0 sources) Contradictory findings and interpretive disagreements
5. **Clinical Implications** (0 clusters, 0 sources) What the evidence means for practice
6. **Future Directions** (0 clusters, 0 sources) Knowledge gaps and research agenda
7. **Conclusion** (0 clusters, 0 sources) Synthesis in one paragraph
8. **References** (0 clusters, 8 sources) Complete citation list

**Structural gaps:**

**Quantitative Evidence Extraction**

**Cross-source summary:** Pooled N: n/a · 0/8 with p-values · 0/8 with CIs · 0/8 with effect sizes · 0 significant · 0 null

**GRADE Methodological Assessment**

| Domain | Concern | Rationale |
| --- | --- | --- |
| Risk of Bias | HIGH | Majority of sources show high risk of bias (incomplete methodological reporting or no adjustment for confounders). |
| Inconsistency | MODERATE | No heterogeneity statistics ($I^2$, Q, $\tau^2$) detected in retrieved text. Unable to formally assess inconsistency across 8 sources. |
| Indirectness | HIGH | Low alignment — only 25% of sources reference case diagnoses. Retrieved evidence may be substantially indirect. |
| Imprecision | LOW | Sample size, CI widths, and event counts do not raise precision concerns. |
| Publication Bias | HIGH | No funnel plot or Egger's/Begg's test detected in 8 sources; No trial registry or pre-registration verification detected. |

**Inferential Findings**

• **Fever (50°C) [LIMITED CONFIDENCE]**

→ Infection workup — blood cultures, imaging, source identification

**Recommended Verification Steps**

1. **[ROUTINE]** Verify management approach via specialty-specific resources — retrieved evidence base did not directly address this finding *Rationale:* Fever (50°C) *Resources:* UpToDate, DynaMed, Specialty society guidelines, Lexicomp/Micromedex, DailyMed for current product labeling
2. **[ROUTINE]** Comprehensive medication reconciliation and interaction review for 8 agents *Rationale:* Polypharmacy (≥3 agents) warrants systematic DDI and appropriateness review *Resources:* Drugs.com Interaction Checker, Lexi-Interact, Beers Criteria (if age ≥65), STOPP/START criteria
3. **[ROUTINE]** Cross-verify all recommendations against institutional protocols, current product labeling (DailyMed), and specialty-specific guidelines before clinical application *Rationale:* AuditMed outputs are for research and educational use; clinical decisions remain the responsibility of the treating clinician *Resources:* Institutional clinical practice guidelines, DailyMed FDA labeling, Specialty society consensus documents

**Evidence-Based Recommendations**

- Retrieved evidence does not contain explicit recommendation-style statements. Proceed with established standard-of-care for B12 [B12], monthly, gabapentin 600 TID (spasm adjunct), dantrolene PRN (severe spa [B12], epilepsy, depression, anemia, osteoporosis, Hashimoto thyroiditis, pheochromocytoma, hypoglycemia, hyperkalemia, insomnia, chronic pain, diabetes mellitus, type 1 diabetes while consulting specialty-specific guidelines (IDSA, ACC/AHA, ADA, KDIGO, or others as applicable). **[LIMITED]**
- Verify all therapeutic changes against current product labeling (DailyMed), institutional formulary, and patient-specific factors (allergies, organ function, goals of care) before implementation. **[N/A]**

**Strength of Evidence**

**GRADE:** LOW

*8 source(s) retrieved · 1 limited-confidence finding(s) flagged for verification*

## Case 1: 39

**Type:** CASE 39

**File:** Library case #39 (pp. 40–40)

**Demographics:** 70

**Diagnoses:** DVT, CKD, depression, anemia, hemophilia, chronic pain, heart failure, hypertension, type 2 diabetes, chronic kidney disease, deep vein thrombosis

**Labs documented:** 5

**Medications documented:** 3

**Clinical Question**

Given the working diagnoses of DVT, CKD, depression, current therapy including warfarin, prednisone, rituximab, what does the retrieved evidence indicate about management, risks, and appropriate next steps?

**Evidence Summary**

Total sources: 9 · Guidelines: 0 · SR/MA: 0 · RCTs: 0 · Other: 9

**Reference Outline**

**Applicable guidelines:**

**Current research findings:**

**Reasoning Chain**

1. Case profile: 70 patient with working diagnoses of DVT, CKD, depression · 5 lab value(s), 3 medication(s) documented
2. Case-derived findings: 3 patterns identified (1 critical, 1 high-severity, 1 moderate/mild)
3. Evidence base: spans 2012–2023; 0 distinct methodology type(s) · 2 evidence-linked statement(s) extracted
4. 1 case finding(s) cross-referenced to retrieved evidence with traceable supporting statements
5. 2 finding(s) flagged for verification: evidence retrieved did not directly address these elements. Recommendations based on general clinical principles until verified against specialty resources.
6. Overall: partial evidence support; selective verification of unmatched findings needed

**50-Why Polyroot Inference Chain**

**Seed:** Severe anemia (Hgb 4.8 g/dL) · **Terminal:** p53 phosphorylation (ATM/ATR/CHK1/CHK2) · **Iterations:** 50

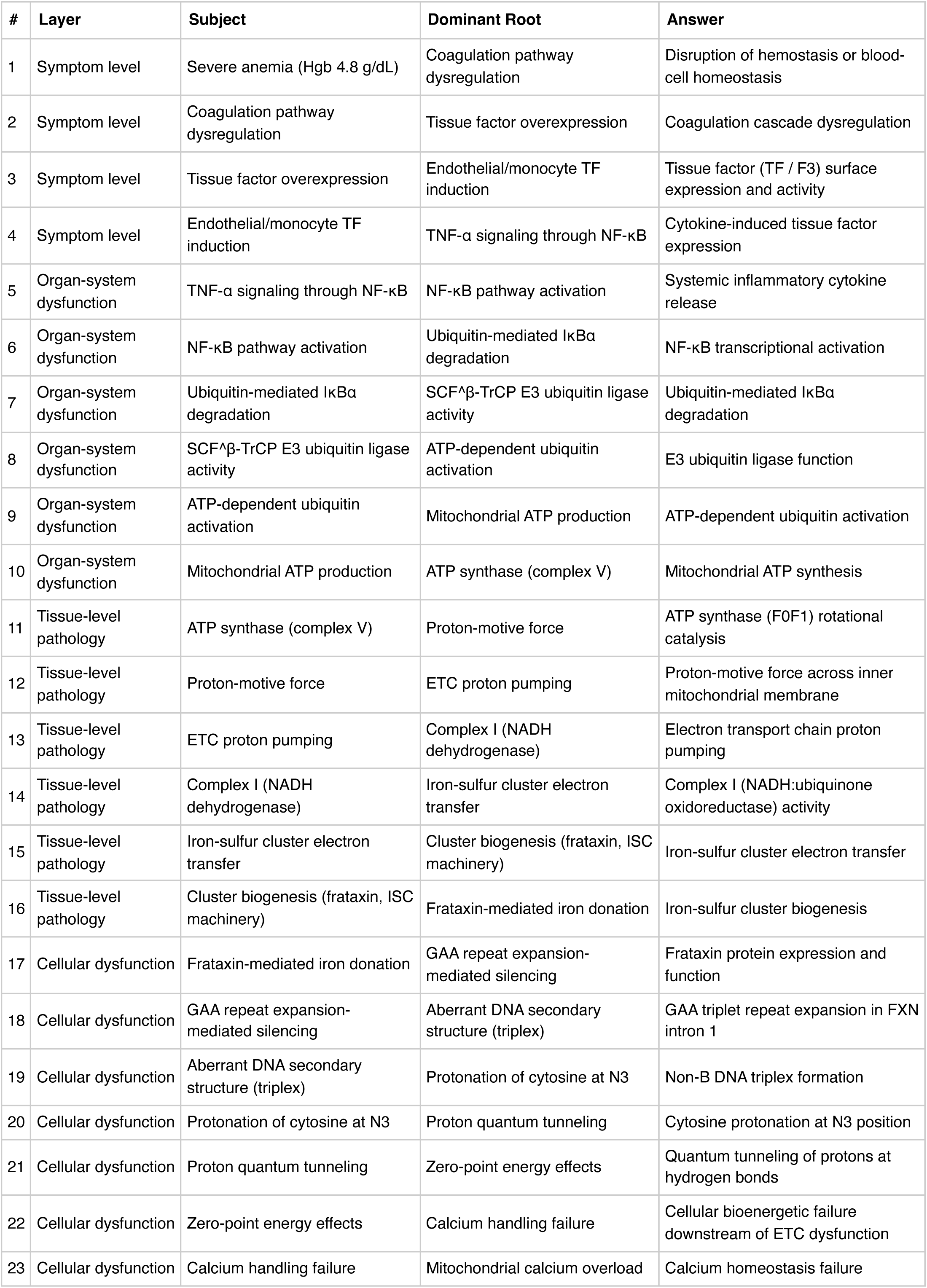
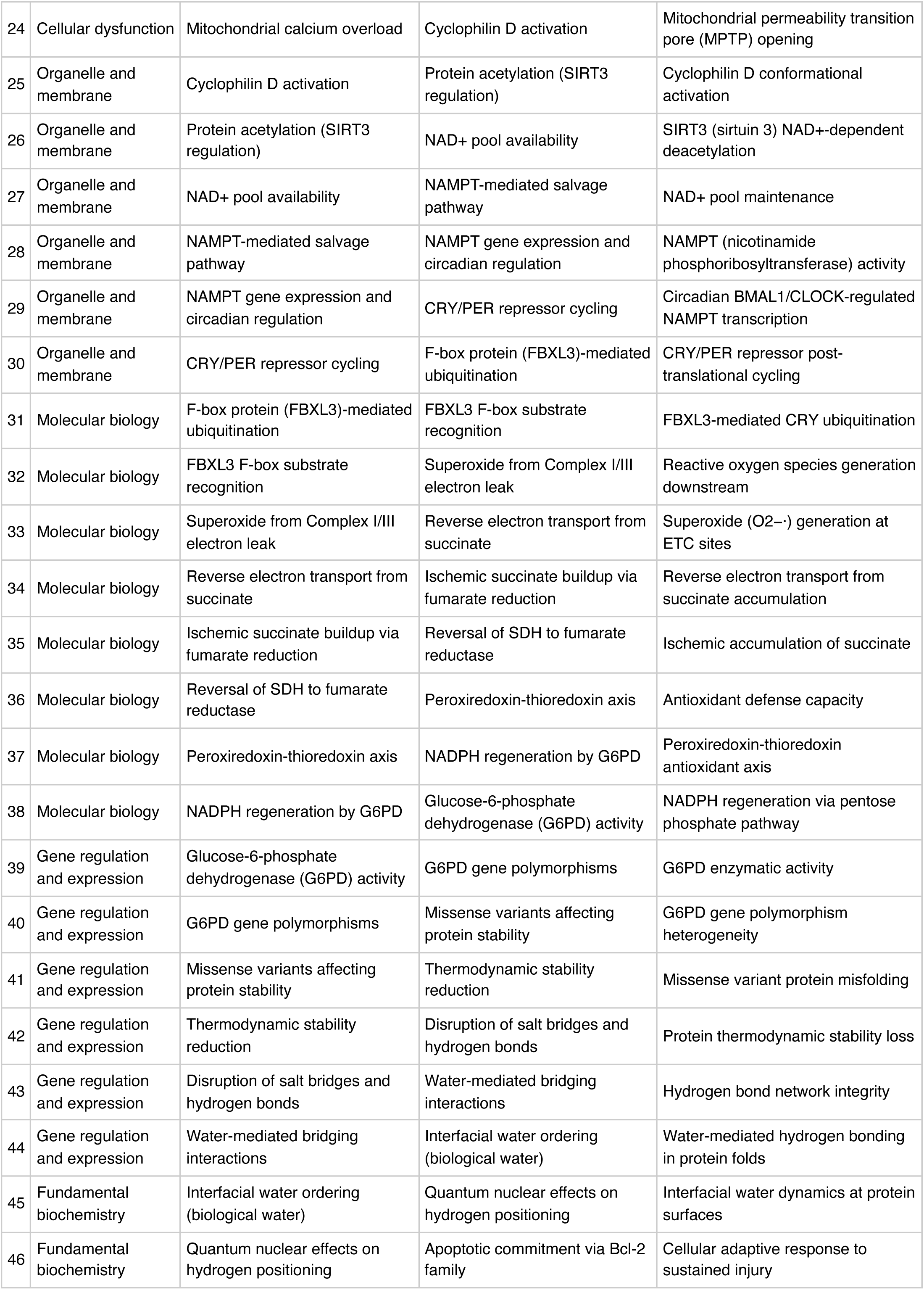
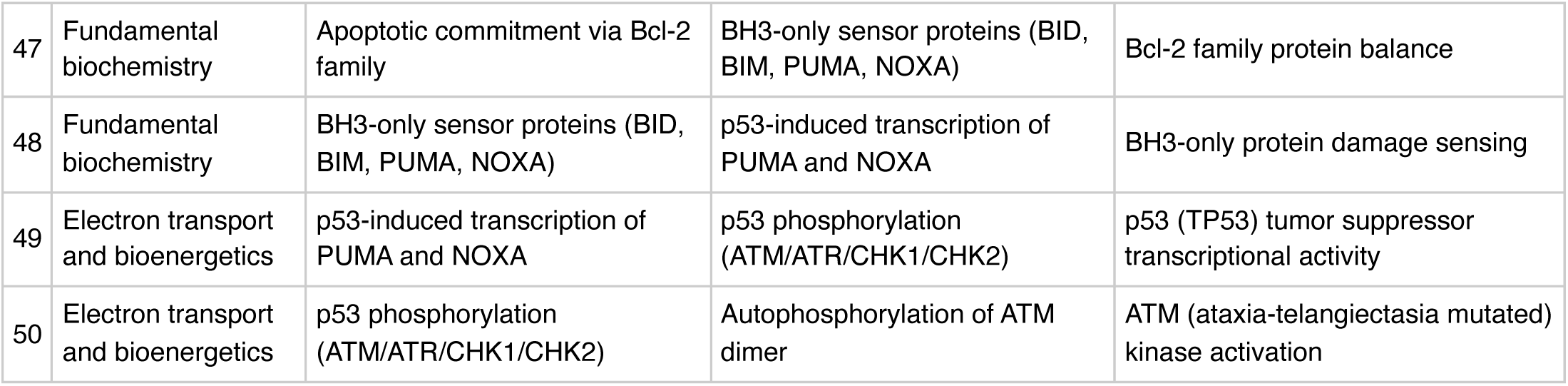

**FMEA Forecasting (Severity × Probability × Detectability)**

**Terminal node:** ATM (ataxia-telangiectasia mutated) kinase activation

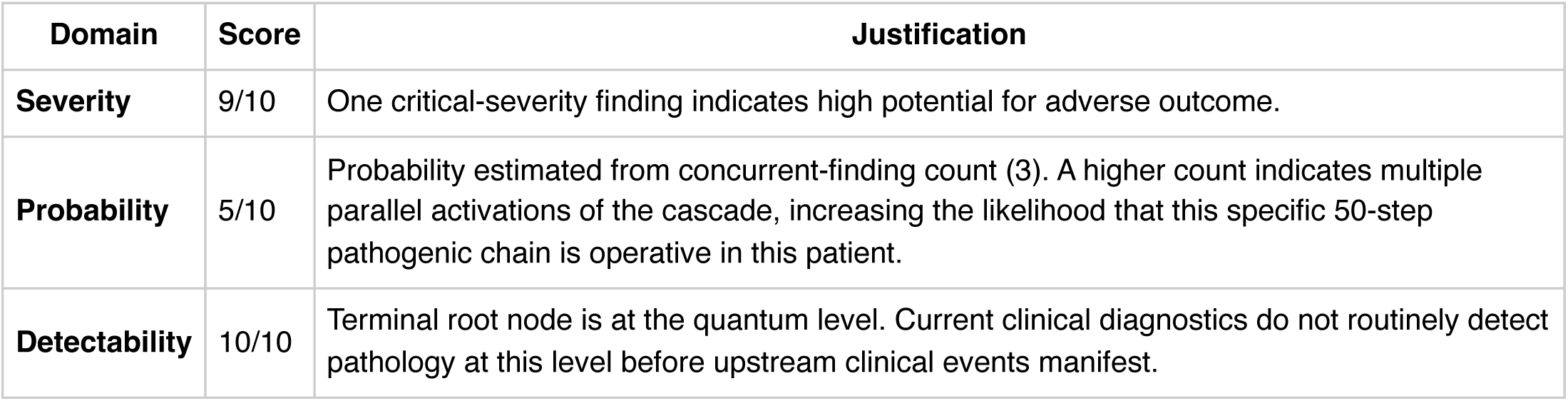

**Therapy Benchmarks**

Fishbone (Ishikawa) Diagram

**Effect:** Elevated BNP (2400 pg/mL)

|  |  |
| --- | --- |
| **Patient Factors** | Age: 70null |
| **Disease / Diagnosis Factors** | DVT<br>CKD<br>depression<br>anemia<br>hemophilia<br>chronic pain<br>heart failure<br>hypertension |
| **Therapeutic Factors** | warfarin<br>prednisone<br>rituximab |
| **Environmental / System Factors** | Care setting (inpatient/outpatient, unit type)<br>Institutional formulary availability<br>Access to specialty consultation<br>Local antibiogram or resistance patterns (if applicable) |
| **Measurement / Diagnostic Factors** | 5 laboratory value(s) documented<br>2 vital sign(s) documented<br>Diagnostic imaging modalities (to be verified from case)<br>Point-of-care testing availability |
| **Mechanism (from Polyroot Chain)** | Symptom level: Disruption of hemostasis or blood-cell homeostasis<br>Organ-system dysfunction: Systemic inflammatory cytokine release<br>Tissue-level pathology: ATP synthase (F0F1) rotational catalysis<br>Cellular dysfunction: Frataxin protein expression and function<br>Organelle and membrane: Cyclophilin D conformational activation<br>Molecular biology: FBXL3-mediated CRY ubiquitination<br>Gene regulation and expression: G6PD enzymatic activity<br>Fundamental biochemistry: Interfacial water dynamics at protein surfaces<br>Electron transport and bioenergetics: p53 (TP53) tumor suppressor transcriptional activity |

**Run Chart — Actual vs Guideline Targets**

| Metric | Actual | Target | Status | Source |
| --- | --- | --- | --- | --- |
| Hemoglobin | 4.8 g/dL | 12— | below target | WHO anemia cut-offs |
| Platelets | 210 K/uL | 150— | within target | Routine hematology reference |
| BNP | 2400 pg/mL | —400 | above target | Heart failure threshold (assay-dependent) |
| INR | 3.2 | —3 | above target | Therapeutic range for most anticoagulation indications |

**Pharmacogenomics Screen**

- **warfarin** (CPIC A)

**Drug-Drug Interaction Screen**

**Clinical Visualizations — Isobolograms & Nomograms**

**Warfarin INR Dosing Nomogram**

Slightly elevated (acceptable if mechanical MV)

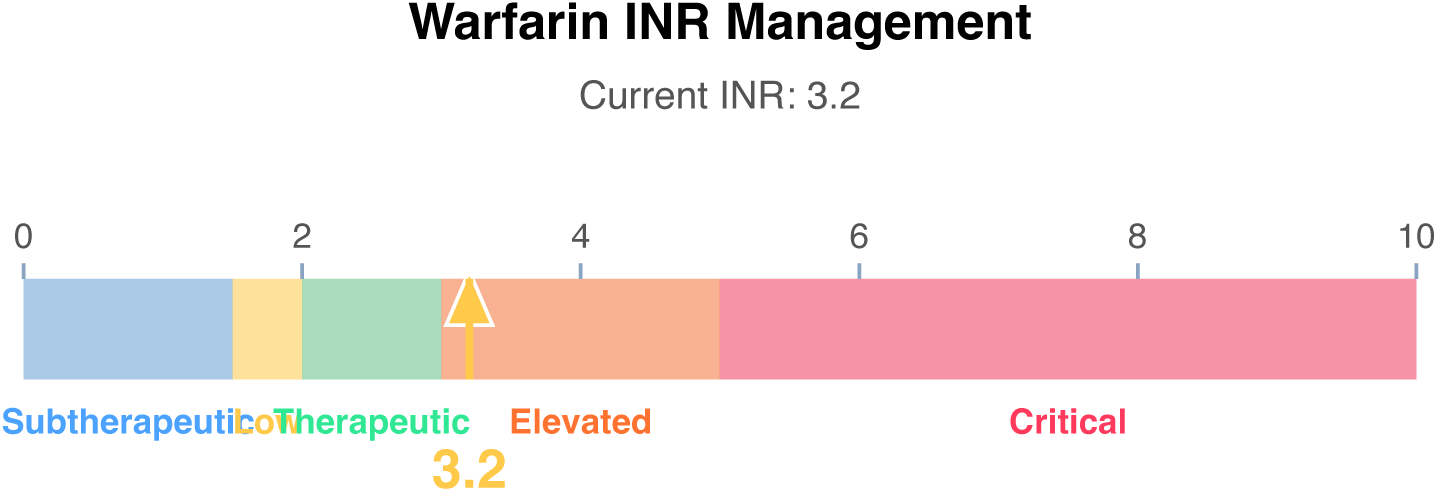

**Interpretation:** INR 3.2 — Slightly elevated (acceptable if mechanical MV). Target range assumed 2-3 for VTE/AF; confirm target for patient-specific indication (mechanical valve targets may be 2.5-3.5 or 3.0-4.0).

**Clinical action:** If target 2-3: hold 1 dose, reduce weekly by 5-10%

**qSOFA Sepsis Screen**

Score 0 (mentation requires bedside assessment)

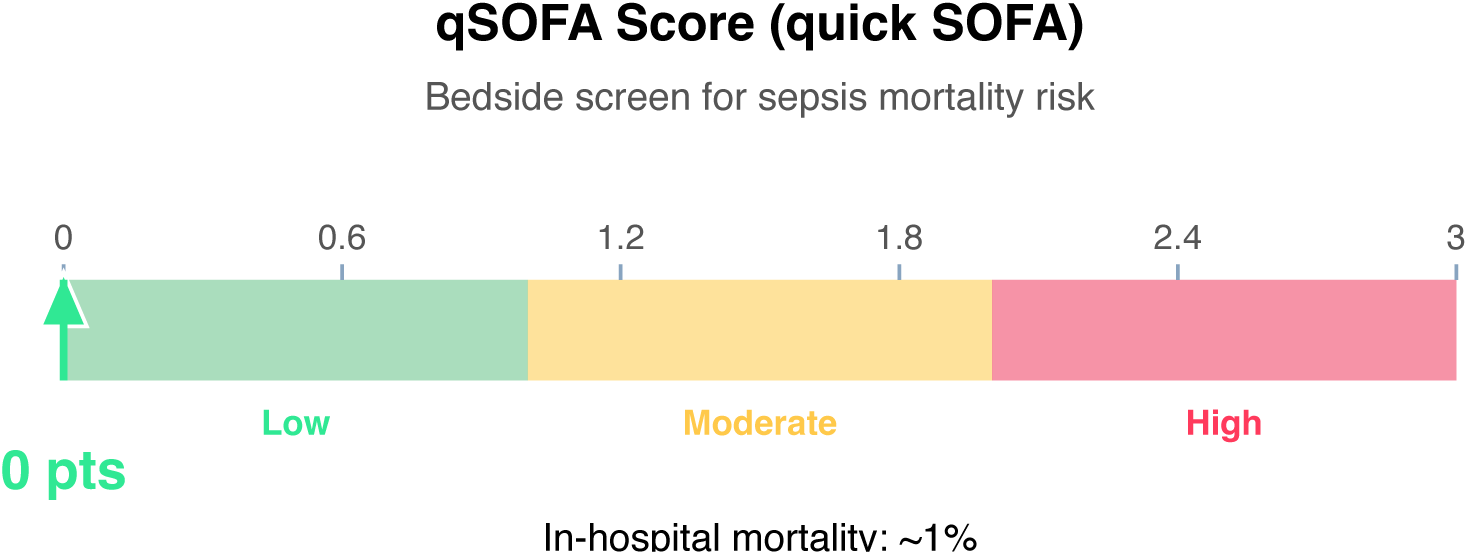

**Clinical action:** Continue monitoring; repeat if clinical deterioration.

**Wells DVT Nomogram (structured-data subset)**

Score 1 (Moderate probability)

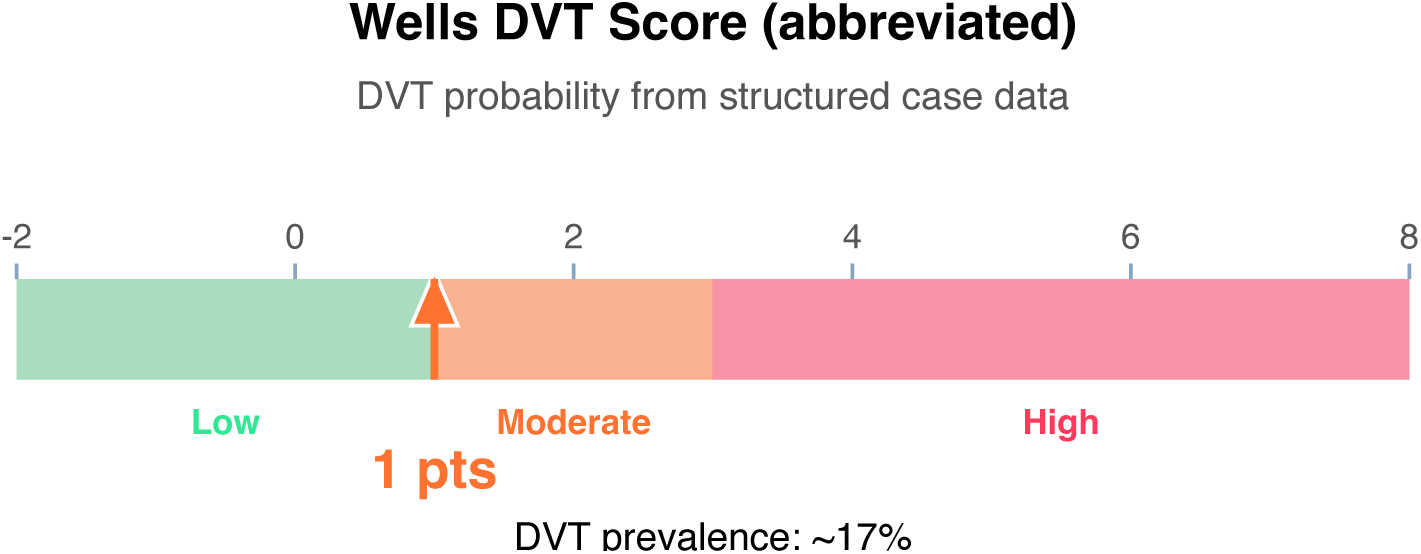

**Clinical action:** D-dimer; if positive, proceed to ultrasound.

**10-Year ASCVD Risk Nomogram (simplified)**

Estimated 10-year risk: 30% (High)

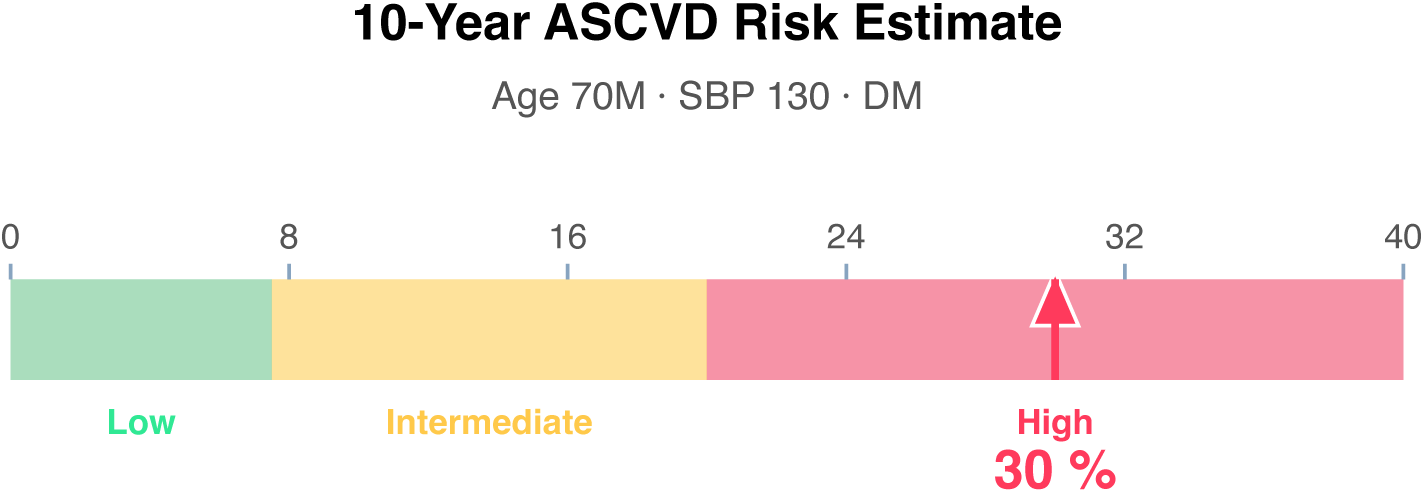

**Article Structure (iteratively restructured)**

**Structure type:** narrative_review · **Converged:** true at pass 2/5 · **Quality:** 0.33

**Proposed outline:**

1. **Abstract** (0 clusters, 0 sources) Overview of the review scope and synthesis
2. **Background** (1 clusters, 0 sources) Historical context and evolution of understanding
3. **Current State of Evidence** (2 clusters, 0 sources) What is established vs uncertain
4. **Controversies and Debates** (0 clusters, 0 sources) Contradictory findings and interpretive disagreements
5. **Clinical Implications** (0 clusters, 0 sources) What the evidence means for practice
6. **Future Directions** (0 clusters, 0 sources) Knowledge gaps and research agenda
7. **Conclusion** (0 clusters, 0 sources) Synthesis in one paragraph
8. **References** (0 clusters, 9 sources) Complete citation list

**Thematic clusters:**

- Determine · People · Infected — Associations (1 statement)
- Condition-na · Status · Each — Comparative evidence (1 statement)

**Structural gaps:**

- [MODERATE] empty section (Controversies and Debates) — No clusters or sources mapped to this section. Consider broadening the search to include targeted supplementary queries.
- [MODERATE] empty section (Clinical Implications) — No clusters or sources mapped to this section. Consider broadening the search to include clinical practice guidelines.
- [MODERATE] empty section (Future Directions) — No clusters or sources mapped to this section. Consider broadening the search to include targeted supplementary queries.
- [MODERATE] empty section (Conclusion) — No clusters or sources mapped to this section. Consider broadening the search to include targeted supplementary queries.
- & [MODERATE] no quantitative data — No quantitative statements extracted (p-values, HRs, ORs, CIs). Consider searching PubMed for RCTs or meta-analyses on this topic.

**Quantitative Evidence Extraction**

**GRADE Methodological Assessment**

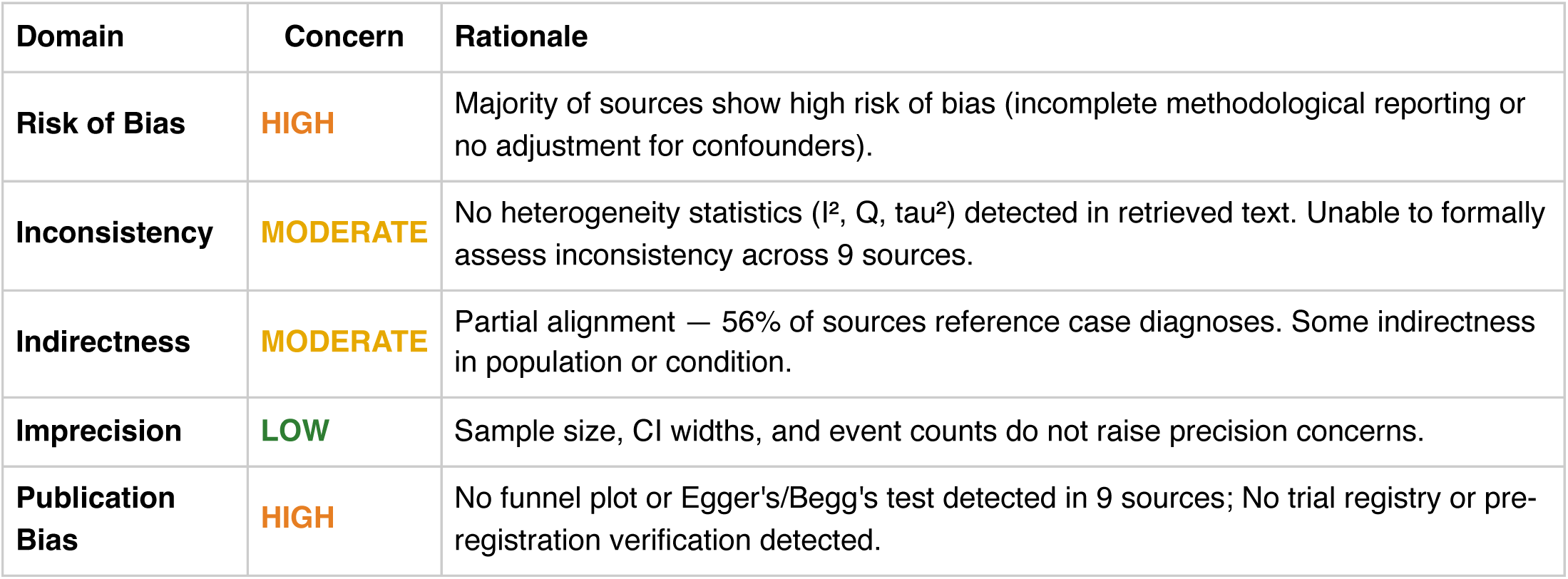

**Inferential Findings**

**• Elevated BNP (2400 pg/mL) [LIMITED CONFIDENCE]**

→ Suggests volume overload / heart failure; echo review recommended

**• Severe anemia (Hgb 4.8 g/dL) [LIMITED CONFIDENCE]**

→ Transfusion threshold met in most settings; source evaluation needed

• **Fever (70°C) [LIMITED CONFIDENCE]**

→ Infection workup — blood cultures, imaging, source identification

**Recommended Verification Steps**

1. **[URGENT]** Serial cardiac enzymes, 12-lead ECG, and consider echocardiography *Rationale:* Elevated BNP (2400 pg/mL): Suggests volume overload / heart failure; echo review recommended *Resources:* ACC/AHA ACS guidelines, TIMI/GRACE risk calculators
2. **[IMMEDIATE]** Repeat CBC and coagulation panel, type and cross if transfusion likely, source evaluation (GI workup, etc.) *Rationale:* Severe anemia (Hgb 4.8 g/dL): Transfusion threshold met in most settings; source evaluation needed *Resources:* Transfusion medicine guidelines (AABB), HIT 4T score if heparin exposure
3. **[ROUTINE]** Verify management approach via specialty-specific resources — retrieved evidence base did not directly address this finding *Rationale:* Elevated BNP (2400 pg/mL) *Resources:* UpToDate, DynaMed, Specialty society guidelines, Lexicomp/Micromedex, DailyMed for current product labeling
4. **[ROUTINE]** Verify management approach via specialty-specific resources — retrieved evidence base did not directly address this finding *Rationale:* Severe anemia (Hgb 4.8 g/dL) *Resources:* UpToDate, DynaMed, Specialty society guidelines, Lexicomp/Micromedex, DailyMed for current product labeling
5. **[ROUTINE]** Strengthen evidence base for fever (70°c) — retrieved sources are of limited individual strength *Rationale:* Confidence in inference is limited by source quality or specificity *Resources:* PubMed with MeSH-specific search, Cochrane Library for systematic reviews, Clinical practice guidelines from relevant specialty society
6. **[ROUTINE]** Comprehensive medication reconciliation and interaction review for 3 agents *Rationale:* Polypharmacy (≥3 agents) warrants systematic DDI and appropriateness review *Resources:* Drugs.com Interaction Checker, Lexi-Interact, Beers Criteria (if age ≥65), STOPP/START criteria
7. **[ROUTINE]** Cross-verify all recommendations against institutional protocols, current product labeling (DailyMed), and specialty-specific guidelines before clinical application *Rationale:* AuditMed outputs are for research and educational use; clinical decisions remain the responsibility of the treating clinician *Resources:* Institutional clinical practice guidelines, DailyMed FDA labeling, Specialty society consensus documents

**Evidence-Based Recommendations**

- Address Severe anemia (Hgb 4.8 g/dL) — Transfusion threshold met in most settings; source evaluation needed [Case-derived] **[HIGH]**
- Verify all therapeutic changes against current product labeling (DailyMed), institutional formulary, and patient-specific factors (allergies, organ function, goals of care) before implementation. **[N/A]**

**Strength of Evidence**

**GRADE:** LOW

*9 source(s) retrieved · 3 limited-confidence finding(s) flagged for verification*

## Notes

### Competing Interest Statement

The authors have declared no competing interest.

### Funding Statement

This study did not receive any funding

### Summary of Updates

The manuscript was updated and the software program was updated to include isobolograms, nomograms, ishikawa diagrams, references, and multilayer rationales for the inference outputs.

